# Cellular and molecular heterogeneities and signatures, and pathological trajectories of fatal COVID-19 lungs defined by spatial single-cell transcriptome analysis

**DOI:** 10.1101/2023.02.24.23286388

**Authors:** Arun Das, Wen Meng, Zhentao Liu, Md Musaddaqul Hasib, Hugh Galloway, Suzane Ramos da Silva, Luping Chen, Gabriel L Sica, Alberto Paniz-Mondolfi, Clare Bryce, Zachary Grimes, Emilia Mia Sordillo, Carlos Cordon-Cardo, Karla Paniagua Rivera, Mario Flores, Yu-Chiao Chiu, Yufei Huang, Shou-Jiang Gao

## Abstract

Despite intensive studies during the last 3 years, the pathology and underlying molecular mechanism of coronavirus disease 2019 (COVID-19) remain poorly defined. Here, we examined postmortem COVID-19 lung tissues by spatial single-cell transcriptome analysis (SSCTA). We identified 18 major parenchymal and immune cell types, all of which are infected by SARS-CoV-2. Compared to the non-COVID-19 control, COVID-19 lungs have reduced alveolar cells (ACs), and increased innate and adaptive immune cells. Additionally, 19 differentially expressed genes in both infected and uninfected cells across the tissues mirror the altered cellular compositions. Spatial analysis of local infection rates revealed regions with high infection rates that are correlated with high cell densities (HIHD). The HIHD regions express high levels of SARS-CoV-2 entry-related factors including ACE2, FURIN, TMPRSS2, and NRP1, and co-localized with organizing pneumonia (OP) and lymphocytic and immune infiltration that have increased ACs and fibroblasts but decreased vascular endothelial cells and epithelial cells, echoing the tissue damage and wound healing processes. Sparse non-negative matrix factorization (SNMF) analysis of neighborhood cell type composition (NCTC) features identified 7 signatures that capture structure and immune niches in COVID-19 tissues. Trajectory inference based on immune niche signatures defined two pathological routes. Trajectory A progresses with primarily increased NK cells and granulocytes, likely reflecting the complication of microbial infections. Trajectory B is marked by increased HIHD and OP, possibly accounting for the increased immune infiltration. The OP regions are marked by high numbers of fibroblasts expressing extremely high levels of COL1A1 and COL1A2. Examination of single-cell RNA-seq data (scRNA-seq) from COVID-19 lung tissues and idiopathic pulmonary fibrosis (IPF) identified similar cell populations primarily consisting of myofibroblasts.

Immunofluorescence staining revealed the activation of IL6-STAT3 and TGF-²-SMAD2/3 pathways in these cells, which likely mediate the upregulation of COL1A1 and COL1A2, and excessive fibrosis in the lung tissues. Together, this study provides an SSCTA atlas of cellular and molecular signatures of fatal COVID-19 lungs, revealing the complex spatial cellular heterogeneity, organization, and interactions that characterized the COVID-19 lung pathology.

## Introduction

Multiple single-cell RNA-seq (scRNA-seq) analyses of coronavirus disease 2019 (COVID-19) patients with different severities have improved our understanding of cellular diversity associated with infection and provided important molecular insights into the host immune response [1–5]. Despite intensive studies in the last 3 years, the pathology and underlying molecular mechanism of COVID-19 remain unclear. Severe COVID-19 is often accompanied by diffuse alveolar damage (DAD) that presents complex pathological manifestations and is heterogeneous within infected tissues and across patients [6]. The dissociation of tissue localization of cells in scRNA-seq has become a bottleneck to decoding the pathology features at the molecular and cellular levels and failed to reveal the immune signatures of the microenvironment for severe COVID-19.

Spatial single-cell transcriptome analysis (SSCTA) promises to reveal the molecular basis of cellular heterogeneity, organization, and interactions in tissues and organs [7, 8]. However, analysis of these complex datasets including defining the spatial cellular organizations, immune microenvironment patterns, cell-cell interactions, and molecular signatures associated with disease pathophysiology remains a daunting task.

Here, we utilized SSCTA to examine postmortem lung tissues from 5 cases with severe COVID-19 and one case without COVID-19. From 10,414,863 detected transcripts of cellular 221 genes from six tissues, we identified 1,719,459 cells that were mapped to 18 major parenchymal and immune cell types, all of which are infected by SARS-CoV-2. We further identified the spatial cellular and molecular signatures that define the patterns of SARS-CoV-2 infection, structural and pathological presentations, and associated immune microenvironments, which project the trajectories of disease progression. Together, this study provides an atlas of cellular and molecular signatures of fatal COVID-19 lungs and reveals the complex spatial cellular heterogeneity, organization, and interactions that characterize COVID-19 lung pathology.

## Results

### Spatial single-cell transcriptome analysis, cell segmentation, cell typing, and spatial mapping of cells

Postmortem lung tissues from five COVID-19 autopsies and one postmortem case without COVID-19 were subjected to SSCTA (Fig. 1A). All five COVID-19 cases contracted SARS-CoV-2 in the first wave of the pandemic and had underlying conditions [9], including hypertension (cases 1-3), HIV infection and asthma (case 2), and Parkinson’s and chronic kidney diseases (cases 4 and 5). Hematoxylin and eosin (H&E) stains revealed various degrees of diffuse alveolar diseases (DAD), pulmonary thromboembolism, and lymphocytic infiltration in all cases (Fig. S1). Tissues 1 and 2 (1- 2C and 2-1A) had prominent organizing pneumonia (OP) or organizing diffuse alveolar damage while edema, hyaline membrane, and fibrin clot or microthrombi were prominent in tissues 3, 4, and 5 (3-1A, 4-3B and 5-3B). We designed probes for detecting the SARS-CoV-2 genome and 221 cellular genes covering markers of common lung parenchymal and immune cells, and immune and inflammatory genes induced by viral infections (Table S1). Following hybridization and gene decoding by *in-situ* sequencing, we stained the tissues with 4′,6-diamidino-2-phenylindole (DAPI) (Fig. S2) to facilitate cell segmentation (Fig. S2).

**Figure 1:**
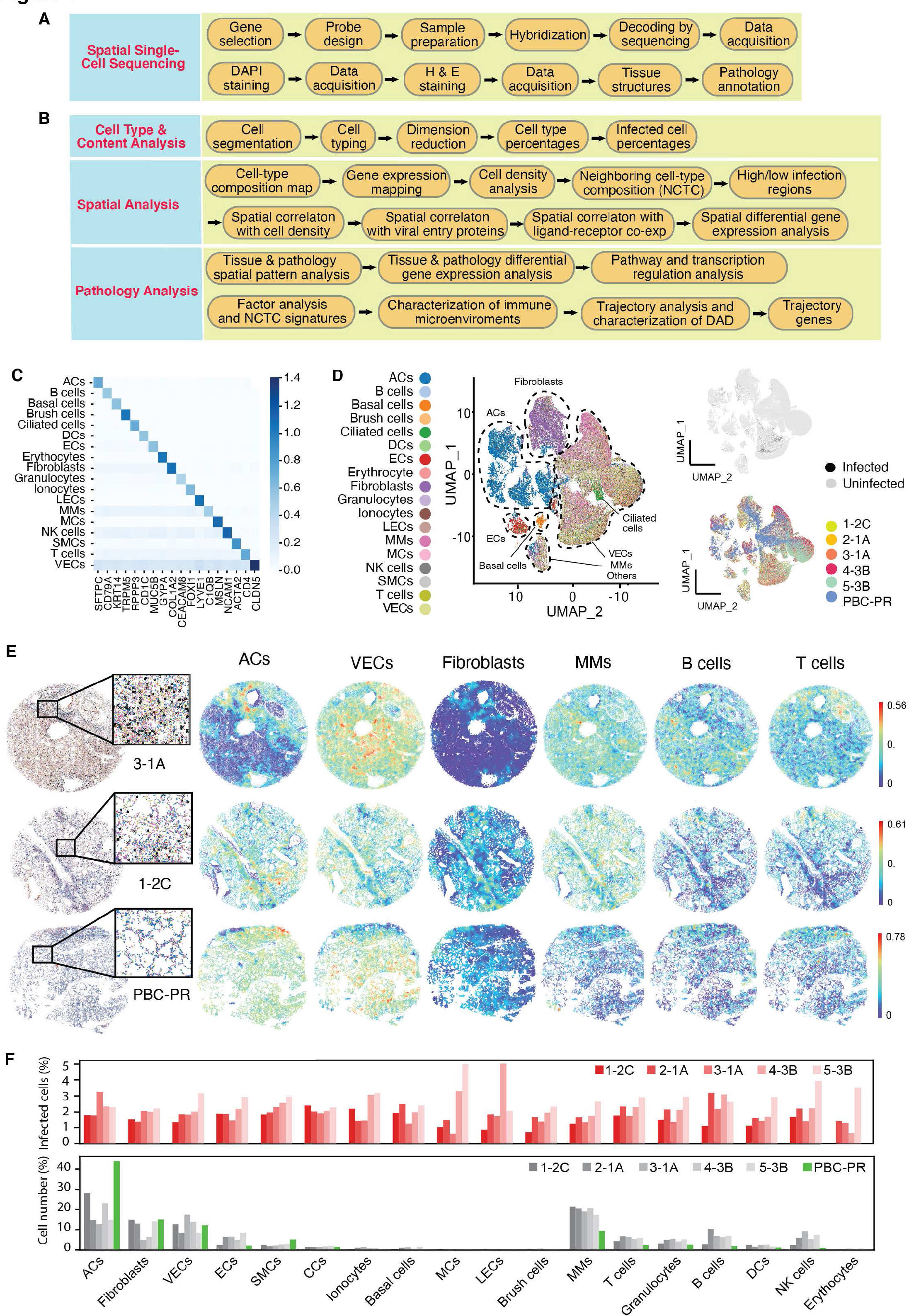
Spatial organization of parenchymal and immune cells in COVID-19 lung tissues revealed by spatial single-cell transcriptome analysis (SSCTA). A. Schematic illustration of the spatial single-cell sequencing pipeline used to generate the target dataset comprised of healthy control (non-COVID-19 tissue, n=1) and COVID-19 tissues (n=5). Samples were hybridized and decoded by sequencing. DAPI and H&E staining were carried out. All samples were annotated by an expert pathologist (Fig. S1). B. SSCTA workflow illustrating cell segmentation, cell typing, neighborhood cell- type composition (NCTC) analysis, and tissue pathology analysis. C. Average expression of cell type markers of all segmented cells across identified cell types in the SSCTA dataset. D. The UMAP projection of the gene expressions of segmented cells colored based on cell type, infection, and sample identifier. The UMAP shows negligible batch differences between the samples. E. Spatial visualization of identified cell types in two COVID-19 tissues and the non-COVID-19 tissue along with individual spatial plots illustrating the neighborhood cell type composition of specific cell types. We identified DAD in all COVID-19 tissues and orderly distribution of major parenchymal cells in non- COVID-19 tissue (PBC-PR). Further plots are provided in Fig. S1. F. Bar plots showing the percentages of identified cell types and their percentages of SARS-CoV-2 infected cells.

A workflow was developed to analyze the SSCTA data (Fig. 1B). We detected 869,453 to 3,424,675 reads in the six lung tissues for a total of 10,414,863 reads (Table S2 and Fig. S3A). Using the Baysor algorithm [10], we segmented ∼89%-95% of reads into cells based on DAPI staining and the spatial distribution of reads (Fig. S4). We identified 186,659 to 470,294 cells for each sample for a total of 1,719,459 cells in these tissues (Table S2 and Fig. S3B). Over 99% of the cells harbored at least 5-15 reads (Fig. S5). We further filtered the cells deemed low quality (see Methods) and retained a total of 779,137 cells for the subsequent analyses.

We summarized the reads for each gene and normalized the gene reads of segmented cells using *scTransform* [11]. We assigned cell types to 70-84% of the segmented cells based on the expressions of cell type-specific markers (Table S1) and identified a total of 18 cell types including 11 types of parenchymal cells and 7 types of immune cells in all six tissues (Fig. 1C and S6). We used Uniform Manifold Approximation and Projection (UMAP) to visualize the relationship of gene expressions of individual cells associated with different cell types in all tissues (Fig. 1D and S7).

There were negligible batch differences as cells from different samples mixed well in individual cell clusters (Fig. 1D and S7). We noticed separated clusters for major parenchymal cells including alveolar cells (ACs) and fibroblasts. By contrast, immune cells were grouped together and mixed with vascular endothelial cells (VECs) in several clusters (Fig. 1D). The poor separation of immune cell types was largely due to highly expressed immunoglobulin kappa light chain (IGKC) and cathepsin L (CTSL) in these cell types (Fig. S8).

We then mapped the individual cells to their spatial locations in the tissues (Fig. S9) and performed the neighborhood cell type composition (NCTC) analysis by computing a vector of 18 cell-type percentages in the neighborhood of individual cells. NCTC reveals spatial variations of the local cell type composition and informs local interplays among cell types. In agreement with the observed pathology that showed DAD in all COVID-19 tissues (Fig. S1), NCTC uncovered disorganized distributions of various cell types, especially the major parenchymal cells such as ACs, VECs, and fibroblasts (Fig. 1E, S1, and S10-S14). In contrast, organized structures and orderly distributions of these cell types were observed in the non-COVID-19 tissue (PBC-PR) (Fig. 1E, S1, and S15). Examination of regions with blood vessels revealed the lining of VECs along the vessels together with other vessel-associated cells including ACs, fibroblasts, and smooth muscle cells (SMCs) (Fig. 1E and S16). Furthermore, we observed abundant infiltrating immune cells including macrophage and monocytes (MMs), natural killer (NK) cells, and T- and B-cells along regions of blood vessels (Fig. 1E and S10-S14). We also observed the expected cell compositions of other structures including bronchiole, capillary, and endothelium (Fig. S17). These results validated the marker-based cell typing approach.

### SARS-CoV-2 infection alters the cell compositions of major parenchymal cells and induces immune infiltrations

In agreement with the results of our previous study [9], we found SARS-CoV-2 reads in diverse cell types with infection rates ranging from 0.6% to 5% except that no infected cell was detected in the small number of erythrocytes (55) identified in tissue 1 (Fig. 1D, 1F, S7 and Table S3). Most infected cells were identified as ACs, fibroblasts, VECs, and MMs as they were the major cell types in the COVID-19 tissues (Fig. 1F).

Consistent with the tissue pathology, we observed significant alterations in the compositions of different cell types in COVID-19 tissues compared to the non-COVID- 19 tissue (Fig. 1E and 1F). Among the parenchymal cells, there were reduced numbers of ACs from 44.08% to 12.81-28.23% and SMCs from 5.22% to 1.72-3.05%, and increased numbers of epithelial cells (ECs) from 2.08% to 2.40-8.42%, ionocytes from 0.22% to 0.41-1.36% and basal cells from 0.25% to 0.31-1.58%, suggesting their likely involvements in COVID-19 lung pathology (Fig. 1E, 1F, and Table S3). Furthermore, we observed increases in cell numbers of most types of the identified immune cells in COVID-19 tissues (Fig. 1E, 1F, and Table S3). In particular, there were increased inflammatory cells including MMs from 9.4% to 17.51-21.37% and NK cells from 1.00% to 2.32-9.19%, and adaptive immune cells including T cells from 2.40% to 4.34-6.81% and B cells from 1.91% to 2.70-10.51%, which were consistent with the reported inflammatory and cellular immune response following SARS-CoV-2 infection [12].

However, these changes were more subtle for some cell types in tissue 1, which had the least infected cells and mildest pathological manifestations with the best integrity of parenchymal cells among all COVID-19 tissues (Fig. 1E, 1F, S1, and S10).

### SARS-CoV-2 infection induces global differential gene expressions that marked pathological damages and inflammation with spatial cellular features

We examined the differential gene expression between COVID-19 and non-COVID-19 lung tissues, and identified 9 upregulated and 10 downregulated genes, respectively (Fig. 2A, first panel). Comparison of SARS-CoV-2-infected cells in COVID- 19 tissues with all cells in the non-COVID-19 tissue confirmed the differential expressions in 16 of these 19 genes (Fig. 2A, second panel). Interestingly, differential gene expressions were also observed in 18 of these 19 genes between uninfected cells in COVID-19 tissues and all cells in the non-COVID-19 tissue (Fig. 2A, third panel). In contrast, there were only 3 differentially expressed genes between SARS-CoV-2- infected and -uninfected cells in the COVID-19 tissues (Fig. 2A, fourth panel). These results indicated that there were strong indirect effects such as those mediated by cytokines and complement activation induced by SARS-CoV-2 infection that likely contributed to the dysregulated gene expressions and pathology in COVID-19 lung tissues. Thus, we focused on comparing COVID-19 tissues with the non-COVID-19 tissue in subsequent analyses of dysregulated genes.

**Figure 2:**
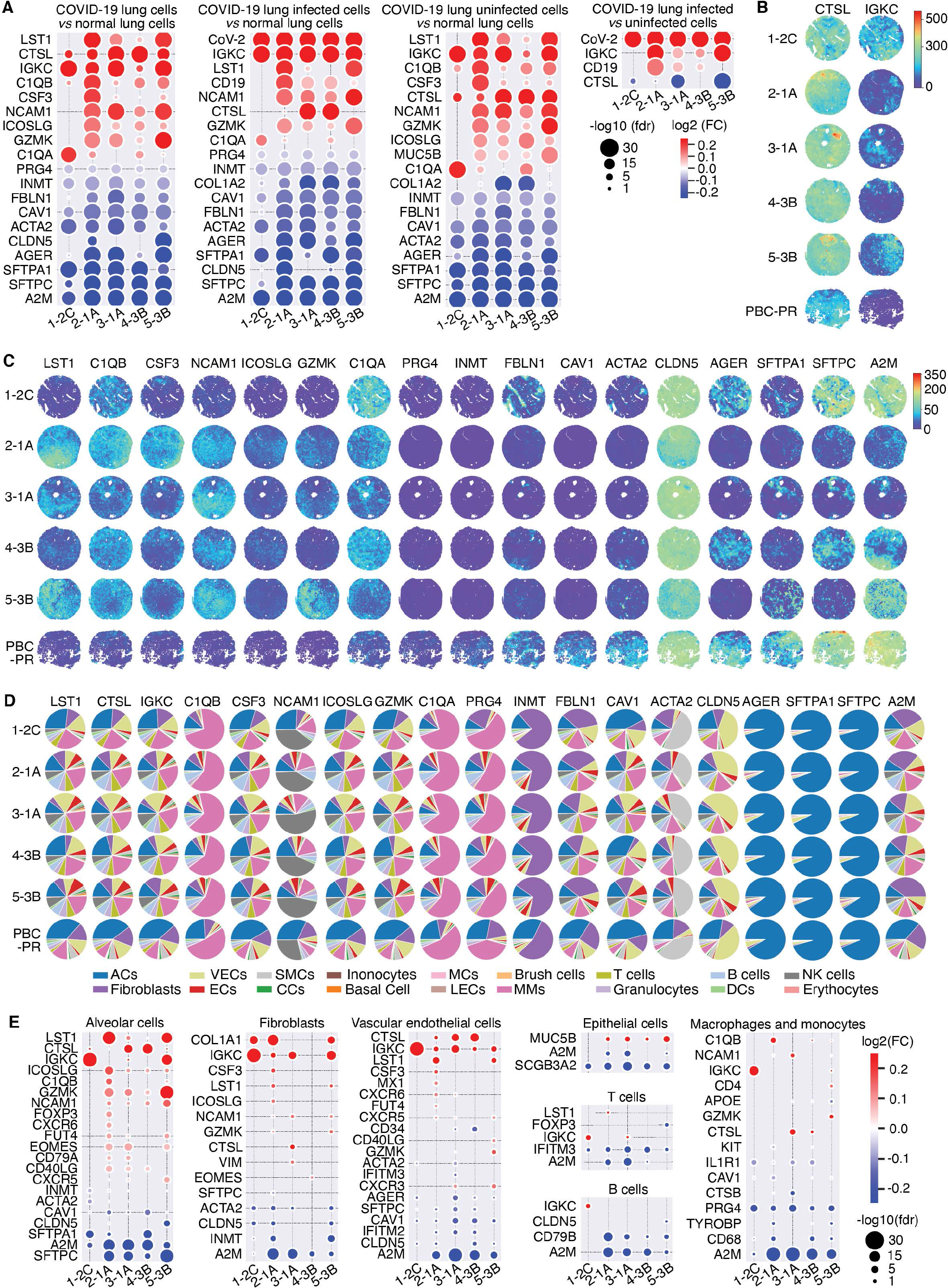
SARS-CoV-2 infection induces global differential gene expression that mediates pathological damages and inflammation with cell type and spatial features. A. Differential gene expressions between the identified cells in the COVID-19 non-COVID-19 tissue with various conditions are illustrated. The title indicates the conducted differential expression analysis. Within each row, the bubble size indicates the -log10 of the corrected p-value and the color indicates the log2 fold change of the corresponding gene. Each column indicates the different COVID-19 tissues. The first three panels indicate differential expression analysis between COVID-19 tissues and non-COVID-19 tissue while the fourth panel indicates the differential expression analysis between the infected and uninfected cells in the respective COVID-19 tissues. B. Spatial gene expression map of highly expressed CTSL and IGKC. C. Spatial gene expression maps of the rest of the differentially expressed genes. D. Pie chart illustrating the percentages of cell types in which the genes are expressed in each tissue. E. Differential expression analysis of COVID-19 tissues compared to non- COVID-19 tissue in cell types that had significant changes in cell numbers based on Fig. 1F.

The expressions of the 19 dysregulated genes in different cell types and their tissue distributions had significant variations suggesting their complex involvements in different aspects of SARS-CoV-2 infection and COVID-19 lung pathology (Fig. 2B, 2C, S8, and S18-S23). We determined the contributions of individual cell types to the expressions of these genes across all the tissues (Fig. 2D). Of the 19 genes, 10 of them were cell markers (Table S1). All 6 parenchymal cell markers including SFTPC, SFTPA1, and advanced glycosylation end product (AGE) receptor (AGER) for ACs, INMT for fibroblasts, smooth muscle actin alpha 2 (ACTA2) for SMCs, and claudin 5 (CLDN5) for VECs were downregulated, and mainly expressed in their respective cell types (Fig. 2A, first panel, 2C, 2D, S8 and S18-S23), of which ACs and SMCs had decreased cell numbers in COVID-19 tissues (Fig. 1E, 1F and S7). Both SFTPC and SFTPA1 are implicated in lung homeostasis and functions, and their mutations and dysregulation are associated with pulmonary fibrosis [13–15]. Additionally, SFTPA1, a C-type lectin, which binds to specific carbohydrate moieties on lipids and the surface of microorganisms, is essential in the defense against respiratory pathogens [16–18] while the pulmonary-associated surfactant protein C encoded by SFTPC is a marker for COVID-19 patients with high viral loads [19]. AGER, predominantly expressed in ACs (Fig. 2C), is a multiligand receptor interacting with AGE and other molecules implicated in lung homeostasis, development and inflammation, and certain diseases such as diabetes and Alzheimer’s disease, and regulates diverse pathways including MyD88-dependent, nuclear receptors, TNF-α, ERK1/2 and p38 MAPK, and p53/TP53 pathways [20–23]. Following interaction with S100A12, AGER triggers the activation of mononuclear phagocytes, lymphocytes, and endothelium by generating key pro- inflammatory mediators [24]. Indeed, AGER-related pathways are activated by SARS- CoV-2 infection and are implicated in the COVID-19 lung pathology [25, 26]. ACTA2 encodes a smooth-muscle actin involved in lung functions including cell motility, tissue structure and integrity, and intercellular signaling [27]. Dysregulation of ACTA2 is linked to a variety of vascular diseases as well as multisystemic smooth muscle dysfunction syndrome [28, 29]. CLDN5 is an integral membrane protein and component of tight junction strands regulating the integrity of epithelial or endothelial cell sheets, and immune cell transmigration [30–32]. SARS-CoV-2 infection suppressed CLDN5 expression contributing to the disruption of respiratory vascular barriers [33]. Thus, downregulations of parenchymal cell markers including SFTPC, SFTPA1 and AGER in ACs, ACTA2 in SMCs, and CLDN5 in VECs might be involved in SARS-CoV-2 infection and COVID-19 lung pathology (Fig. 2D).

Several other downregulated genes including caveolin 1 (CAV1) and alpha-2- macroglobulin (A2M) are implicated in numerous lung functions (Fig. 2A). CAV1, a component of caveolae involved in multiple pathways such as integrins-mediated and Ras-ERK signaling, is essential for lung functions and during the host response to infections [34, 35]. A2M is an inhibitor of broad spectrum of proteases including trypsin, thrombin, and collagenase [36, 37]. It disrupts inflammatory cascades by inhibiting inflammatory cytokines, is implicated in Alzheimer’s disease, and regulates extracellular matrix organization and platelet cytosolic Ca2+ [36, 37]. CAV1 and A2M were downregulated and mainly expressed by ACs, fibroblasts, VECs, and MMs (Fig. 2A, first panel, 2D, S8, and S18-S23), of which ACs and fibroblasts had decreased while MMs had increased cell numbers in COVID-19 tissues (Fig. 1F).

Three upregulated immune marker genes including complement component C1q, A chain (C1QA) and B chain (C1QB), and neural cell adhesion molecule 1 (NCAM1) are involved in innate immune response and inflammation (Fig. 2A, first panel) [38, 39].

C1QA and C1QB were predominantly expressed in MMs (Fig. 2D, S8 and S18-S23). SARS-CoV-2 infection induced robust complement activation, contributing to tissue damage and COVID-19 lung pathology [40–42]. Both cathepsin L (CTSL) and granzyme K (GZMK) could be involved in complement activation [43, 44]. CTSL was induced after SARS-CoV-2 infection and enhanced SARS-CoV-2 infection [45] while GZMK, a serine protease from the cytoplasmic granules of cytotoxic lymphocytes (CTL) and NK cells that recognize and lyse specific target cells [46], could limit the spread of SARS-Co-V-2.

Both CTSL and GZMK were upregulated and mainly expressed by ACs, fibroblasts, MMs, and VECs while CTSL exhibited a comparable expression level but with extremely high levels in localized regions in COVID-19 tissues (Fig. 2A, 2B, 2C, 2D, S8 and S18-S23). NCAM1, the marker of NK cells, is a member of the immunoglobulin superfamily involved in cell-to-cell interactions as well as cell-matrix interactions [47].

The upregulations of C1QA, C1QB, CTSL, GZMK, and NCAM1 were consistent with the increased numbers of MMs and NK cells in COVID-19 tissues (Fig. 1F, S8, and S18- S23). Thus, increased infiltrations of MMs and NK cells were likely involved in COVID- 19 lung complement activation and inflammation.

Two immune-modulating cytokines were upregulated including colony-stimulating factor 3 (CSF3), a member of the IL-6 superfamily of cytokines that controls the production, differentiation, and function of granulocytes involved in the innate immune response [48, 49], and inducible T cell costimulator ligand (ICOSLG or B7-H2) involved in positive regulation of interleukin-4 production and CD28 signaling in T-helper cells [50, 51]. The major cells that expressed CSF3 and its receptor CSF3R, as well as ICOSLG and its receptor ICOS, were ACs, fibroblasts, VECs, and MMs (Fig. 2D).

Numerous other upregulated genes in COVID-19 tissues are involved in inflammatory and immune responses (Fig. 2A). Leukocyte-specific transcript 1 (LST1), a myeloid leukocyte-specific transmembrane adaptor protein recruiting protein tyrosine phosphatases SHP-1 and SHP-2 to the plasma membrane, inhibits the proliferation of lymphocytes, and its expression is enhanced by lipopolysaccharide (LPS), interferon- gamma (IFN-γ) and bacteria [52, 53]. Immunoglobulin kappa light chain (IGKC) is essential for antibody production but is also expressed in non-lymphoid cells [54–56].

Both LST1 and IGKC were mainly expressed in ACs, fibroblasts, VECs and MMs (Fig. 2B-E, S8 and S18-S23), which might mediate dysregulation of immune cells in COVID- 19 lung tissues. Finally, fibulin 1 (FBLN1), a secreted glycoprotein incorporated into a fibrillar extracellular matrix in a calcium-dependent manner and mediates platelet adhesion via binding fibrinogen [57, 58], was mainly expressed in ACs, fibroblasts, VECs, and MMs (Fig. 2D).

We examined differential gene expressions in cell types that had significant changes in cell numbers (Fig. 1F). Compared to non-COVID-19 tissue, ACs from COVID-19 tissues had 20 differentially expressed genes, of which 14 were identified at the whole tissue level (Fig. 2A, first panel and 2E). The most upregulated genes were LST1, CTSL, IGKC, ICOSLG, C1QB, GZMK, and NCAM1 while the most downregulated genes were SFTPC, A2M, SFTPA1, CLDN5, and CAV1 (Fig. 2E), which likely contribute to the decreased cell numbers in COVID-19 tissues (Fig. 1F). Several differentially expressed genes were also identified in fibroblasts, VECs and ECs (Fig. 2E). Among the immune cells, MMs had the most differentially expressed genes including upregulation of C1QB, NCAM1, and IGKC, and downregulation of A2M, CD68, TYROBP and PRG4 (Fig. 2E).

### Spatial analysis identifies regions with high SARS-CoV-2 infection rates that match high cell densities, high levels of viral entry-related factors, and localized pathology

Even though we did not observe notable global differences in gene expression between infected and uninfected cells within each COVID-19 tissue, the spatial nature of the SSCTA permitted the identification of the local impact of SARS-CoV-2 infection on lung pathology. We performed the local infection rate analysis to examine the spatial distributions of local SARS-CoV-2 infection (Fig. 3A) and identified regions with high and low infection rates in each tissue (Fig 3B; local Moran’s I, p-value < 0.05). SARS-CoV-2 infects host cells by binding to its entry receptor, angiotensin-converting enzyme 2 (ACE2), and subsequent engagement of host proteases and other entry-related factors including FURIN, transmembrane protease, serine 2 (TMPRSS2), and neuropilin 1 (NRP1) [59]. We assessed the spatial correlation between high-infection regions and cells expressing transcripts of these viral entry-related proteins. We found a significant correlation between high infection rates and high expressions of ACE2, FURIN, TMPRSS2, and NRP1 (Fig. 3C; bivariate local Moran’s I, p-value < 0.05), supporting the essential roles of these cellular proteins in SARS-CoV-2 infection in lung tissues.

**Figure 3:**
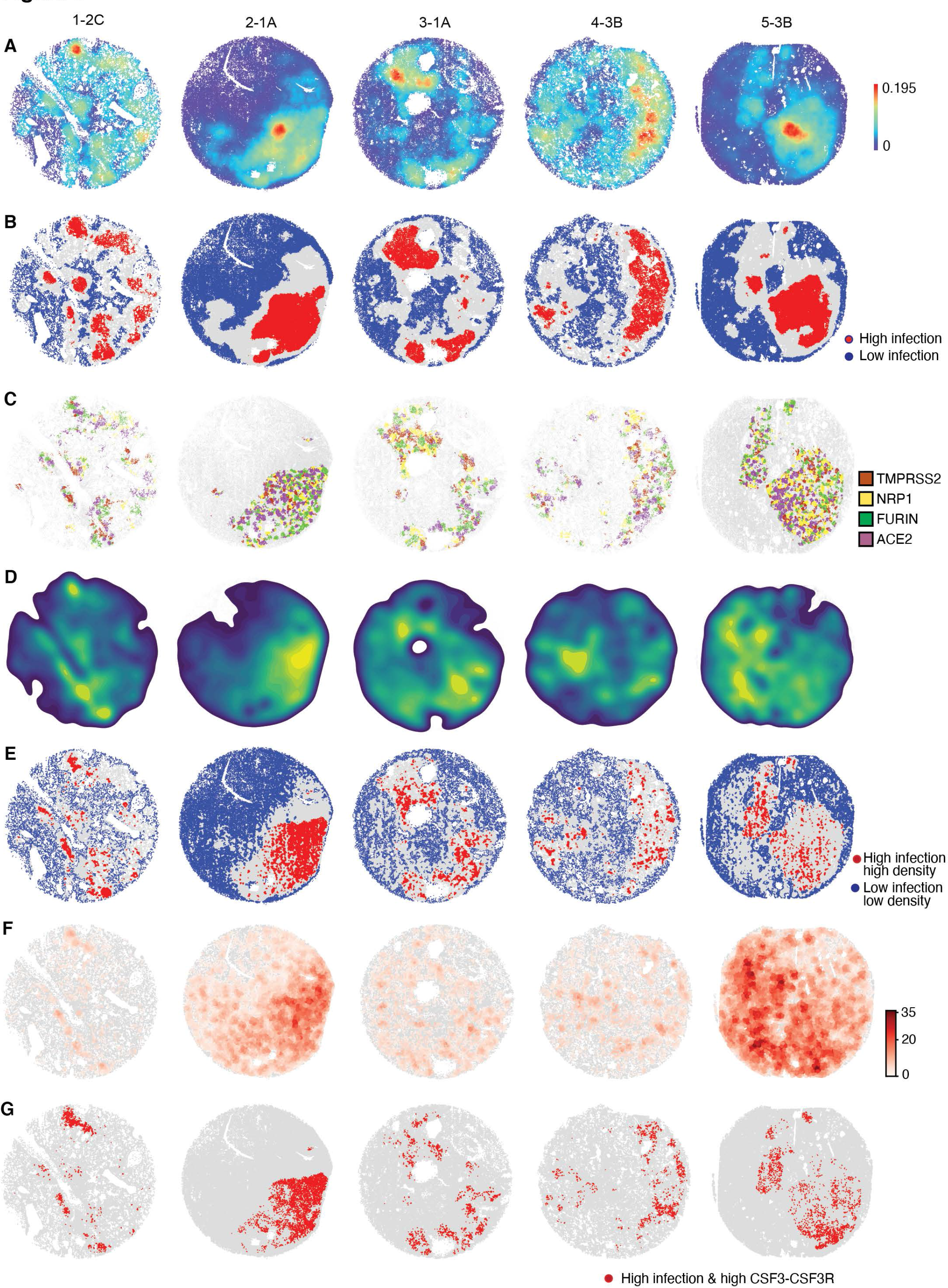
Spatial analysis of local SARS-CoV-2 infection rates identifies regions with high infection rates that match high cell densities, high levels of viral entry- related factors, and localized pathology. A. Spatial distribution of the SARS-CoV-2 infected cells. The spatial visualization of the infection rates revealed infection hotspots in the COVID-19 tissues. B. Moran’s I analysis of spatial patterns revealed spatially distinct high and low infection regions in each COVID-19 tissue. C. Spatial visualization of the significant local regions identified by Bivariate Moran’s I analysis, in which infection rates and the expressions of TMPRSS2, NRP1, FURIN and ACE2 are spatially correlated. D. Spatial visualization of the kernel density plot for the cell densities revealed highly dense cellular hotspots in the COVID-19 samples. E. Spatial visualization of the significant local regions identified by Bivariate Moran’s I analysis, in which cell densities and the infection regions are spatially correlated, identified as high- density high-infection (HIHD) regions. F. Spatial visualization of the ligand-receptor coexpression maps for the CSF3-CSF3R pair, which are highly expressed in the high infection regions. G. Spatial visualization of the significant local regions identified by Bivariate Moran’s I analysis, in which both high infection and high CSF3-CSF3R coexpression regions are spatially correlated.

Previous studies reported increased densities of both parenchymal and immune cells in COVID-19 lung tissues [6]. Here, we observed a positive spatial correlation between cell densities and SARS-CoV-2 infection rates especially in tissues 1-4 (Fig. 3D, Table S4), suggesting that increased cell densities might be associated with high infection rates. We segmented regions with high infection rates and high cell densities (HIHD) (Fig. 3E; bivariate local Moran’s I, p-value < 0.05). Compared with H&E staining (Fig. S1), we found a close association between HIHD and OP, which was most prominent in tissues 1 and 2, as well as lymphocytic and immune infiltration, which was most severe in tissues 3 and 4 (Fig. 3E). Both are the most common lung pathological manifestations in COVID-19 lungs [12]. Hence, high infection rates might cause persistent damages to the lung tissues, resulting in the infiltration of immune cells, and increased wound repair and fibrosis [60, 61]. We also observed HIHD areas adjacent to blood vessels in tissues 1 and 3 (Fig. 3E and S1), which might be due to the homing of the circulating virus and immune cells in the bloodstream to these sites or that these cells were more susceptible to SARS-CoV-2 infection [9, 62].

### Regions with high SARS-CoV-2 infection rates have unique cellular and gene expression features

We next examined cell type compositions of the high and low SARS-CoV-2 infection regions. Compared to low-infection regions across the tissues, the compositions in high-infection regions exhibited significant increases in fibroblasts, echoing the observed OP in HIHD areas, and an increase in ACs, possibly due to the proliferation of ACs response to injury repair of alveoli and alveoli capillaries (Fig. 3E and 4A). However, there were decreases in ECs and VECs, which might reflect the damages caused by the infection (Fig. 3E and 4A). These results suggested that high-infection regions might suffer more damages. However, except for T cells and granulocytes, the compositions of major immune cell types including MMs between high and low infection regions remained similar (Fig. 4A), suggesting induction of a broad immune infiltration by SARS-CoV-2 infection, as well as the observed broad impact on the tissue-wide gene expression (Fig. 2A).

**Figure 4:**
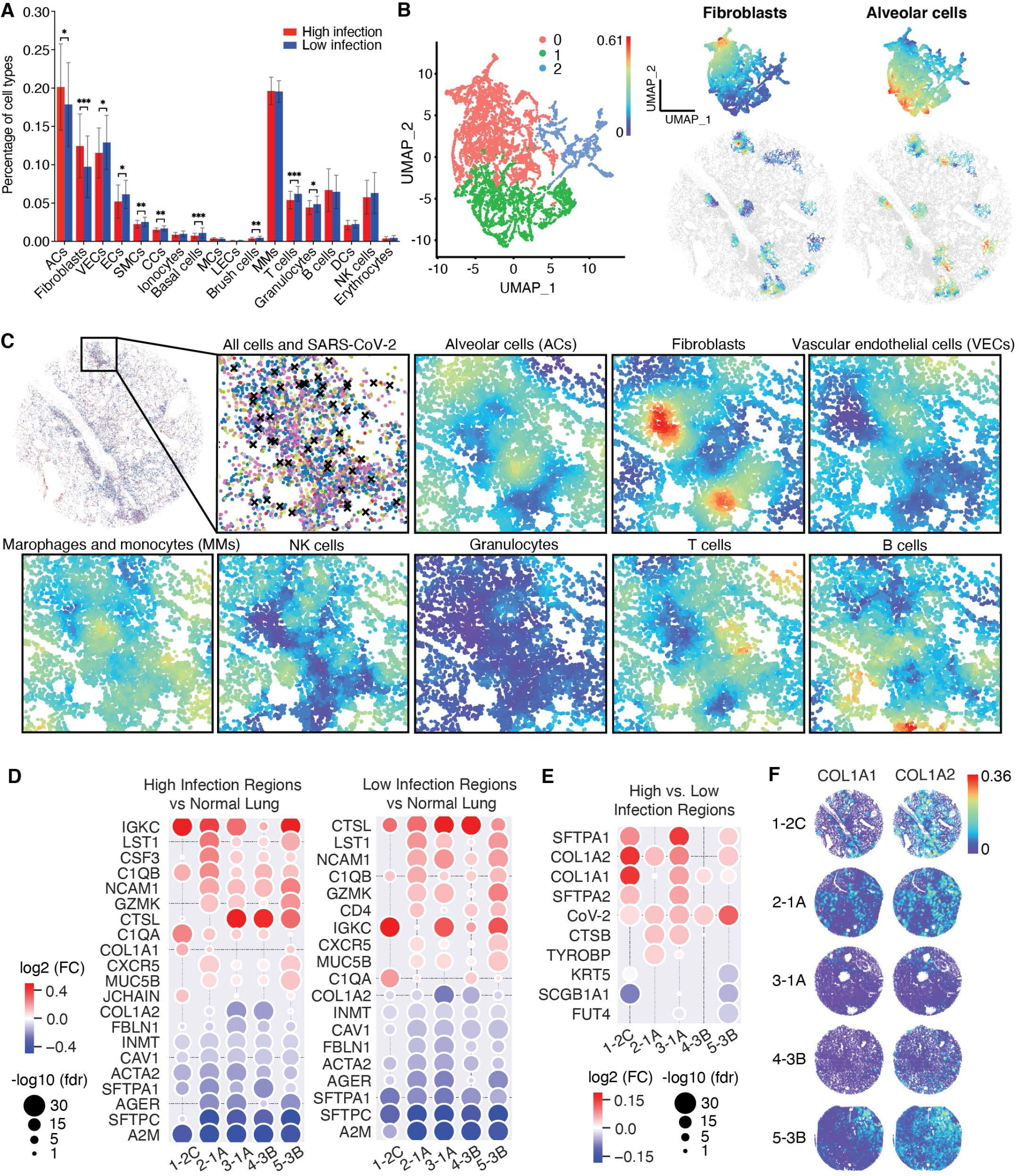
Spatial analysis reveals cellular and gene expression features in regions with high SARS-CoV-2 infection rates. A. A bar plot showing the cell type composition of all identified cell types in the high and low infection regions. The increase in alveolar cells (ACs) might suggest the proliferation of ACs due to injury repair of alveoli and alveoli capillaries and a decrease in epithelial cells (ECs) and vascular endothelial cells (VECs) might suggest the damages caused by the infection. **B.** UMAP projection of the NCTC analysis results of high infection regions (refer to figure 3B) revealed three distinct clusters out of which two were distinctly enriched in local compositions of ACs and fibroblasts, which are illustrated on the right with their spatial visualizations. **C.** Spatial visualization of the compositions in high infection regions revealed negative correlation of fibroblasts and ACs (-0.6, Pearson correlation) which overlapped with annotated OP regions (high fibroblasts, low ACs). **D.** Differential expressions of high and low infection regions of the COVID-19 tissues compared to the non-COVID-19 tissue, which share 18 of the 20 differentially expressed genes, confirming strong indirect effect of SARS-CoV-2 infection. **E.** Differential expressions between the high and low infection regions of the COVID-19 tissues revealed distinct differentially expressed genes showing upregulation of AC markers SFTPA1 and SFTPA2 in tissues 1, 3, and 5, and fibroblast markers COL1A1 or COL1A2 in all COVID-19 tissues. **F.** Spatial visualization of the spatial gene expression map of COL1A1 and COL1A2 markers.

The patchwork of high infection regions in tissue 1 prompted us to inspect patterns of local cell-type compositions. Interestingly, NCTC analysis showed three clusters in high-infection regions, of which two were distinctly enriched with prominent local compositions of fibroblasts and ACs, respectively (Fig. 4B). Examining their spatial distributions confirmed a negative *in-situ* correlation between local compositions of fibroblasts and ACs (-0.6, Pearson correlation), with regions showing high (low) compositions of fibroblasts accompanied with low (high) compositions of ACs (Fig. 4B and C). Strikingly, the local regions with high fibroblasts but low ACs were mostly annotated OP regions (Fig. S1).

We next investigated the gene expression patterns in the high- and low-infection regions. Compared with the normal tissue, the high and low infection regions presented highly consistent differential expression patterns as they shared 18 of the 20 differentially expressed genes (Fig. 4D), confirming the strong indirect effect of SARS-CoV-2 infection (Fig. 2A). One of the two uniquely differentially expressed genes in high infection regions was again cytokine CSF3, which was upregulated in all infected tissues. We, therefore, investigated spatial co-expression of CSF3 and its receptor CSF3R (Fig. 3F) and found that their co-expression patterns also presented spatial correlations in high infection regions (Fig. 3G; bivariate local Moran’s I, p-value < 0.05), suggesting that the CSF3-CSF3R axis might modulate the immune response in high-infection regions. In contrast to the consistent differentially expressed patterns of both high- and low-infection regions when compared with the normal control, direct comparison between high- and low-infection regions revealed distinct differentially expressed patterns. Particularly, we observed, in high- *vs.* low-infection regions, upregulation of AC markers SFTPA1 or SFTPA2 in tissues 1, 3, and 5 (Fig. 4E and 2C) and fibroblast markers COL1A1 or COL1A2 in all COVID-19 tissues (Fig. 4E and 4F), despite that SFTPA1 and COL1A2 displayed down-regulation in either high- or low-infection regions in COVID-19 tissues when compared to the normal tissue (Fig. 4D).

The increased expressions of these AC and fibroblast markers were also consistent with the observed increases in ACs and fibroblasts in high-infection regions in the corresponding tissues (Fig. 4A). Cell type-specific differential expressions further confirmed the upregulation of SFTPA1 or SFTPA2 in ACs, and COL1A2 in fibroblasts in infected tissues (Fig. 4E and S24). However, different cell types exhibited complex differential expression patterns between high *vs.* low infection regions in individual COVID-19 tissues (Fig. S24), likely due to the distinct COVID-19 pathologies or stages associated with the individual infected tissues.

### Sparse non-negative matrix factorization analysis identifies seven cell composition signatures that recapitulate different healthy and disease statuses

Although different DAD phases can inform COVID-19 severity, they are confounded by many factors including the underlying conditions of the patients as illustrated by our five COVID-19 cases (Fig. S1). Importantly, the complex tissue microenvironments of parenchymal and immune cells associated with different stages of DAD within and across tissues have not been well characterized. Therefore, we determined the extent to which tissue-independent and tissue-specific spatial patterns in spatially resolved cell-type compositions are associated with different DAD stages. For this purpose, we performed the sparse non-negative matrix factorization (SNMF) [63] of the NCTC vectors of all cells in all tissues. SNMF decomposed these NCTC vectors into a sparse linear combination of cell-type composition signatures that define spatial cell-type patterns in these tissues. We obtained a factorization of seven NCTC signatures after assessing the compactness and biological meanings of the factorization results for different maximum numbers of factors (Fig. 5). Close examination of cell type compositions of the signatures (Fig. 5A and Table 5) and their spatial prevalence across all tissues (Fig. 5B) defined by the factor loading revealed their associations with normal structures, and broad and tissue-specific infection. Specifically, Signature 1 likely defined the cell type composition of “normal-like lung alveoli” because it has 56.4% ACs, 18.5% VECs, 11.9% fibroblasts, and 5.3% SMs, which were comparable to the percentages of the non-COVID-19 tissue (Fig. 1F and Table S3). Thus, the high-loading regions of Signature 1 in COVID-19 tissues might mark the less damaged regions.

**Figure 5:**
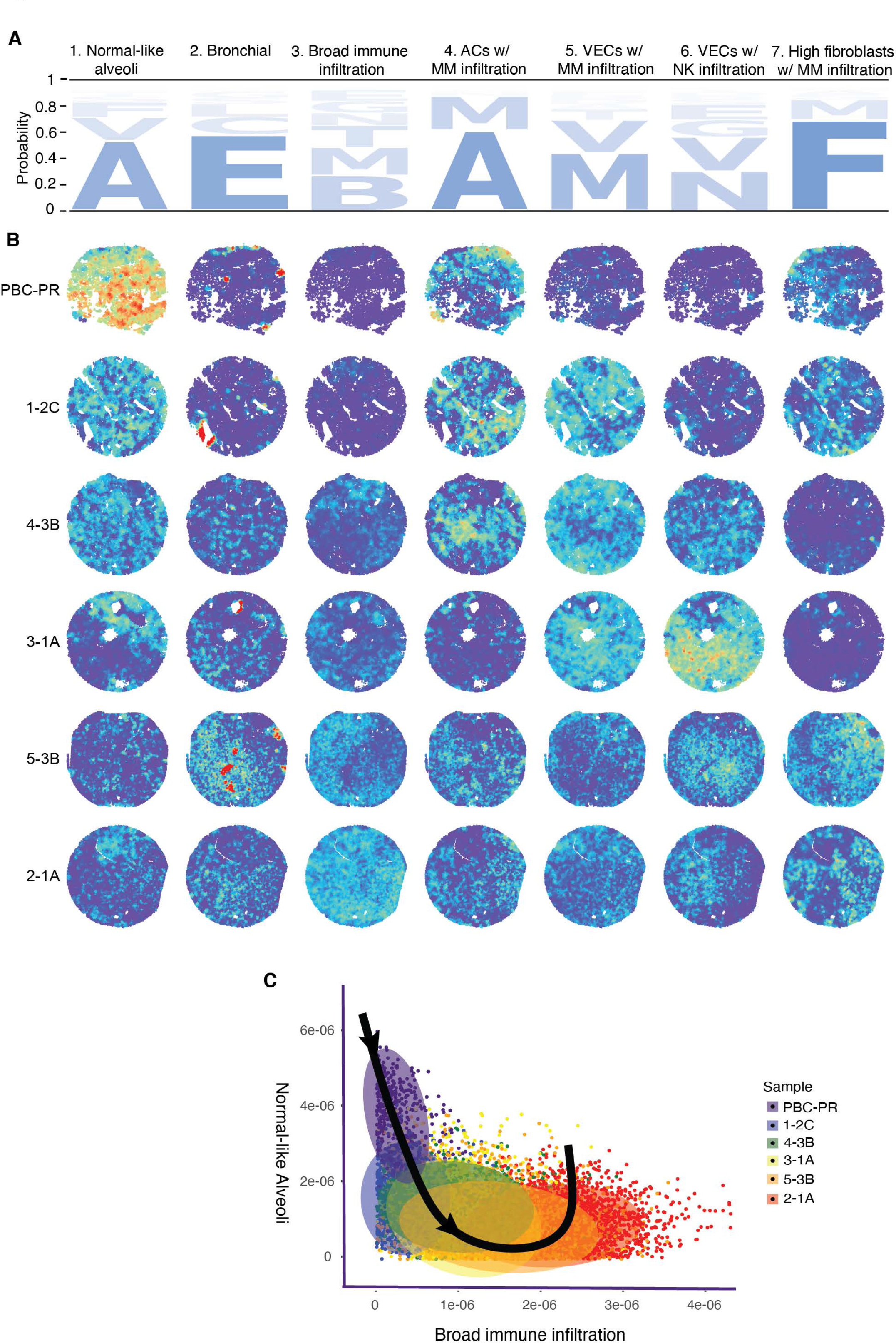
Sparse non-negative matrix factorization (SNMF) identifies seven cell composition signatures that recapitulate different healthy and disease statuses. **A.** SNMF revealed seven NCTC signatures which are illustrated in a logo plot. **B.** Spatial NCTC signatures visualized after factor loading for each tissue revealed their associations with normal structures and broad and tissue-specific infection. **C.** Trajectory analysis of the cell type compositions with Signatures 1 and 3 generated a trajectory of reduced normal alveoli and increased immune infiltration from the non- COVID-19 tissue to COVID-19 tissues 1, 4, 3, 5, and 2. The trajectory agrees with the order of the severity of pathology in the tissues identified in the H&E stained images (Fig. S1).

Naturally, the non-COVID-19 tissue had a high prevalence of Signature 1 (Fig. 5B and S25). Tissue 1 had the highest prevalence of Signature 1 among all COVID-19 tissues reflecting its least damaged pathology, followed by tissue 4, 3, 2, and 5 (Fig. 5B, S1, and S25). Signature 2 described a composition of 61% ECs, 14% ciliated cells, and 10% basal cells. It had sparse but high-intensity loadings in localized regions identified as bronchial tubes (Fig. 5B and S1). In contrast to Signatures 1 and 2 which characterized cell-type compositions of normal structures, Signature 3 captured the “broad immune infiltration” as it contained 28.6% B cells, 22.3% MMs, 17.5% T cells, 9.6% granulocytes, and 8.7% ECs (Fig. 5A and Table S5). As expected, it was the least common in the non-COVID-19 tissue (Fig. 5B and S25). Its loading distributions in COVID-19 tissues were consistent with the lowest immune infiltration in tissue 1, followed by 4, 3, 2, and 5 (Fig. 5B and S25), closely corroborating the order of damage revealed by Signature 1. Trajectory analysis of the cell type compositions with Signatures 1 and 3 using *Slingshot* [64] defined a trajectory of reduced normal alveoli and increased immune infiltration from the non-COVID-19 tissue to tissue 1, 4, 3, 5, and 2 (Fig. 5C), which was in agreement with the order of their increased severity of pathology observed in H&E staining (Fig. S1). Remarkably, Signatures 4 to 7 captured four distinct types of immune microenvironments or niches in these COVID-19 tissues. Signature 4 was characterized by high ACs (64.8%) with MMs (27.3%), likely reflecting the infiltration of alveolar MMs and proliferation of ACs in wound healing, all of which are features of exudative DAD [65]. The relatively higher prevalence of Signature 4 in the less severe tissues 1 and 4 suggested its association with early phases of DAD (Fig. 5B and S25). There were also overlaps in regions with higher loadings of Signature 4 with the annotated regions of immune infiltration in the H&E image in tissue 4 (Fig. 5B and S1). Signatures 5 and 6 depicted inflammations around VECs, where Signature 5 was associated with extensive infiltration of MMs (46.7%) and T cells (8.7%) around VEC (26.3%), whereas Signature 6 was associated with substantial infiltration of NK cells (31.8%) and granulocytes (13.6%) around VECs (27.6%) and ECs (12.2%). These results were consistent with the influx of MMs and NK cells (Fig. 1E and F), which was likely due to the activation of complement and coagulation systems (Fig. 2A and C), resulting in damages and therefore reduced numbers of ACs and VECs (Fig. 1E and F). Signature 5 was more common in the less severe tissues 1, 3 and 4 but Signature 6 appeared predominantly in 3 (Fig. 5B and S25), particularly in low infection region (Fig. 3A). Indeed, lymphocytic infiltrations were also identified in the regions associated with Signatures 5 and 6 (Fig. S1). Signature 7 denotes extremely high fibroblasts (73.4%) with infiltration of MMs (15.8%). Advanced disease in COVID-19 patients often has an excessive accumulation of fibroblasts and may develop pathological fibrosis due to chronic inflammation from the infiltrated immune cells and dysregulation of the extracellular matrix triggered (ECM) remodeling [66]. As expected, there was a high correlation between the COL1A1 expression level (Fig. 4F) with Signature 7. The loadings of this signature were high in tissues 1 and 2, which were annotated as OP regions, and in tissue 5, which were annotated as fibrin and hyaline membrane in addition to OP regions (Fig. 5B, S1 and S25), suggesting a connection with OP, fibrin and hyaline membrane.

### Unique cell composition signatures define two separate pathological trajectories

We then applied Signatures 4 to 7 to delineate the trajectories of DAD characterized by the immune responses against viral infection. We added Signature 1 “normal-like alveoli” so that the non-COVID-19 tissue could be included as a reference. We used these 5 signatures to redefine the landscape of spatially resolved cell type compositions in these tissues. The UMAP visualization (Fig. 6A) recapitulated the tissue severity inferred by Signatures 1 and 3 (Fig. 5C) but presented more complex patterns, with each immune signature defining a largely distinct group of cell populations (Fig. 6B). We applied *Slingshot* to infer pseudo-progressions of cells defined by these signatures and identified two trajectories (Fig. 6A). Clustering of cells based on pseudo- time values further segmented the trajectories into 6 stages (Fig. 6A). At the onset, the two trajectories shared a common path including stages T1 and most T2, which traversed the cells enriched by Signature 1 “normal-like alveoli” and Signature 4 “ACs with MMs infiltration” in T1 and then those enriched by Signature 4 and Signature 5 “VECs with MMs infiltration” in T2 (Fig. 6A). These stages were consistent with the less damaged presentations of tissues 1 and 4 (Fig. 6D and S1) and showed a reduction of ACs due to likely lung injury and an increase of MMs possibly promoted by innate immune responses (Fig. 6C). Inspecting associated tissue regions in the H&E images showed characteristics of early DAD manifested by shedding of ACs, capillary leakage, lymphocytic/immune infiltration, and yet preserved alveoli structure (Fig. 6D and E, T1 and T2). After the shared section, the trajectories diverged into two paths. The first path (Trajectory A) included T3a and T4a and progressed through cells mostly from tissues 3 and 4 enriched in Signature 5 “VEC with MMs infiltration” ending in Signature 6 “VEC with NK cell infiltration”, which was almost exclusively from tissue 3 (Fig. 6A, B and D). We observed a persistent decrease in ACs but an increase in VECs, NK cells and B cells (Fig. 6C). The H&E staining of tissue regions for T3a and T4a showed hyaline membrane, fibrin, lymphocytic infiltration, and collapsed alveoli and capillary structure (Fig. 6E, T3a and T4a, and S1). These features are consistent with a more advanced DAD. The close association of this path with tissues 3 and 4 suggested that they encompassed similar DAD-related tissue patterns and immune niches. Indeed, the postmortem report showed similar findings for these two patients including bilateral pulmonary consolidation [9].

**Figure 6:**
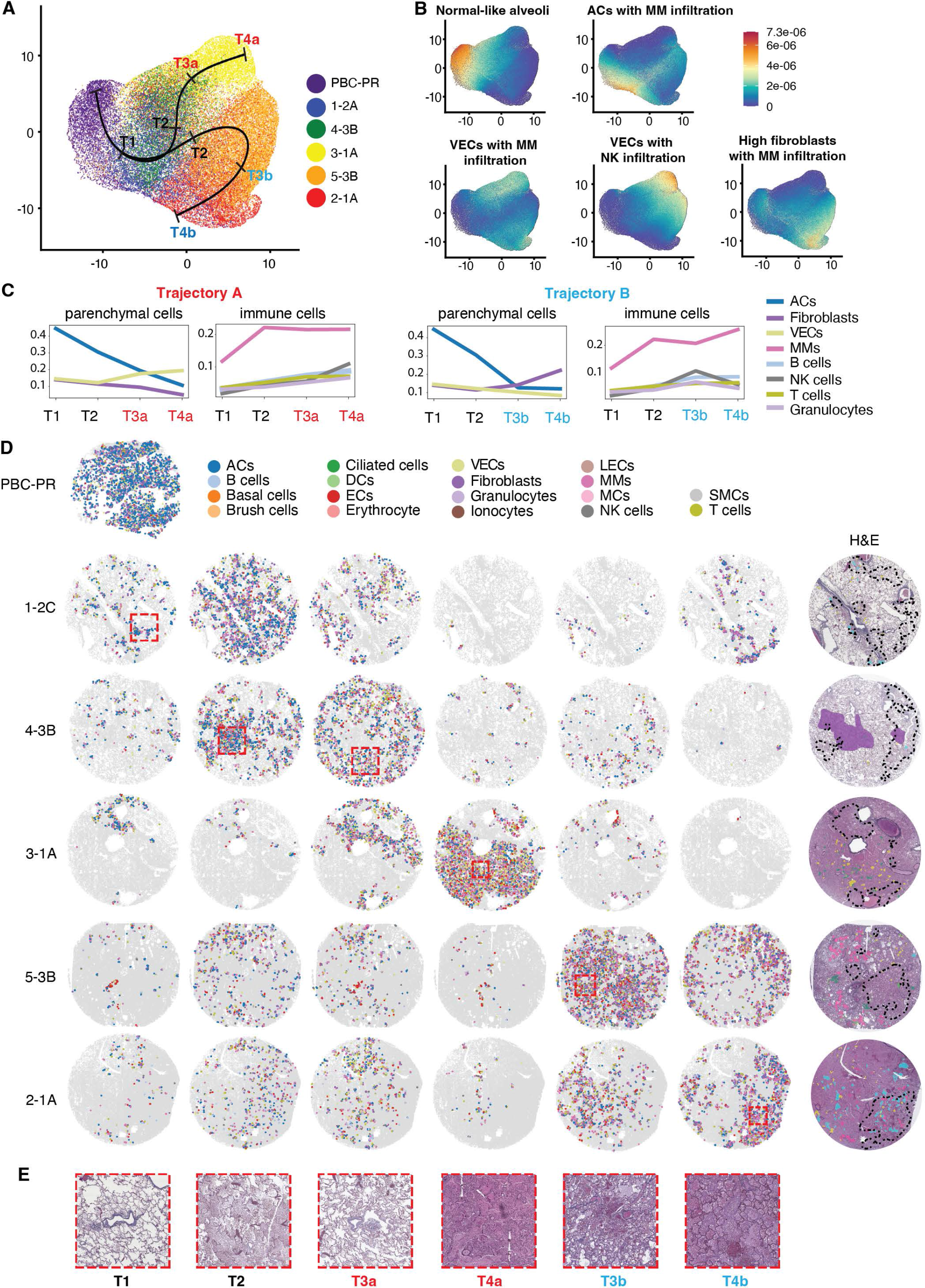
Unique cell composition signatures define two separate pathological trajectories. A. The UMAP projection of the Signatures 1 (for non-COVID-19 tissue reference) and Signatures 4-7 recapitulated the severity of pathological progression identified in figure 5C. Two trajectories were derived from the pseudo-progressions of cells defined by these signatures. The trajectories showed a common shared onset T1 and T2 which traverses through Signature 1 “normal-like-alveoli” and Signature 4 “ACs with MMs infiltration” in T1 and then those enriched by Signature 4 and Signature 5 “VECs with MMs infiltration” in T2. The trajectory then diverged into two paths T3a and T4a, and T3b and T4b. Path/Trajectory A defined by T3a and T4a progressed through cells mostly from tissues 3 and 4 enriched in Signature 5 “VEC with MMs infiltration” ending in Signature 6 “VEC with NK cell infiltration”, which was almost exclusively from tissue 3. Path/Trajectory B defined by T3b and T4b are enriched by Signature 6 in T3b and Signature 7 “high fibroblasts with MMs infiltration” in T4b. **B.** UMAP projections of the five signatures colored based on their individual signature loadings. **C.** Line plots illustrating the cell type percentages in band T1, T2, T3(a, b), and T4(a, b). **D.** Spatial visualization of the cells by cell type in each band is shown along with an identified representative ROI (red box with dotted lines) for each COVID-19 tissue. Column 7 illustrates the H&E morphology image of the COVID-19 tissues with black dotted lines highlighting the high infection regions. **E.** Zoomed-in images of the ROIs selected are shown. A close examination revealed characteristics of early DAD in T1 and T2, hyaline membrane, fibrin, lymphocytic infiltration, collapsed alveoli and capillary structure in T3a and T4a, and edema, increased fibrin, and organizing pneumonia (OP) in T3b and T4b suggesting a more advanced stage of DAD.

The second path (Trajectory B) included two stages, i.e., T3b and T4b, which are enriched by Signature 6 in T3b and Signature 7 “high fibroblasts with MMs infiltration” in T4b highlighted by increased fibroblasts and immune infiltration of especially MMs (Fig. 6A, B, and C). Their associated tissue regions were mostly in tissues 1, 2, and 5 (Fig. 6D), suggesting potentially similar DAD tissue patterns and underlying immune niches in these tissues. The postmortem findings indeed revealed massive pulmonary emboli that were the main cause of death for cases 1 and 2 [9]. Close examination of these regions in the H&E staining showed edema, increased fibrin, and OP (Fig. 6D, E, T3b and T4b, and S1), which are pathological signatures of DAD. There was a striking association between regions in tissues 1, 2 and 5 in T4b with the identified OP, fibrin and hyaline membrane regions and a significant positive correlation between the progression of Trajectory B and cell density (0.504, Spearman correlation, *p-value* < 0.05) suggesting a more advanced stage of T4b than T3b with tissue 1 manifested in a more localized while tissues 2 and 5 in a more systematic manner. These results demonstrated a high degree of inter and intra-tissue heterogeneity in DAD-associated patterns. While tissue 1 contained regions affiliated with both trajectories, tissues 2 and 5 were mostly associated with the trajectory featured by high fibroblasts and their reorganization, and tissues 3 and 4 were with the trajectory featured by increased VEC and immune infiltration.

Next, we investigated the genes whose expressions in a cell type were correlated with the trajectories. We found 53 and 67 genes significantly correlated with Trajectory A and B in at least one cell type, respectively (Spearman correlation, *p-value*<0.05, permutation test; Table S6). In both trajectories, we obtained a negative correlation for ACs marker SFTPC in ACs (Fig. S26A, S26B, Table S6, and Table S7), suggesting a decrease in its expression, which echoes the reductions in ACs in both trajectories (Fig. 6C). We also found a positive correlation of NCAM1 in NK cells with Trajectory A and positive correlations of COL1A1 in Fibroblasts and C1QB in MMs with Trajectory B (Fig. S26A, S26B, Table S6, and Table S7), all of which were consistent with the observed changes of cell type compositions and DAD pathologies along the trajectories (Fig. 6C).

To gain further insight into potential ligand-receptor interactions that underlie the immune patterns associated with the trajectories, we correlated the ligand-receptor co-expressions with the trajectories. We found that CD40LG-CD40, CD80-CD28 and CXCL10-CXCR3 had the highest positive correlations with Trajectory A in all key cell types including especially VECs, ACs and Fibroblasts (Fig. S26C) while CD40LG-CD40, CSF3-CSF3R and CXCL10-CXCR3 were the top positive correlated pairs with Trajectory B in ACs, Fibroblasts, VECs and MMs (Fig. S26D). Thus, these ligand-receptor pairs might contribute to the chemoattraction of the immune cells as a result of SARS-CoV-2 infection.

### IL6-STAT3 and TGF-β-SMAD2/3 pathways mediate COVID-19 lung fibrosis and organizing pneumonia

Because Trajectory B progressed into tissue regions that manifested extensive OP, we investigated the spatial patterns and molecular signatures associated with OP, which was mostly observed in tissues 1 and 2 and to a less extent in tissue 5, which also contained fibrin and hyaline membrane (Fig. 7A and S1). Analysis of cell compositions in regions with OP revealed an overall increase in cell density, and obvious decreases in cell numbers of ACs and VECs but an increase in cell numbers of fibroblasts compared to other regions in the same tissues and the non-COVID-19 tissue (Fig. 7B, C and D). As in the whole tissues, we observed downregulations of AGER, CAV1, SFTPA1, SFTPC, and A2M, and upregulation of IGKC in OP regions compared to the normal tissue (Fig. 7E). The changes of AGER, SFTPA1, SFTPC, and A2M were primarily observed in fibroblasts, MMs and VECs with downregulation of A2M also being observed in ACs (Fig. 7F, G and H). The most striking observation is the upregulation of genes encoding for collagen type I alpha 1 and 2 chains (COL1A1 and COL1A2) in OP in both tissues compared to the normal tissue or to regions without OP in the same COVID-19 lung tissues (Figure 7E and G). Furthermore, both COL1A1 and COL1A2 were upregulated in fibroblasts in OP regions compared to fibroblasts from either normal tissue or non-OP regions from the same tissue (Fig. 7G, H and I). COL1A1 and COL1A2 form the triple helix of a fibril-forming collagen, which is essential for lung homeostasis. Dysregulation of COL1A1 and COL1A2 leads to lung inflammation and fibrosis [67, 68]. Both COL1A1 and COL1A2 were mainly expressed in fibroblasts (Fig. 7G and 7H), whose cell numbers were doubled in OP regions in both tissues compared to non-OP regions (Fig. 7D). Interestingly, the expression levels of COL1A1 and COL1A2 were highly heterogeneous in the individual cells in OP regions with extremely higher levels in 10-20% of the fibroblasts, which were absent in fibroblasts in non-OP regions or any other types of cells in the same COVID-19 tissues (Fig. 7J). COL1A1 and COL1A2 were not expressed in high levels in any types of cells in normal lung tissue (Fig. 7J). These results indicated that COVID-19 OP regions had severe dysregulation of fibroblasts with upregulation of COL1A1 and COL1A2, and abnormal ACs and VECs as a result of downregulation of AGER, SFTPA1, and SFTPC.

**Figure 7:**
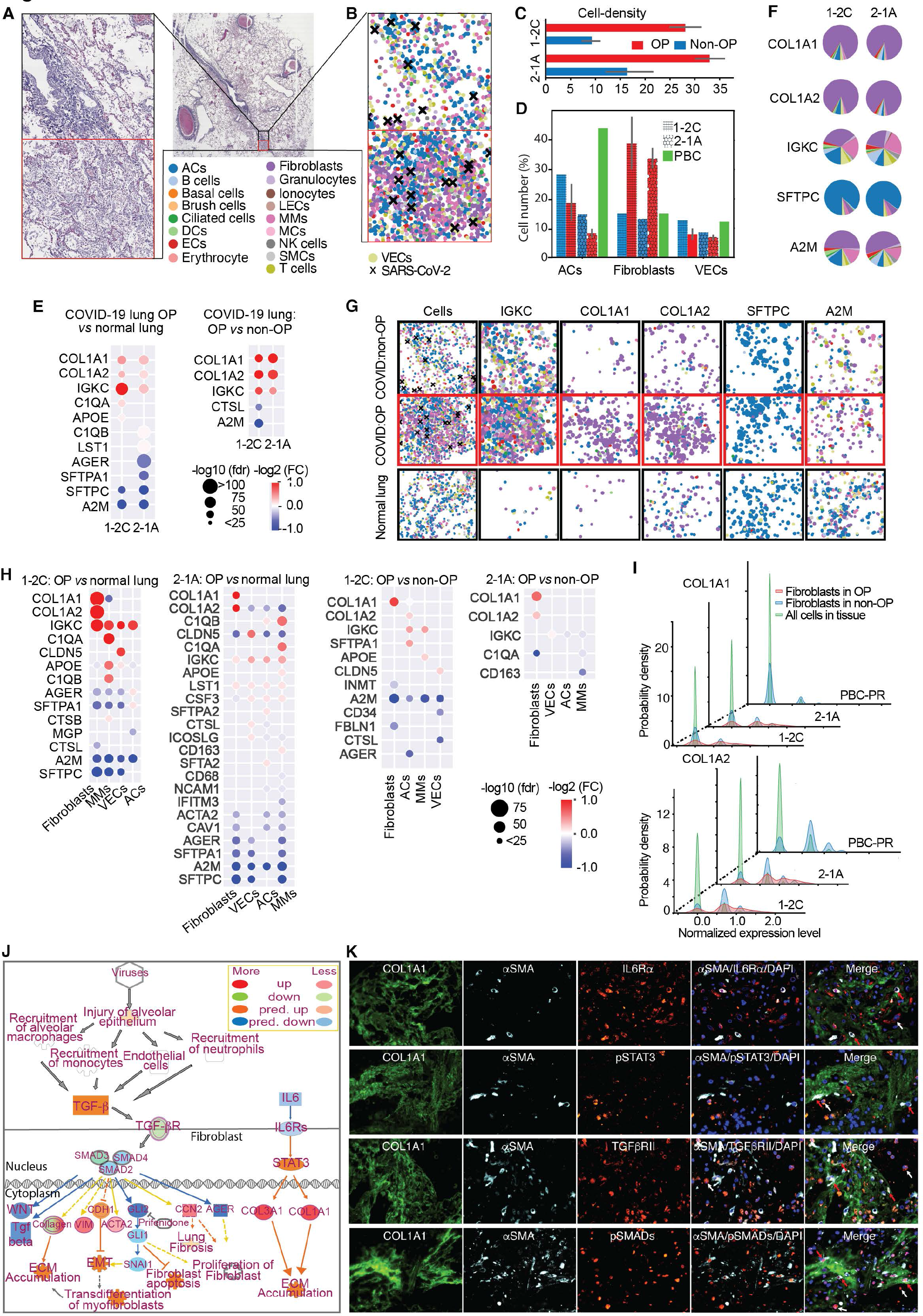
IL6-STAT3 and TGF-β-SMAD2/3 pathways mediate COVID-19 lung fibrosis and organizing pneumonia. A. Zoomed in OP region (bottom, red color border) and other non-OP regions (top, black color border) from the H&E image of COVID-19 tissue 1-2C are illustrated. **B.** Identified cells colored based on the cell types revealed differences in cell composition signatures between OP and other regions. **C.** Bar plot showing an increase in cell density in OP regions compared to other regions. **D.** Bar plot showing the percentage of cells per cell type in COVID-19 and non-COVID-19 tissues. There is a decrease in ACs and VECs, and an increase in fibroblasts in the OP regions of the COVID-19 tissue. An obvious increase in ACs is seen in the non-COVID- 19 sample. **E.** Differential gene expressions between the identified cells in the OP regions of the COVID-19 tissue sample with cells from the non-COVID-19 tissue and cells from the other non-OP regions of the same COVID-19 tissue is illustrated. We observed an upregulation of genes encoding for collagen type I alpha 1 and 2 chains (COL1A1 and COL1A2) in OP in both tissues compared to the normal tissue or to regions without OP in the same COVID-19 lung tissues. **F.** Pie chart illustrating the percentages of cell types in which the selected genes are expressed in tissues 1 and 2. **G.** Spatial visualization of differentially expressed genes from figure 7E. The size of the dots indicates the expression level, and the color indicates the cell type of the cell where the gene is expressed. **H.** Differential gene expressions between the identified cells in the OP regions of the COVID-19 tissue with cells from the non-COVID-19 tissue as well as non-OP regions from the same COVID-19 tissue based on their cell types. Both COL1A1 and COL1A2 were upregulated in fibroblasts in OP regions compared to fibroblasts from either normal tissue or non-OP regions from the same tissue. **I.** The density plot of the COL1A1 and COL1A2 expressions in OP and other non-OP regions in COVID-19 tissue samples 1-2C, 2-1A, and non-COVID-19 tissue is illustrated. **J.** Examination of the networks associated with the Pulmonary Fibrosis Idiopathic Signaling Pathways reported by the Ingenuity Pathway Analysis is illustrated. Both IL6-STAT3 and TGF-β-SMAD2/3 pathways mediate the expression of COL1A1 and COL1A2. **K.** Multi-color IFA staining revealed that cells with a high COL1A1 expression level also had high levels of IL6Rα, p-STAT3, TGF-βR2 and SMAD2/3, confirming the activation of these pathways and their potential roles in the upregulation of COL1A1.

To further confirm that increased expressions of COL1A1 and COL1A2 were hallmarks of COVID-19 OP, we examined their expressions in different cell types in single-cell RNA-seq (scRNA-seq) data from COVID-19 lung tissues [4]. Consistent with our observation, we found that COL1A1 and COL1A2 expressed mainly in fibroblasts and also showed varying expression levels (Fig. S27A). Specifically, they showed a higher expression level in a subset of fibroblasts enriched with myofibroblasts (p < 0.001, Gene Set Enrichment Analysis (GSEA): Fig. S27A and B). Myofibroblasts are differentiated fibroblasts, whose dysregulation may lead to idiopathic pulmonary fibrosis (IPF) including COVID-19 lung fibrosis, a lung disease that exhibits OP [69–71]. To provide a linkage between the OP regions and IPF, we examined the scRNA-seq data from lung tissue patients with IPF and nonfibrotic controls [72]. Like in the COVID-19 data, we observed a subpopulation of fibroblasts with high expressions of COL1A1 and COL1A2 (Fig. S27C) and they were also enriched in myofibroblasts (p-values<0.001, GSEA; Fig. S27D). We further examined the differentially expressed genes in IPF fibroblast cells with high COL1A1 and COL1A2 expressions *vs.* fibroblast cells with low COL1A1 and COL1A2 expressions or nonfibrotic cells, respectively (Fig. S27E and S27F). The two lists of significant differentially expressed genes (DEGs) were similar (p- values<0.001, GSEA; Fig. S27G), suggesting that the subpopulation with low COL1A1 and COL1A2 expressions could serve as normal control. As a result, we compared the fibroblasts with a high *vs.* low COL1A1 and COL1A2 expressions in the COVID-19 scRNA-seq data and obtained the DEGs (Fig. S27H). These DEGs are enriched by two DEG lists in IPF fibroblasts with high COL1A1 and COL1A2 expressions (p< 0.001, GSEA; Fig. S27I and S27J), suggesting that these cells likely had similar molecular characteristics as those in IPF. Indeed, functional enrichment analysis using the Ingenuity Pathway Analysis reported “Pulmonary Fibrosis Idiopathic Signaling Pathways” as the 2^nd^ top-ranked pathway (Fig. S27K). Several other top enriched pathways related to extracellular matrix remodeling were also identified. Examination of the networks of these pathways identified IL6-STAT3 and TGF-β-SMAD2/3 pathways that could directly regulate the expressions of COL1A1 and COL1A2 (Fig. 7K). Multi- color indirect immunofluorescence antibody assay (IFA) staining indeed revealed that cells with a high COL1A1 expression level also had high levels of IL6 receptor-α (IL6Rα), phospho-STAT3 (p-STAT3), TGF-βR2 and phospho-SMAD2/3 (p-SMAD2/3), confirming the activation of these pathways and their potential roles in the upregulation of COL1A1 (Fig. 7L). Both IL6 and TGF-β are highly induced, and implicated in fibrosis and lung injury [73, 74] as well as upregulation of COL1A1 and other extracellular matrix proteins in COVID-19 lung tissues [75, 76]. These results demonstrated the important roles of IL6 and TGF-β in the induction of severe COVID-19 lung OP and fibrosis.

## Discussion

Using *in-situ* sequencing and our spatial single-cell analysis pipeline, we have painted an atlas of cellular and molecular signatures of lung tissues from severe COVID-19 cases. The atlas describes the cellular heterogeneity, organization, and interactions associated with inflammation, damage, and immune responses in COVID- 19 tissues. It provides molecular and cellular insights into the mechanisms underlying SARS-CoV-2 infection and the pathological manifestations in COVID-19 lungs.

We detected a total of 10,414,863 transcripts in five COVID-19 and one non-COVID-19 lung tissues. Spatial single-cell transcriptomics analysis at this scale for COVID-19 and other pathological conditions is still very limited. We have overcome considerable challenges and developed a pipeline of robust, scalable, interpretable computational tools and visualization methods for targeted and exploratory analyses emphasizing on spatial discovery. The first critical component of this pipeline is a transcript-based cell segmentation strategy that integrates cell nuclei in the matching DAPI staining identified by *CellPose* with spatially localized reads using *Baysor* and iteratively segments reads into cells of potentially different sizes [10, 77]. Using this strategy, we segmented 93% of total transcripts into 1,719,459 cells including 186,659 to 470,294 cells for different tissues. A customized filter was further applied to retain only a subset of high-quality cells.

Cell typing of these filtered cells based on marker genes showed that SARS-CoV-2 infected all the 18 identified cell types, a result consistent with those of our previous study [9] and others [4]. However, only a small fraction of these cells (<5%) are infected. These results agree with those of genome-wide studies that detected small numbers of viral reads in blood, lung, and nasopharyngeal samples from severe COVID-19 patients [4, 78–81]. Despite the detected low infection rates, SARS-CoV-2 inflicts similar effects on the infected and uninfected cells across the tissues as shown by differential expression analysis of genes in infected or uninfected cells in COVID-19 tissues against cells in the non-COVID-19 lung tissue (Fig. 2A). Thus, indirect effects such as those induced by the immune response, inflammation, cytokines, and complement activation might play a significant role on lung pathology in COVID-19 patients. In fact, we have observed vast pulmonary microthrombi, thrombosis, and immune infiltrations in these COVID-19 lung tissues (Fig. S1). The major parenchymal cells including ACs, fibroblasts, and VECs have the highest numbers of infected cells. The COVID-19 lung tissues have reduced numbers of ACs and SMCs, but an increased number of ECs accompanied by vast immune infiltrations of innate immune cells including MMs, granulocytes, and NK cells, and adaptive immune cells including T and B cells.

Despite these global and consistent impacts of SARS-CoV-2 infection on the lungs, there are extensive spatial heterogeneities in addition to varying cell type percentages across the tissues. To reveal spatial dynamics of cellular features presented in considerable spatial discontinuity due to sparse measurements, we adopted a strategy to compute local feature distributions in the neighboring region of the cell with a fixed radius or cell number. This approach assesses the spatial gene expression maps, local infection rates, cell densities, and co-expression maps of ligand- receptor pairs. To further characterize spatial patterns of an individual feature or joint pattern relationship between two features, we extensively applied the Moran’s I statistics, a method widely adopted in geoscience but scarcely in spatial transcriptomics analysis. Using these novel strategies, we identified regions with high infection rates, which are found in regions with high cell densities expressing higher levels of SARS- CoV-2 entry-related factors including ACE2, FURIN, TMPRSS2, and NRP1. Thus, SARS-CoV-2 may preferentially infect cells expressing these factors and the infected cells could also aggregate to these high-density regions following infection. Importantly, these regions are mapped to OP (tissues 1 and 2) or regions with fibrin, hyaline membrane, and edema (tissues 3, 4, and 5), and manifest cellular and molecular patterns resembling tissue damage and wound healing including infiltration of immune cells and pattern of fibrosis. We have found that OP harbors high densities of fibroblasts expressing high levels of COL1A1 and COL1A2, a phenomenon common in fibrosis and wound healing, which are unique to OP regions unseen in other regions of COVID-19 lung tissues or in the non-COVID-19 tissue. We have identified similar cell populations enriched by myofibroblasts in 10X scRNA-seq data from COVID-19 lung samples and IPF samples, suggesting that SARS-CoV-2 might preferentially infect myofibroblasts or reprogram other cell type(s) into myofibroblasts. Pathway enrichment analysis revealed the enrichment of multiple pathways related to IPF signaling and extracellular matrix remodeling including IL6-STAT3 and TGF-b-SMAD2/3 pathways. Indeed, we have detected high levels of IL6Ra, p-STAT3, TGF-bRII, and p-SMAD2/3 in cells expressing myofibroblast markers in OP regions. These results suggest that SARS-CoV-2 infection might induce aggregation of fibroblasts, and IL6 and TGF-b to promote wound healing in OP regions.

To define spatial patterns associated with COVID-19 lung pathology, we performed the NCTC analysis, or the niche analysis, commonly used for probing interactions of cells such as in immune microenvironments. However, the COVID-19 tissues contain complex pathology manifestations resulting in highly heterogenous cellular organizations and thus disparate NCTC patterns. To tackle this challenge, we applied SNMF, owing to its scalability and interpretability, to NCTCs of all cells in six tissues to reveal potentially latent features underlying these seemingly less organized NCTCs. We indeed identified 7 latent signatures of spatial cell type compositions that define normal lung structures and SARS-CoV-2 infection-induced immune niches. While many existing studies have revealed the global immune landscape of SARS-CoV-2 infection [1, 2, 5], few report spatial signatures of immune niches. We showed that these niches are mostly tissue-independent, but COVID-19 tissues presented heterogeneous distributions of these niches: while tissues 1, 2, 4, and 5 contain multiple types of niches, tissue 3 has predominately a single type. We further drew the connections between these niches and different stages of DAD. Such a linkage prompted us to apply the trajectory analysis to NCTCs of these signatures. While the trajectory analysis is most commonly applied to gene expressions to predict a pseudo-progression of the cells, here, this innovative use of trajectory analysis defined the relative severities of damages among COVID-19 tissues and constructed two different pathological routes of progression in COVID-19 patients. Both routes start with enriched “ACs with MM infiltration” and “VECs with MMs infiltration” niches but Route A is marked by increased numbers of NK cells and granulocytes, likely reflecting complications of microbial infections, while Route B is characterized by increased HIHD and OP, marked by increased fibrosis. Both routes are also correlated with multiple cytokine-cytokine receptor pairs such as CD40LG-CD40 and CXCL10-CXCR3 that are likely to mediate the chemoattraction of the immune cells (Fig. S26C and S26D). By mapping these routes to individual cells, we further revealed considerable inter and intra-tissue heterogeneity in the inferred progression of COVID-19 pathology with tissue 3 and 4 associated with Route A, tissue 2 and 5 with Route B, and tissue 1 with both routes.

In summary, we have presented an atlas of spatial patterns of different cellular features that characterize SARS-CoV-2 infection, its induced immune infiltrations, inflammation, and damages in severe COVID-19 lungs, in addition to changes in gene expression in multiple cell types, including immune cells and lung parenchymal cells. These results provide insights into the spatial mechanisms at both molecular and cellular levels, which characterize the development of ARDS in COVID-19 patients.

Overall, this study demonstrates the power of spatial single-cell transcriptomics and enables spatial computational analyses in the study of COVID-19 lung pathology. The developed innovative methods can benefit spatial single-cell analyses for other healthy and diseased conditions.

## Methods

### Lung tissue samples

COVID-19 lung tissues were collected from 5 adults with fatal SARS-CoV-2 infection by the Autopsy Service of the Department of Pathology, Molecular and Cell-Based Medicine at the Icahn School of Medicine at Mount Sinai, and non-COVID-19 lung tissue was obtained from Pitt Biospecimen Core [9].

### Study approval

Specimens obtained at autopsy do not meet the definition of a living individual per Federal Regulations 45 CFR 46.102, and as such, research using specimens obtained at autopsy does not meet the requirements for Institutional Review Board (IRB) review or oversight under the Icahn School of Medicine Program for the Protection of Human Subjects. The University of Pittsburgh IRB determined that the study is not research involving human subjects as defined by DHHS and FDA regulations and waived of ethical oversight (STUDY20050085).

### In-situ sequencing (ISS)

HS Library Preparation kit (P/N 1110-02, CARTANA AB, part of 10x Genomics) was used to prepare the library according to the manufacturer’s instruction with minor modification [82]. Four μm FFPE tissue sections were baked for 1 hour at 60°C and deparaffinized by incubating in xylene twice for 7 minutes (min) each. The sections were then rehydrated by incubating in 100% ethanol (EtOH) for 5 min, followed by 70% EtOH for 5 min, and nuclease-free deionized distilled water (SH30538.02, HyClone) for 2 min. Sections were then permeabilized by incubating in citrate buffer pH 6.0 (C9999, Sigma Aldrich) for 3 hours at 95°C. Chimeric padlock probes (from 1 custom panel (Table S1) and 4 predesigned panels: hImmune I1A, hImmune I3D, hLung L1C, hLung L2E, CARTANA AB) directly targeting RNA and containing an anchor sequence as well as a gene-specific barcode were hybridized overnight at 37°C, then ligated overnight at 30°C. Quality control of the library preparation was performed by applying anchor probes which labeled by Cy5 to simultaneously detect all rolling circle amplification products from all genes in the panel. All incubations were performed in SecureSeal™ hybridization chambers (621502, Grace Biolabs). Slow Fade Antifade Mountant (S36936, ThermoFisher) was used for mounting. All samples passed the quality control and were sent to CARTANA AB (part of 10x Genomics) for *in-situ* barcode sequencing, imaging, and data processing. Briefly, the fluorescent signals for quality control were stripped. Adapter probe pool 1 and a sequencing pool containing 4 different fluorescent labels were hybridized to the *in-situ* libraries to detect gene-specific barcodes. The scanning was fulfilled by using epifluorescence microscopy, and raw data consisting of 20x magnification images from 5 fluorescent channels (DAPI, Alexa Fluor® 488, Cy3, Cy5 and Alexa Fluor® 750) and individual z-stacks, were flattened to 2D using maximum intensity projection with a Nikon Ti2 Microscope (software NIS elements) utilizing a Zyla 4.2 camera. In total, 6 sequencing cycles were achieved for full decoding of all designed genes. After image processing, which includes image stitching, background filtering, and a sub-pixel object registration algorithm, true signals were scored based on signal intensities from individual multicolor images. The results were summarized in a CSV file and gene plots were generated using MATLAB. Each ISS spot provided a unique fluorescent barcode identifying the targeted RNA of a gene marker. The experiment generated a map of gene expressions of selected genes recorded on the natural morphology of the tissue at a single-cell level. The metadata generated from the images were further analyzed to obtain data containing the reads and their 2D (X, Y) positions of each gene, and the DAPI images. TissuuMaps [83, 84] was used to visualize spatial read locations of multiple genes against DAPI or histology images.

### Hematoxylin-eosin staining (H&E) and whole slide scanning

H&E was carried out with the same slides subjected to *in-situ* sequencing using Hematoxylin & Eosin Stain Kit (H-3502, Vector Laboratories) according to the manufacturer’s instructions. The slides were then scanned with VS200 Slide Scanner (Olympus).

### Indirect immunofluorescence antibody assay (IFA)

IFA was carried out as previously described [9]. Primary antibodies included CoraLite® Plus 647-conjugated smooth muscle actin (1:200, Proteintech, CL647- 67735), CoraLite® Plus 488-conjugated Collagen Type I (1:100, Proteintech, CL488- 67288), IL6Rα (1:500, Proteintech, 23457-1-AP), p-STAT3 (Tyr705) (1 to 20, Invitrogen, 710093), p-SMAD2/3 (p-SMAD2-S465/467 and p-SMAD3-S423/425, 1:500, ABclonal, AP0548), and TGF-βRII (1:10, R&D Systems, AF-241-NA). Secondary antibodies included Goat anti-Rabbit IgG (H+L) Highly Cross-Adsorbed Secondary Antibody, Alexa Fluor™ Plus 555 (1:400, Invitrogen, A32732), Donkey anti-Mouse IgG (H+L) Highly Cross-Adsorbed Secondary Antibody, Alexa Fluor™ 568 (1:400, Invitrogen, A10037).

### Cell segmentation

Reads in a tissue sample were segmented into corresponding cells using the Baysor cell segmentation algorithm [10]. Baysor applies a Markov Random Field model to identify spatial clustering of reads from the same cell. It can perform cell segmentation based on read coordinates alone but can also incorporate nuclear information from DAPI staining. We first utilized the DAPI-stained images of the tissues and performed the nuclei segmentation using the *CellPose* anatomical segmentation algorithm with the default setting of parameters [77]. Segmented nuclei masks with locations were provided to Baysor to define its scale and standard deviation parameters. Baysor was able to segment 71%- 91% of reads after the first run for all samples/tissues. However, we found that among the unsegmented, the so-called noisy background reads determined by Baysor, there is a considerable portion that could still be assigned to cells visually. To address this issue, we applied Baysor again to these noisy reads from the first Baysor run. We optimized the scale and standard deviation parameter for the second Baysor run to maximize reads assignment to cells. Including the second run significantly improved the percentages of the segmented reads to 88%-95% for all samples/tissues (Fig. S3, S4 and Table S2). We implemented only two runs because the remaining unsegmented reads were mostly noisy background reads.

### Normalization and filtering

Segmented gene reads assigned to cells were then filtered using multiple criteria.

First, we removed cells that did not contain reads from any marker genes. Second, we removed cells with total read counts <5 or less than 4 genes with reads. Of the remaining cells, read counts for each gene within a cell were tallied and gene expressions were normalized using the *scTransform* algorithm [11] to further remove biases due to technical variability. The normalized expressions of 220 genes were used for subsequent analysis.

### Cell typing

We determined the cell type of each cell using an in-house curated list of marker genes for 18 cell types (Table S1). For each cell, we calculated the average expression of marker genes for each of the 18 cell types and then assigned the cell type with the highest average expression to this cell. We further annotated infected cells as those with at least one SARS-CoV-2 read. We used UMAP to visualize the expression pattern of cells associated with the identified cell types in a lower dimension by using the visualization pipeline in the Seurat package [85].

### Image registration

The DAPI images of tissues that were used as a prior for cell segmentation had translational and rotational differences with their corresponding H&E stained images. For single-cell resolution datasets, small spatial geometric changes in the available raw morphology images could have dramatic differences in downstream analysis for studying the pathology signatures using annotated regions in the H&E image. To mitigate these spatial geometric differences, we registered the DAPI and H&E images by maximizing the phase correlation between both images. Here, we selected the DAPI image as a fixed reference because we used DAPI images as references for reads mapping and cell segmentation. To register both images, the H&E stained image is transformed by changing the scale, rotation, and translation parameters and moved across the DAPI image to improve the phase correlation of both 2D images. Once a peak correlation value was found, the optimization algorithm returned the 2D geometric transformations required to warp the H&E-stained image and registered it to the corresponding DAPI image. We applied the same image registration algorithm to all six samples used in this study. Pathology annotations were carried out on the registered H&E images.

### Cell density analysis

The abundance of cells relative to the tissue context could reveal biology related to cell proliferation, cell damage, as well as other pathologies related to infection. In our spatial analysis pipeline (Fig. 1B), we defined cell density based on the number of cells within a neighborhood of a fixed radius of 200 units (200 x 0.32 μm/unit = 64 um) centering a cell, covering an area of 0.013 mm^2^. By calculating the number of cells within the fixed neighborhoods of each cell, we summarized the global cell density patterns across the tissue. We further presented the cell density as a contour plot by fitting a 2D kernel density estimator (KDE) on the cell density feature (Fig. 3).

### Neighboring cell type composition analysis

Local cell type distributions can reveal tissue structures and different COVID-19 pathology. We assessed the local cell type distribution by computing the neighboring cell type composition (NCTC) of each cell. NCTC defined a cellular neighborhood of 200 cells and counts the number of different cell types in the neighborhood to create a neighborhood count vector for each cell. This neighborhood count vector was normalized to obtain the percentages of cell types. NCTC summarized the distributions of each cell type in the vicinity of a cell and was further used to analyze the spatial correlation of different cell types in a region-of-interest (ROI), study spatial gene expression patterns of local neighborhoods, identify local hotspots of individual cell types, and more. Furthermore, the neighborhood composition of individual cell types is spatially mapped by painting the neighborhood count/percentage of the selected cell type with a continuous “rainbow” color scheme where deeper violet to deeper reds indicates a range of low-to-high neighborhood cell composition (Fig. S10-S15).

### Spatial gene expression map

SSCT captures gene expression of single cells in the context of intact tissue structures. In our study, we investigated the spatial organization of cells in tissue and their gene expression by studying their spatial gene expression maps (SGM). For each gene, its SGM is defined as the sum of the expression of that gene in its 200 nearest neighboring cells. Thus, SGM represents a spatial gradient by summarizing the regions with high or low gene expression patterns. We then painted the SGM using the continuous rainbow colormap similar to the NCTC analysis, generating the spatial gene expression maps illustrated in Fig. 2B, 2C, and 4F. By comparing the SGM with available pathology annotations, pathology signatures with relevant expression patterns can be localized to specific spatial regions in the tissue. Additionally, novel pathology signatures or ROIs can be discovered using the proposed SGM analysis.

### Local infection rate analysis

We used the local infection rate analysis to study the spatial distribution of SARS-Cov-2 infection. It was similar to the NCTC analysis and computed the percentage of infected cells in a fixed physical area of 1.286 mm^2^ or a radius of 2,000 units (640 μm) around each cell across the tissue. These spatial rates can be further visualized using the same rainbow color scheme as NCTC (Fig. 3A).

### Ligand-receptor coexpression map

Ligand-receptor interactions among cells in a defined location can define spatial patterns of immune microenvironments due to SARS-CoV-2 infection. To infer the interactions, we examine the spatial co-expression between a ligand-receptor pair.

Inspired by Moran’s I spatial cross-correlation, we computed the spatial coexpression between ligand *x* and receptor *y* for cell *i* as

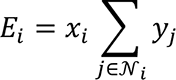

where *x*_*i*_ = 1 (*y*_*j*_ = 1) if ligand *x* (receptor *y*) expressed in cell *i* (*j*) and 0, otherwise and 𝒩_*i*_ defines a set of neighboring cells of cell *i*, which are cells within a radius of 1,000 units (320 μm) or a fixed physical area of 0.322 mm^2^ around cell *i*. Then, we visualized the coexpression in a tissue using a fixed monochrome color scheme (Fig 3F).

### Moran’s I analysis of spatial patterns

Spatial global autocorrelation of one variable or more variables can be summarized using Moran’s I score. However, calculating the global spatial autocorrelation using Moran’s I across the whole tissue assumes homogeneity of the studied variable and yields only one statistic that summarizes the complete spatial pattern across the tissue disregarding their difference over space. Since our SARS-CoV-2 infected tissue samples show spatial heterogeneity, in this study, we used Local Indicators of Spatial Association (LISA) [86] to evaluate the spatial autocorrelation and the statistical significance of a study variable in each location using Local Moran’s I.

Consider *x*_*i*_ as, e.g., the neighboring infection rate of a cell at location *i*, the univariate spatial autocorrelation can be found as the degree of linear association between *x*_*i*_ and a weighted average of the neighboring cells *x*_*j*_, based on a spatial weight *W*_*ij*_between cells at location *i* and *j*. Thus, formally, Moran’s I for each location *i* is given by

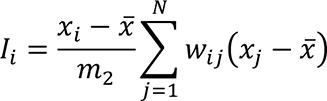

where *N* defines the number of cells, *x* defines the mean of all the cells, and *m*_2_ is given by

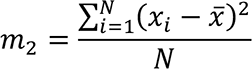

*I*_*i*_ ranges from -1 to 1 with -1 indicating highly negative and +1 indicating highly positive spatial autocorrelation. Such formulation can be used to identify spatial regions with high or low autocorrelation. For example, in Fig. 3B, we identified regions with high and low infection rates in each tissue.

Under the same formulation, bivariate spatial autocorrelation can be used to assess spatial cross-correlation of a feature *x*_*i*_, e.g., the neighboring infection rate of a cell at location *i* and another feature *y*_*j*_, e.g., the cell density of a neighboring cell *j* as

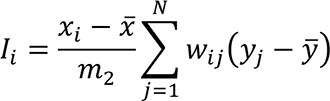

where *I*_!_ defines the degree of linear association between the neighboring infection rate of a cell at location *i* and a weighted average of the cell density of the neighboring cells. Bivariate spatial associations can uncover cross-correlations between any such variable *x* at location *i* and another variable *y* at neighboring locations by ignoring correlations between *x* and *y* for cases where *i* = *j*. In our study, the bivariate LISA method is used to calculate the Local Moran’s I to identify HIHD regions described in Fig. 3E.

### Sparse Non-negative matrix factorization (SNMF) of neighboring cell type compositions

SNMF was applied to extract interpretable, tissue-specific, and tissue-independent signatures from NCTC vectors from all tissue samples [63]. Let ***A*** ∈ (**0,1**)^*N×M*^ represent the NCTC matrix of *N* cell types and *M* cells and it is factored as

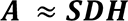

where ***S*** ∈ (***0,1***)^*N×K*^ is the matrix of *K* NCTC signatures with each column denoting a signature, ***H*** ∈ (0,1)^*K×M*^ is the signature loading matrix with *m*th column denoting the contributions of each signature to the NCTC vector of cell *m*, and ***D*** ∈ ℝ^*K×K*^ is a scaling matrix.

R package *RcppML* was used for the SNMF analysis [63], which implemented an alternating least squares (ALS) algorithm to minimize the mean squared error (MSE) between ***A*** and ***SDH***. *L*_1_ regularization was introduced to promote a compact, sparse signature loading.

Determining the best number of signatures *K* is important for uncovering meaningful signatures. While an underestimated number could miss critical signatures, an overestimated number would include many noisy signatures. To this end, we evaluated different maximum numbers of factors from 4 to 12 and examined the meaning regarding its cell type composition and spatial distributions of loadings of each signature and chose *K* = 7 for the SNMF analysis.

### Trajectory analysis

The goal of trajectory analysis is to infer a progression or ‘pseudotime’ of infection severity and associated tissue damages based on NCTC patterns in all tissues. We applied the trajectory analysis to uncover the pseudotime of 1) infection severity among tissues and 2) tissue damage defined by immune microenvironments. For 1), we used loadings of Signature 1 “Normal-like Alveoli” and Signature 3 “Broad

Immune Infiltration” obtained from SNMF and defined tissue samples as clusters. For 2), we used the loadings of Signature 1 and four immune microenvironment-related signatures, *i.e.*, Signature 4-7, and performed Louvain clustering to define the clusters. The R package *Slingshot* was used to infer the trajectory [64]. The loadings, clusters, and the signatures or the 2-D UMAP matrix were fed into *Slingshot* to obtain so-called ‘pseudotime’ scores for each cell. For both cases, the cluster enriched by cells from the non-infected tissue (PBC-PR) cells was set as the origin.

The pseudotime scores of the cells in each trajectory can correlate with gene expression to uncover genes that correlate with tissue damage. Additionally, ligand-receptor coexpressions that correlate with each trajectory could inform potential ligand-receptor interactions that govern immune responses. To this end, we computed the Spearman correlation of the pseudotime score with the expression of each gene per cell type in each trajectory. We also computed the correlation of ligand-receptor coexpressions with the pseudotime score of each trajectory. In both cases, to properly assess *p-values*, we performed a permutation test to obtain the empirical null distribution of the correlation coefficients and chose *p-value*<0.5 as the significant level. We then plotted heatmaps to illustrate the cell-type-wise correlations of the genes in each trajectory (Fig. S26A-D).

### Publicly available scRNA-seq datasets

Two scRNA-seq datasets were obtained and processed as follows.

1. COVID-19 scRNA-seq dataset [4]. The dataset (SCP1052, lung.h5ad.gz) was downloaded from the Single Cell Portal (https://singlecell.broadinstitute.org/single_cell). The processed data file lung.h5ad was accessed using the Python package Scanpy (Scanpy 1.9.1). UMAP coordinates for the processed data were accessed from the file upload.scp.X_umap.coords.txt. No follow-up processing was performed on the processed *anndata* object after it was loaded into Scanpy.
2. IPF scRNA-seq dataset [72]. The dataset (GSE135893, GSE135893_ILD_annotated_fullsize.rds.gz) was downloaded from Gene Expression Omnibus (GEO). The Seurat object file was converted into the h5ad file format using the SeuratData package (Seurat 4.3.0, SeuratData 0.2.2). After conversion, the data was loaded in Scanpy (Scanpy 1.9.1) and all cells not originating from an IPF sample or control sample were removed.

### Statistics and reproducibility

Reads-based cell segmentation was performed using *Baysor* (version 0.5.2).

Nuclei segmentation from DAPI images was performed using *CellPose* (version 2.1.1). All other image analyses including imaging registration were performed using MATLAB (version 2022a). Unless otherwise specified, *p*-value < 0.05 was considered significant. The differential expression analysis was performed in R (version 3.6.3) using the Wilcoxon rank-sum test with Bonferroni’s correction of multiple testing as necessary.

The Moran’s I analyses for spatial patterns were performed using GeoDa (version 1.20). Pearson correlation coefficients were calculated using function *pearsonr* in Python Scipy (version 1.9.3). UMAP dimension reduction and visualization were performed using function *DimPlot* in Seurat (version 4.2.0). Trajectory analyses were performed using R package *Slingshot* (version 2.6.0).

## Data Availability

The data supporting findings in this study including all the original spatial single-cell transcriptome data, segmented and filtered reads of all cells, and DAPI and H&E staining images will be deposited to Zenodo. The spatial single-cell transcriptome data will also be deposited in GEO.

## Acknowledgments

We thank members of Drs. Shou-Jiang Gao and Yufei Huang laboratories for technical assistance and discussions. This study was supported by grants from the National Institutes of Health (CA096512 and CA124332 to S.-J. Gao), UPMC Hillman Cancer Center Startup Funds to S.-J. Gao and Y. Huang, and in part by award P30CA047904. This research was also supported in part by the University of Pittsburgh Center for Research Computing, RRID:SCR_022735, through the resources provided. Specifically, this work used the HTC cluster, which is supported by the National Institutes of Health (S10OD028483).

## Author contributions

S.-J. Gao and Y. Huang conceived, designed, supervised and managed the project. W. Meng, S.R. da Silva and L.P. Chen performed the experiments. G.L. Sica and W. Meng performed the pathological examination. A. Das, Z.T. Liu, D.M. Hasib, H. Galloway and Y. Huang developed the computational pipeline and performed the analyses. A. Das, W. Meng, Z.T. Liu, D.M. Hasib, H. Galloway, S.R. da Silva, L.P. Chen, Y.F., K.P. Rivera, M. Flores, Y.-C. Chiu, Y. Huang and S.-J. Gao interpreted the data and participated in the discussions throughout the analysis. A. Paniz-Mondolfi, C. Bryce, Z. Grimes, E.M. Sodillo and C. Cordon-Cardo obtained the COVID-19 samples. Y. Huang and S.-J. Gao wrote the manuscript with input from all the authors. All the authors read, reviewed and approved the manuscript.

## Competing interests

The authors declare no competing interests.

## Data availability

The data supporting findings in this study including all the original spatial single- cell transcriptome data, segmented and filtered reads of all cells, and DAPI and H&E staining images will be deposited to Zenodo. The spatial single-cell transcriptome data will also be deposited in GEO.

## Code availability

All codes will be available at Zenodo.

## Supplementary information

The online version contains supplementary material available at:

Correspondence and requests for materials should be addressed to Shou-Jiang Gao or Yufei Huang.

## Supplemental Information

### Supplemental Tables

**Table S1.**
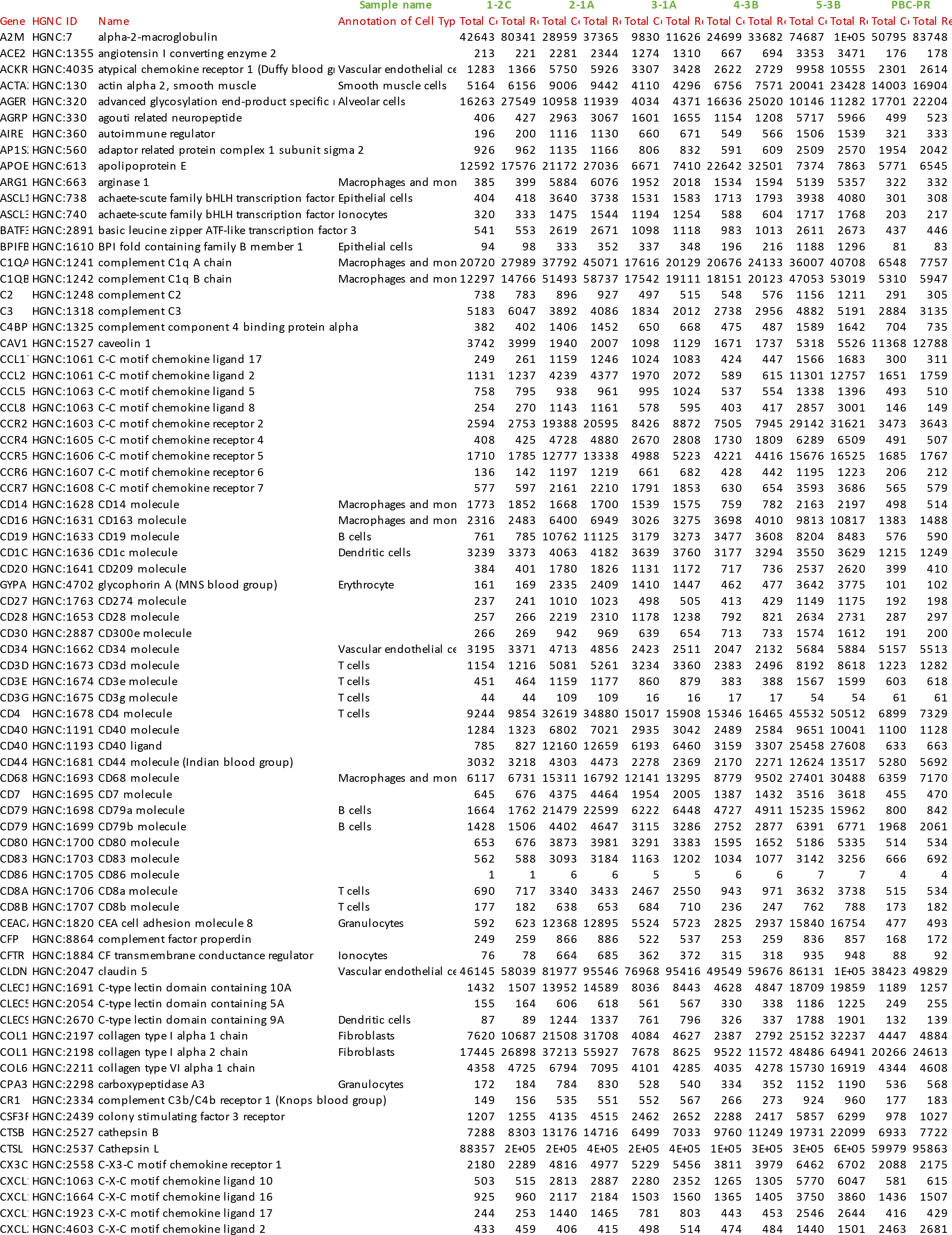
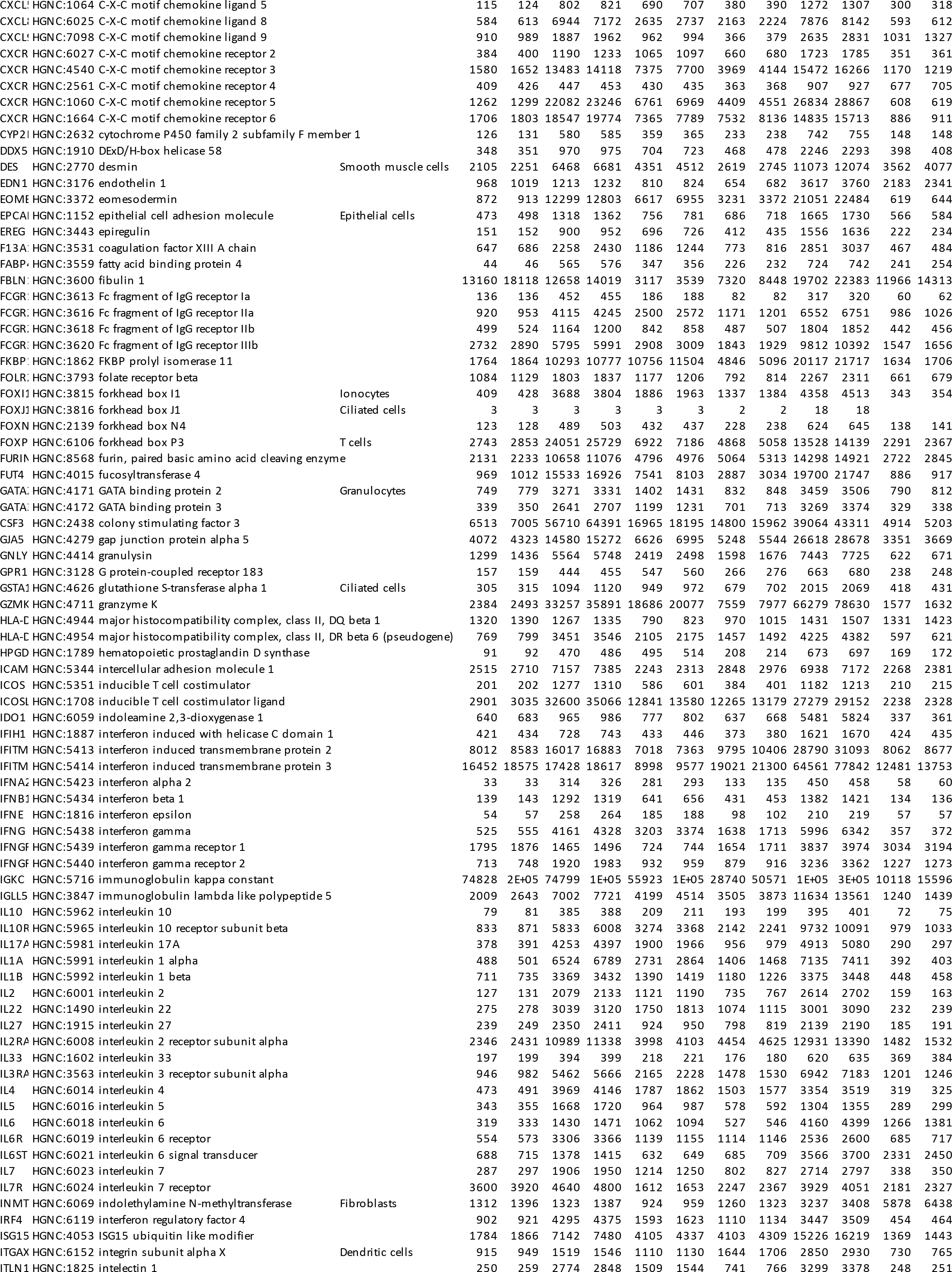
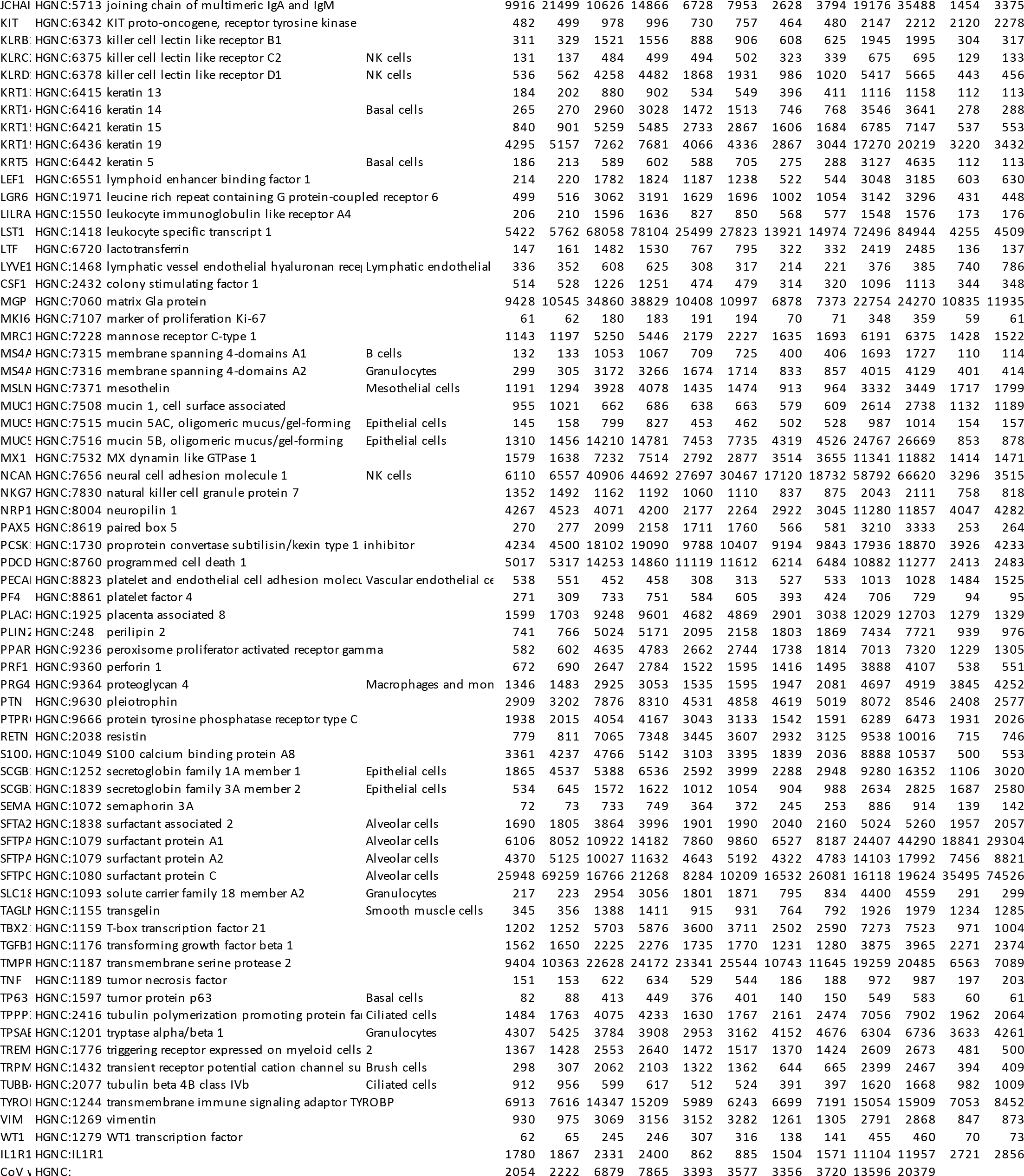
Gene annotation, cell type markers, and total cells expressing individual genes and total reads of the individual genes in each lung tissue after segmentation.

**Table S2.** Summary of cell segmentation results.

| Sample name | 1-2C | 2-1A | 3-1A | 4-3B | 5-3B | PBC-PR | Total |
| --- | --- | --- | --- | --- | --- | --- | --- |
| Total Reads | 1,137,704 | 2,380,636 | 1,441,118 | 1,161,277 | 3,424,675 | 869,453 | 10,414,863 |
| Number of Genes | 222 | 222 | 222 | 222 | 222 | 221 |  |
| Total Segmented Reads | 1,074,491 | 2,224,394 | 1,339,792 | 1,112,148 | 3,231,976 | 773,102 | 9,755,903 |
| Percentages of Segmented Reads (%) | 94.44381 | 93.43696 | 92.96893 | 95.7694 | 94.37322 | 88.9182 | 93 |
| Total Cells | 186,659 | 406,963 | 276,676 | 227,585 | 470,294 | 151,282 | 1,719,459 |
| Total Cells with Cell Type Marker Gene | 137,260 | 317,259 | 193,293 | 163,431 | 369,042 | 126,531 | 1,306,816 |
| Percentages of Cells with Cell Type Marker Gene (%) | 73.53516 | 77.9577 | 69.86258 | 71.81097 | 78.47049 | 83.63916 |  |
| Total Cells after Filtering | 67,701 | 196,221 | 87,701 | 74,843 | 287,832 | 62,674 | 776,972 |
| Average Transcripts per Cell | 5.756469 | 5.465852 | 4.842476 | 4.886758 | 6.87226 | 5.110371 |  |

**Table S3.**
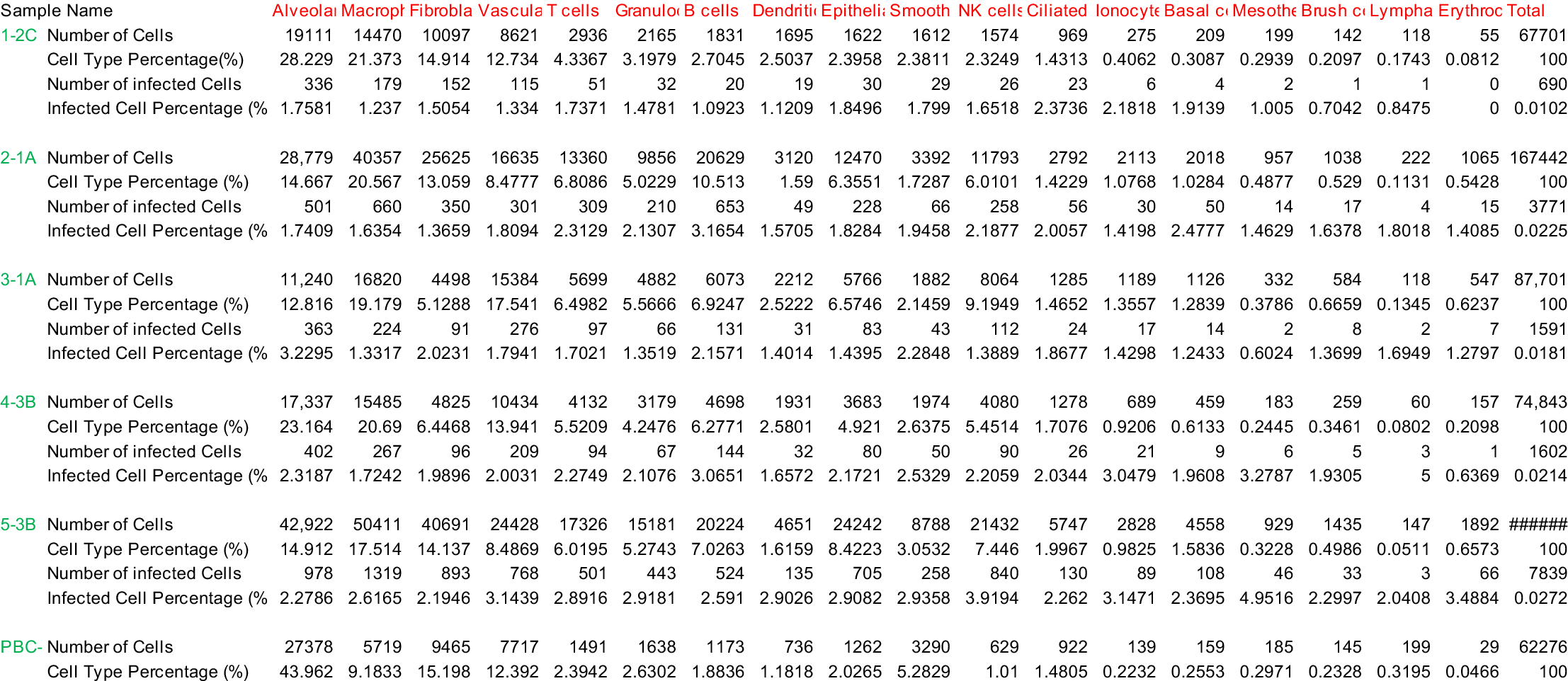
Summary of cell typing results and SARS-CoV-2 infection status.

**Table S4.** Global spatial correlations between local SARS-CoV-2 infection rates and cell densities by global Moran’s I.

| Sample name | 1-2C | 2-1A | 3-1A | 4-3B | 5-3B |
| --- | --- | --- | --- | --- | --- |
| Coefficient (r) | 0.325 | 0.636 | 0.287 | 0.248 | 0.078 |

**Table S5.** Cell composition signatures identified by sparse non-negative matrix factorization (SNMF).

|  | Normal-like<br>Alveoli | Bronchile | Broad<br>immune<br>infiltration | ACs w/ MM<br>infiltration | VECs w/ MM<br>infiltration | VECs w/ NK<br>infiltration | High<br>fibroblasts<br>w/ MM |
| --- | --- | --- | --- | --- | --- | --- | --- |
| n-Alveolar | 0.5643179 | 0.0121138 | 0 | 0.6477053 | 0.0404455 | 0 | 0.0336691 |
| n-B | 0.0177815 | 0 | 0.2864881 | 0.0001489 | 0.0357681 | 0.0298888 | 0.0017179 |
| n-Basal | 0 | 0.1029554 | 0.0025063 | 0 | 0 | 0.0135613 | 0 |
| n-Brush | 0 | 0 | 0.0019199 | 0 | 0 | 0.0032618 | 0 |
| n-Ciliated | 0.0105512 | 0.1403089 | 0.002555 | 0.002187 | 0.0131246 | 0 | 0.0055952 |
| n-Dendritic | 0.0076009 | 0.006494 | 0.0079063 | 0.0128054 | 0.0373424 | 0.0272293 | 0 |
| n-Epithelial | 0 | 0.6141461 | 0.0867002 | 0.0092846 | 0 | 0.1219462 | 0 |
| n-Erythrocyte | 0 | 0.0014696 | 0.0023376 | 0 | 0 | 0.0068894 | 0 |
| n-Fibroblasts | 0.1193691 | 0 | 0 | 0 | 0.031177 | 0 | 0.7342991 |
| n-Granulocytes | 0.0125711 | 0.0494878 | 0.0963014 | 0.0253557 | 0.0136434 | 0.1360229 | 0.0080645 |
| n-Ionocytes | 0 | 0 | 0.0145924 | 0 | 0.0018038 | 0.0202377 | 0 |
| n-Lymphatic | 0 | 0 | 0 | 0 | 0 | 0 | 0 |
| n-Macrophages | 0 | 0.0162048 | 0.2233146 | 0.2728252 | 0.4667314 | 0 | 0.1577208 |
| n-Mesothelial | 0 | 0 | 0 | 0 | 0 | 0 | 0 |
| n-NK | 0 | 0.0066883 | 0.0999384 | 0.0296879 | 0 | 0.3184191 | 0.0179067 |
| n-Smooth | 0.0529362 | 0.0501313 | 0 | 0 | 0.0099377 | 0.0428492 | 0.0410268 |
| n-T | 0.0294544 | 0 | 0.17544 | 0 | 0.0869477 | 0.0040867 | 0 |
| n-Vascular | 0.1854178 | 0 | 0 | 0 | 0.2630784 | 0.2756075 | 0 |

**Table S6.**
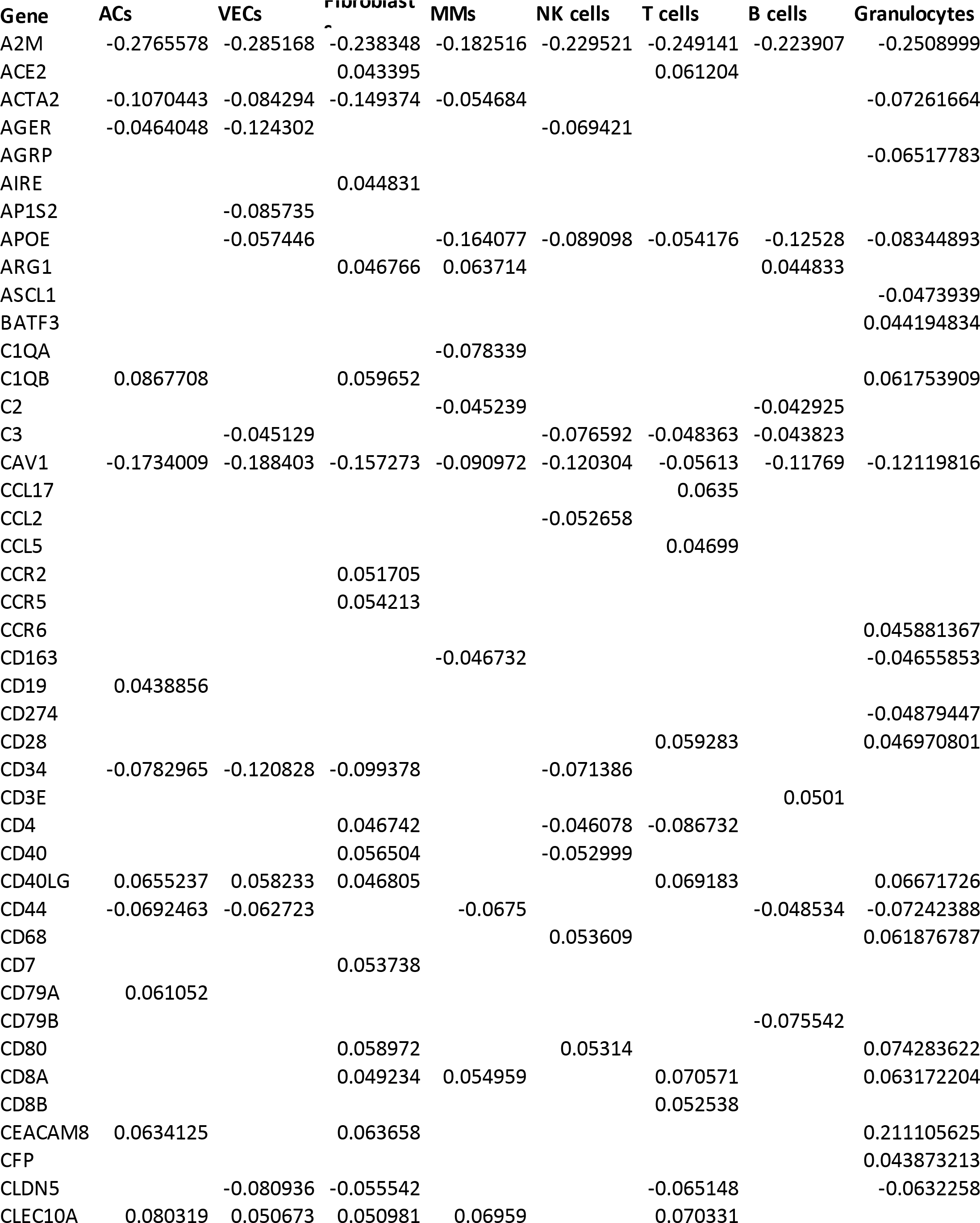
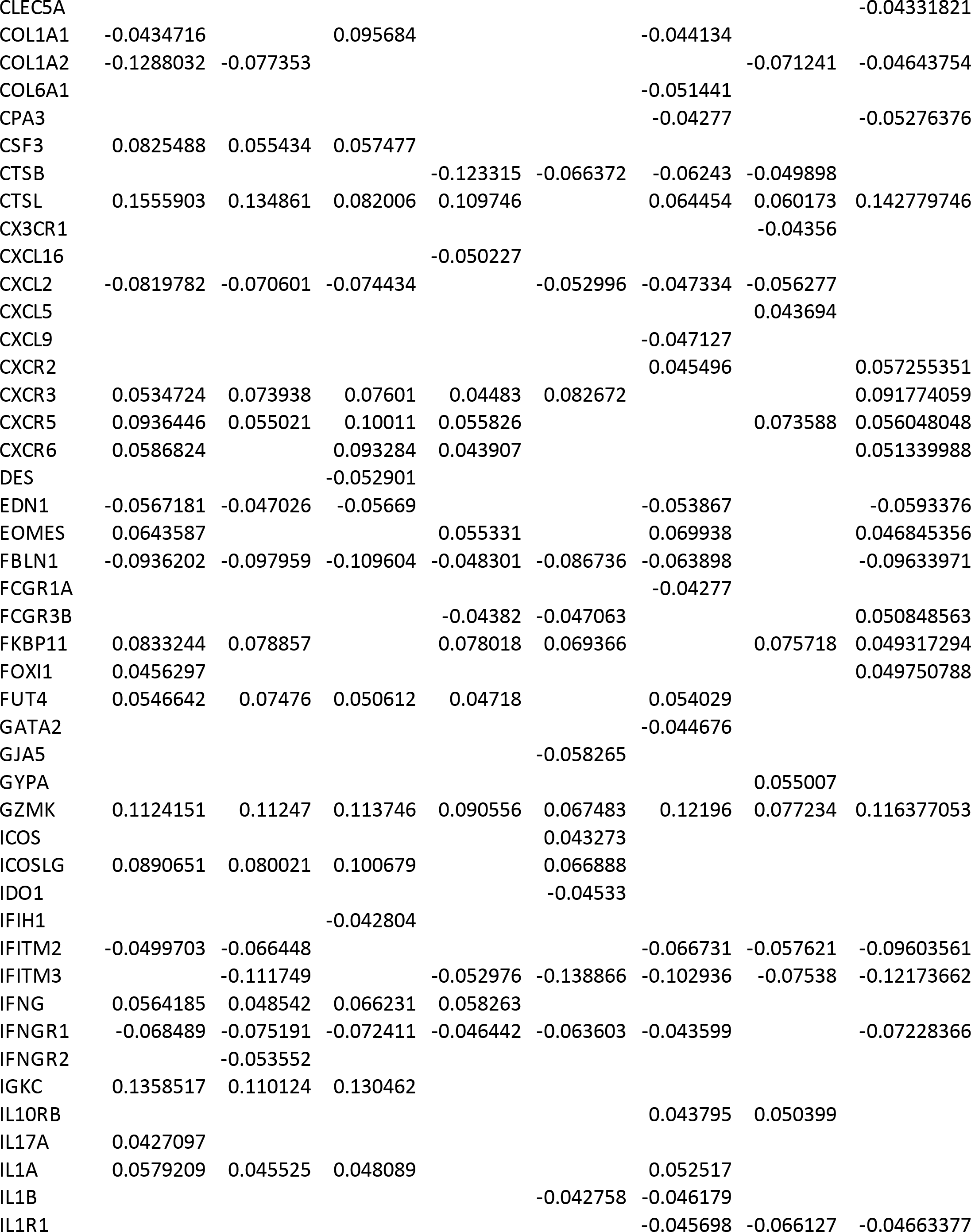
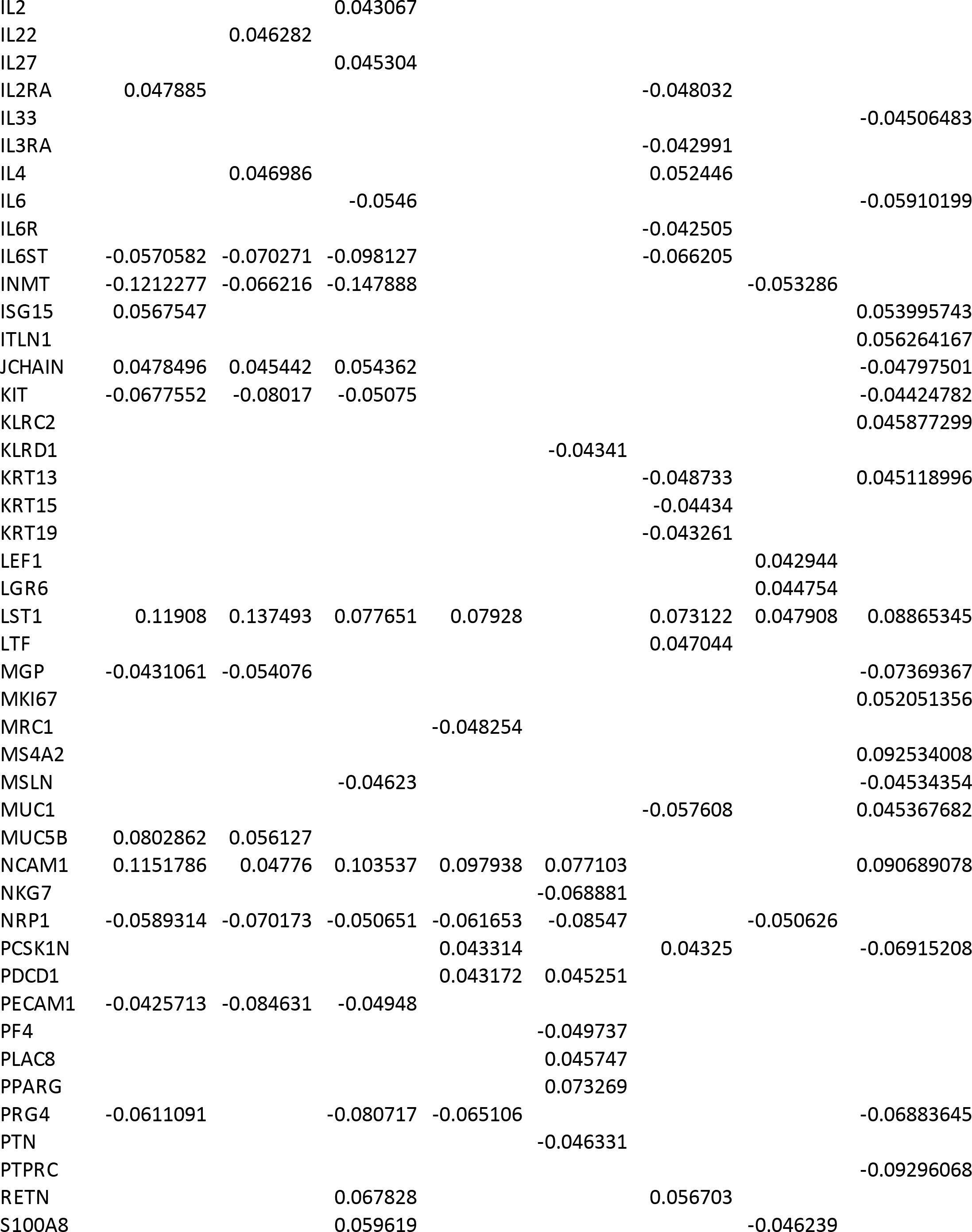
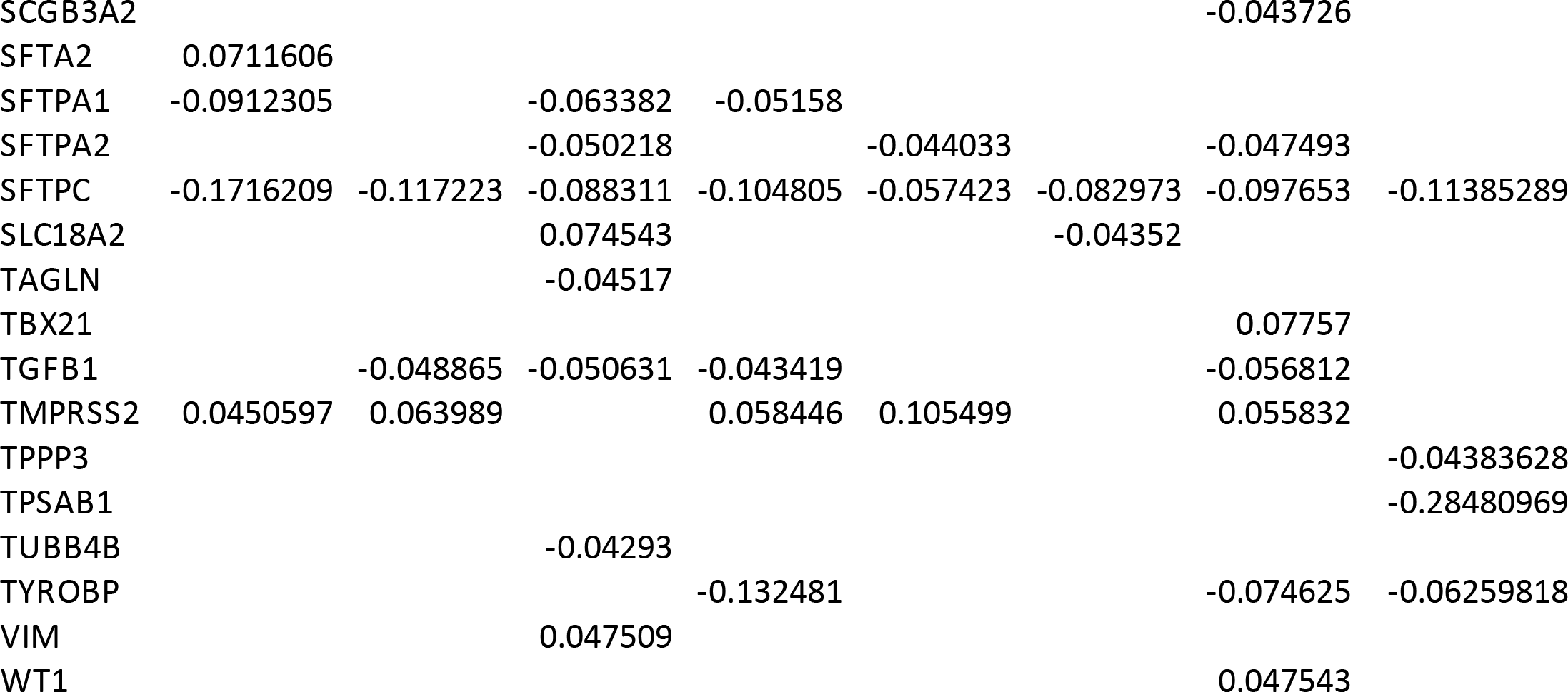
Spearman correlations of pseudotime Trajectory A versus gene expressions.

**Table S7.** Spearman correlations of pseudotime Trajectory B versus gene expressions.

| Gene | ACs | VECs | Fibroblasts | MMs | NK cells | T cells | B cells | Granulocytes |
| --- | --- | --- | --- | --- | --- | --- | --- | --- |
| A2M | -0.23417 | -0.18242 | -0.18695 | -0.11699 | -0.07008 | -0.13915 | -0.11668 | -0.199064005 |
| ACE2 |  |  |  |  |  | 0.047165 |  |  |
| ACKR1 |  | 0.048854 |  |  |  |  | -0.04833 |  |
| ACTA2 | -0.09122 | -0.10498 | -0.09113 |  |  | -0.04457 |  |  |
| AGER | -0.0654 | -0.12614 | -0.04869 |  |  |  |  |  |
| AGRP | 0.049945 |  |  |  |  |  |  |  |
| AP1S2 | -0.04246 | -0.07303 |  |  |  |  | -0.04535 |  |
| APOE |  |  |  | -0.07856 |  | -0.03913 |  |  |
| ARG1 | 0.047952 |  |  |  |  | 0.046863 |  |  |
| ASCL1 |  |  |  |  |  |  |  | -0.037819398 |
| BATF3 |  |  |  |  |  | 0.037658 |  |  |
| C1QA | 0.058691 |  |  |  |  |  |  | 0.09027356 |
| C1QB | 0.117966 | 0.054073 | 0.089102 | 0.096327 |  |  | 0.058879 | 0.125662283 |
| C3 |  |  |  |  |  |  |  | -0.063321455 |
| CAV1 | -0.15927 | -0.18698 | -0.14654 | -0.07419 | -0.07115 | -0.05335 | -0.04429 | -0.103846221 |
| CCL2 |  |  |  |  |  | 0.056462 |  | 0.041291612 |
| CCR2 | 0.06541 | 0.063104 |  | 0.046706 |  | 0.038923 |  | 0.045579992 |
| CCR4 |  | 0.05497 | 0.03879 |  |  |  |  |  |
| CCR5 | 0.06756 |  |  | 0.037975 | 0.051752 |  |  |  |
| CCR6 |  |  |  |  | 0.045262 |  |  |  |
| CD163 |  |  |  |  |  |  |  | 0.037295985 |
| CD19 | 0.060925 | 0.040203 | 0.041826 |  |  |  | 0.044888 |  |
| CD34 | -0.0689 | -0.09669 | -0.06878 |  | -0.0509 |  |  |  |
| CD4 | 0.049011 |  |  |  |  |  |  |  |
| CD40 | 0.039212 |  |  |  |  |  | 0.058188 |  |
| CD40LG | 0.107726 | 0.077691 |  | 0.045761 |  | 0.08453 |  | 0.068746563 |
| CD44 | -0.05332 |  |  |  | 0.052098 |  |  | -0.050139472 |
| CD68 |  |  |  | -0.07668 |  |  |  |  |
| CD7 |  |  |  |  |  | 0.052587 |  |  |
| CD79A | 0.08843 |  | 0.042754 |  |  |  | 0.092262 |  |
| CD79B |  |  |  |  |  |  | -0.12707 |  |
| CD83 |  |  |  |  |  |  |  | 0.047211302 |
| CD8A |  |  |  |  |  |  | 0.038345 |  |
| CEACAM8 | 0.067966 |  |  |  |  |  | 0.055024 | 0.214210161 |
| CFTR |  |  |  |  |  |  | 0.054296 |  |
| CLDN5 | -0.0661 | -0.08001 | -0.08032 |  |  | -0.1043 | -0.05523 | -0.079253269 |
| CLEC10A | 0.068948 | 0.065709 | 0.055969 | 0.057083 |  | 0.074607 |  |  |
| COL1A1 |  |  | 0.238038 |  | 0.050449 |  | 0.063025 |  |
| COL1A2 | -0.08054 |  | 0.074501 |  |  |  | 0.060741 | -0.043885551 |
| CPA3 |  |  |  |  |  | -0.0388 |  | -0.058816528 |
| CSF3 | 0.113034 | 0.10195 | 0.072741 | 0.102288 | 0.071468 |  | 0.088778 | 0.098683964 |
| CTSB |  |  |  | -0.04035 |  |  |  | 0.055171045 |
| CTSL | 0.104649 | 0.07678 | -0.04173 |  |  |  | -0.06617 |  |
| CX3CR1 |  |  |  |  | 0.043399 |  | -0.04299 |  |
| CXCL2 | -0.07253 | -0.0844 | -0.05969 | -0.05841 | -0.06559 | -0.05153 | -0.04755 |  |
| CXCL5 |  |  |  |  |  |  |  | 0.03806835 |
| CXCL9 |  |  |  |  |  | -0.03978 |  |  |
| CXCR3 | 0.069483 | 0.048237 |  |  | 0.037495 |  |  | 0.085780591 |
| CXCR5 | 0.125108 | 0.091302 | 0.039747 | 0.064412 |  |  | 0.053469 | 0.056036 |
| CXCR6 | 0.056839 | 0.055155 | 0.063929 |  |  |  |  | 0.057363906 |
| DES |  |  |  |  |  | 0.055714 | 0.041941 |  |
| EDN1 | -0.04828 |  | -0.04554 |  |  |  |  |  |
| EOMES | 0.084288 | 0.076171 |  | 0.045833 |  |  |  | 0.050475746 |
| FABP4 |  |  |  | -0.03939 |  |  |  |  |
| FBLN1 | -0.07795 | -0.06019 | -0.0729 | -0.04823 |  |  | -0.06675 | -0.099788911 |
| FCGR2A |  | 0.044455 |  |  |  |  |  |  |
| FCGR3B | 0.040975 | 0.037913 |  |  |  |  |  | 0.072317848 |
| FKBP11 | 0.063241 | 0.040355 | 0.056659 |  |  |  |  |  |
| FOLR2 |  |  |  |  |  | -0.04681 |  |  |
| FOXI1 | 0.045565 |  |  |  |  |  |  |  |
| FOXN4 |  |  |  |  |  | 0.043416 | 0.043758 |  |
| FOXP3 | 0.039537 |  | 0.050424 |  | 0.048612 |  | 0.060758 | 0.056544832 |
| FURIN |  |  |  |  |  |  | 0.045908 |  |
| FUT4 | 0.08674 | 0.072236 | 0.040281 | 0.06246 |  | 0.044159 |  |  |
| GATA2 |  |  |  |  |  | -0.04118 |  |  |
| GJA5 | 0.042105 | 0.043181 |  |  |  |  |  |  |
| GNLY | 0.049135 |  |  |  |  |  | 0.039547 |  |
| GPR183 |  |  |  |  |  |  | -0.05082 |  |
| GYPA |  | 0.044047 |  |  |  |  |  |  |
| GZMK | 0.162548 | 0.117448 | 0.066982 | 0.07485 |  | 0.102471 | 0.053519 | 0.121910499 |
| HPGDS |  |  |  |  |  |  |  | -0.058750952 |
| ICAM1 |  |  |  |  |  |  | 0.051938 |  |
| ICOSLG | 0.094865 | 0.108561 | 0.059662 | 0.058779 | 0.06085 | 0.041286 | 0.046443 | 0.048873673 |
| IDO1 |  | 0.039485 |  |  |  | 0.037285 |  |  |
| IFITM3 |  |  |  |  |  |  |  | -0.0401819 |
| IFNB1 |  |  |  |  | 0.064311 |  |  |  |
| IFNG |  | 0.047643 |  |  | -0.03724 |  |  | 0.04148565 |
| IFNGR1 | -0.06606 | -0.06479 | -0.04459 |  | -0.06533 | -0.0605 |  | -0.043583766 |
| IFNGR2 |  | -0.04789 |  |  |  |  |  | 0.041613098 |
| IGKC | 0.200164 | 0.207234 | 0.10137 | 0.08933 | 0.115094 | 0.106026 | 0.061718 | 0.065923338 |
| IGLL5 |  |  |  |  | 0.045037 |  |  |  |
| IL10RB |  |  | 0.04243 |  | 0.042023 | 0.043005 |  |  |
| IL17A | 0.051116 | 0.052451 |  |  |  |  | 0.049536 |  |
| IL1A | 0.061588 | 0.050096 |  |  |  |  |  |  |
| IL1B |  | 0.038184 |  |  |  |  | 0.038756 |  |
| IL22 |  |  |  |  |  |  | 0.041126 | 0.037526937 |
| IL27 | 0.040339 |  |  |  |  |  |  |  |
| IL2RA | 0.073001 |  |  |  | 0.053015 |  | 0.054686 |  |
| IL33 |  |  | -0.03945 |  |  |  | -0.03722 |  |
| IL3RA |  |  |  |  |  |  | 0.043497 |  |
| IL4 | 0.03914 |  |  |  |  |  |  |  |
| IL5 |  |  |  |  |  | 0.065467 | 0.045197 |  |
| IL6 |  |  |  |  |  |  |  | -0.053733035 |
| IL6R |  |  |  |  |  |  | 0.044341 |  |
| IL6ST | -0.05528 |  |  |  |  |  |  |  |
| IL7R |  | -0.04831 |  |  |  | -0.05257 |  |  |
| INMT | -0.12332 | -0.06816 | -0.19792 |  |  |  | -0.06262 |  |
| ISG15 | 0.066814 | 0.038402 | 0.040259 |  |  |  |  |  |
| ITLN1 | 0.044288 |  |  |  |  |  |  | 0.049348458 |
| JCHAIN | 0.082298 | 0.052541 |  |  | 0.063171 | 0.058752 |  |  |
| KIT | -0.05272 | -0.08017 |  |  |  |  |  | -0.053323915 |
| KLRC2 |  |  |  |  | -0.04922 |  | 0.050395 |  |
| KLRD1 |  |  |  |  |  |  | 0.044409 |  |
| KRT13 |  |  |  |  |  |  |  | 0.041034312 |
| KRT14 | 0.040772 |  |  |  |  |  |  |  |
| KRT15 | 0.046743 |  |  |  |  |  |  |  |
| LST1 | 0.176173 | 0.140293 | 0.075228 | 0.097155 | 0.048409 | 0.150042 | 0.066195 | 0.117086543 |
| LTF |  |  |  |  |  | 0.058064 |  |  |
| MGP |  |  |  |  | 0.060665 |  |  | -0.047453011 |
| MRC1 |  |  |  |  | 0.039171 |  |  |  |
| MSLN |  |  |  |  |  |  | -0.04002 | -0.053807806 |
| MUC5AC |  |  |  |  |  |  | -0.04623 |  |
| MUC5B | 0.100919 | 0.037387 | 0.038258 |  |  | 0.037932 |  |  |
| MX1 | 0.037898 |  |  |  |  |  |  |  |
| NCAM1 | 0.129607 | 0.038924 | 0.070591 | 0.060254 | 0.040041 |  |  | 0.054425241 |
| NRP1 | -0.0381 | -0.04668 |  | -0.05298 |  |  |  |  |
| PAX5 |  |  |  |  |  |  |  | 0.053371989 |
| PCSK1N |  |  |  |  |  |  |  | -0.076337283 |
| PDCD1 |  |  | 0.037544 |  |  |  |  |  |
| PECAM1 | -0.04893 | -0.09513 | -0.05137 | -0.04777 | -0.06127 |  |  |  |
| PLAC8 | 0.039513 |  |  |  |  |  |  |  |
| PLIN2 |  | 0.049445 |  |  |  |  |  |  |
| PPARG | 0.037847 |  |  |  |  |  |  |  |
| PRF1 |  |  |  |  |  |  | -0.04373 |  |
| PRG4 | -0.06815 | -0.04201 | -0.07122 | -0.06506 |  |  |  |  |
| PTN |  |  | -0.03789 |  | -0.06056 |  |  |  |
| PTPRC |  |  |  |  |  |  |  | -0.05500837 |
| RETN | 0.066851 | 0.040041 |  |  |  |  |  |  |
| S100A8 | 0.048937 |  |  |  |  |  |  |  |
| SEMA3A |  |  |  |  | 0.059976 |  |  |  |
| SFTA2 | 0.090868 |  |  |  |  |  |  |  |
| SFTPA1 | -0.07088 | -0.05154 |  |  |  |  |  |  |
| SFTPA2 |  |  |  |  |  |  | -0.04342 |  |
| SFTPC | -0.18456 | -0.09881 | -0.07617 | -0.07591 | -0.05003 | -0.06539 | -0.06338 | -0.11238912 |
| SLC18A2 | 0.059352 |  |  |  |  |  |  | 0.053280437 |
| TGFB1 |  | -0.0503 |  |  |  |  |  | 0.042625481 |
| TMPRSS2 |  |  |  |  |  | -0.04518 |  | -0.07145939 |
| TPPP3 |  |  |  |  |  |  |  | -0.038857276 |
| TPSAB1 |  | -0.04459 | -0.04399 |  |  | 0.038693 | -0.04669 | -0.291267486 |
| TYROBP |  |  |  | -0.04596 |  |  |  |  |

### Supplementary Figures

**Supplementary Figure 1:**
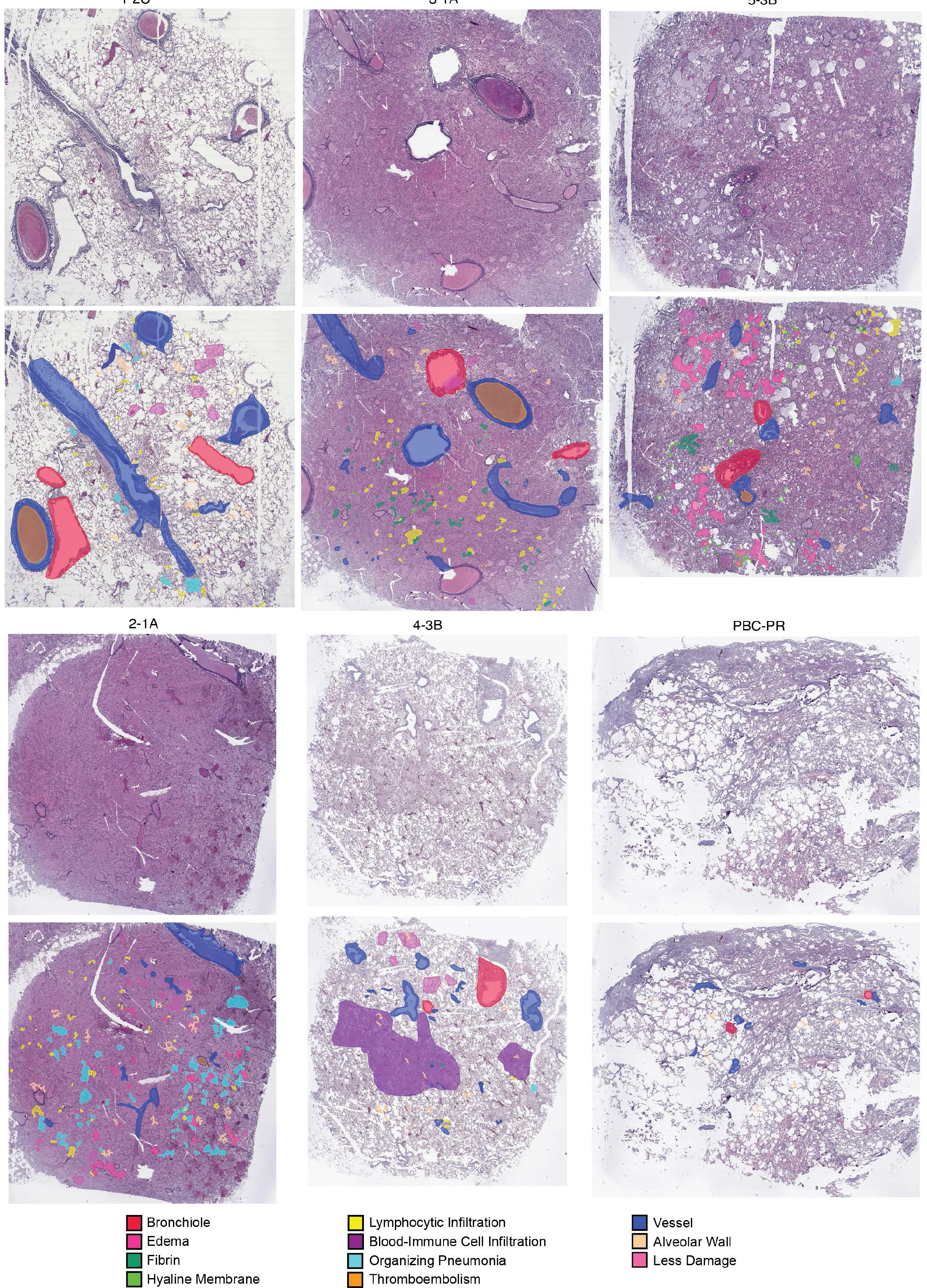
Hematoxylin and eosin (H&E) stained images of the five COVID-19 tissues (1-2C, 2-1A, 3-1A, 4-3B, 5-3B) and the non-COVID-19 tissue (PBC- PR) along with pathology annotations are illustrated. We observe various degrees of diffuse alveolar damage (DAD) in the COVID-19 tissues. Tissues 1 and 2 (1-2C and 2- 1A) had prominent organizing pneumonia (OP) while edema, hyaline membrane, and fibrin clot were prominent in tissues 3, 4, and 5 (3-1A, 4-3B, and 5-3B).

**Supplementary Figure 2:**
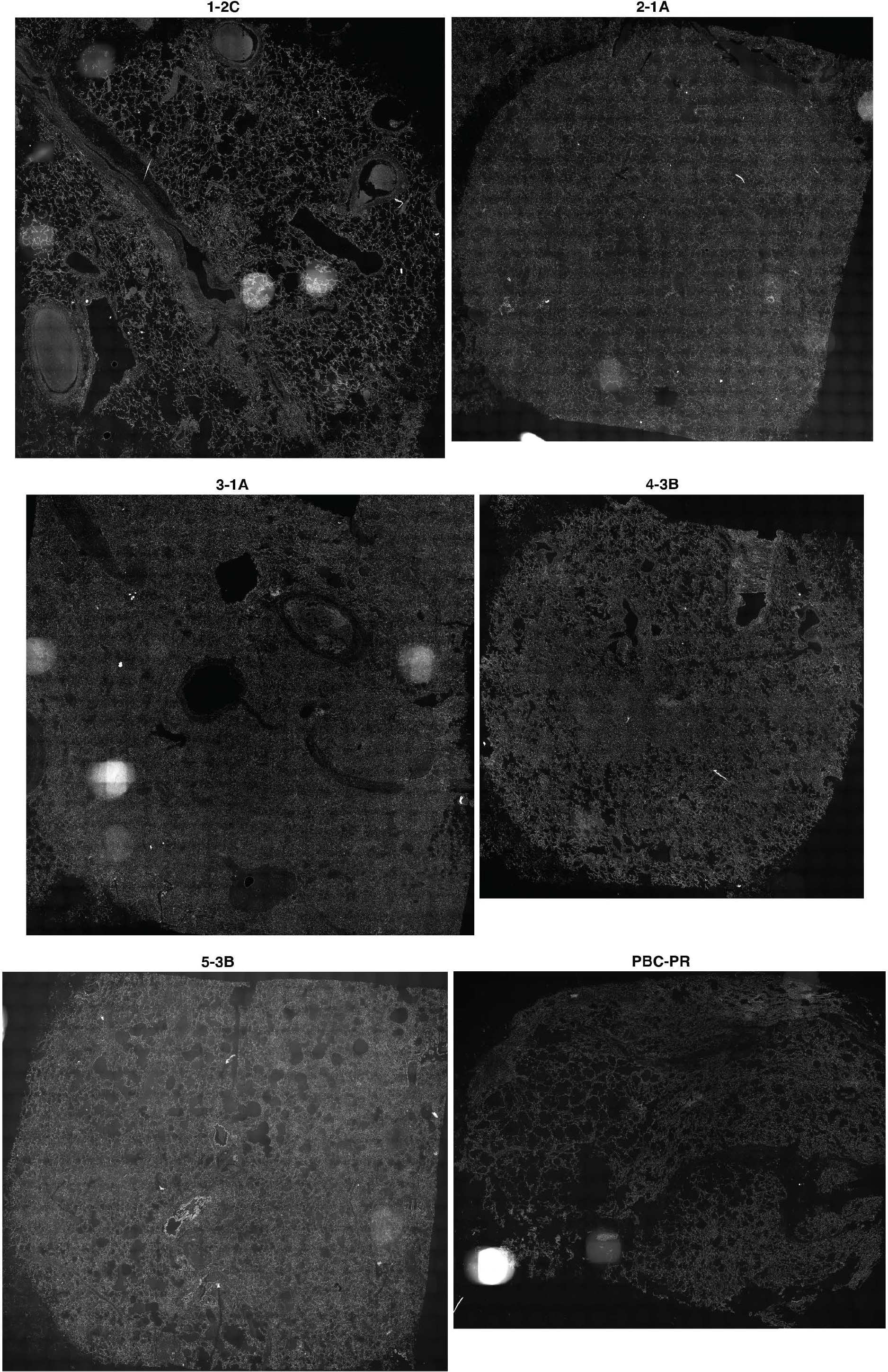
4′,6-diamidino-2-phenylindole (DAPI) staining of all tissues under study revealed the nuclei location. We used the DAPI-stained images to facilitate cell nuclei segmentation using the CellPose algorithm.

**Supplementary Figure 3:**
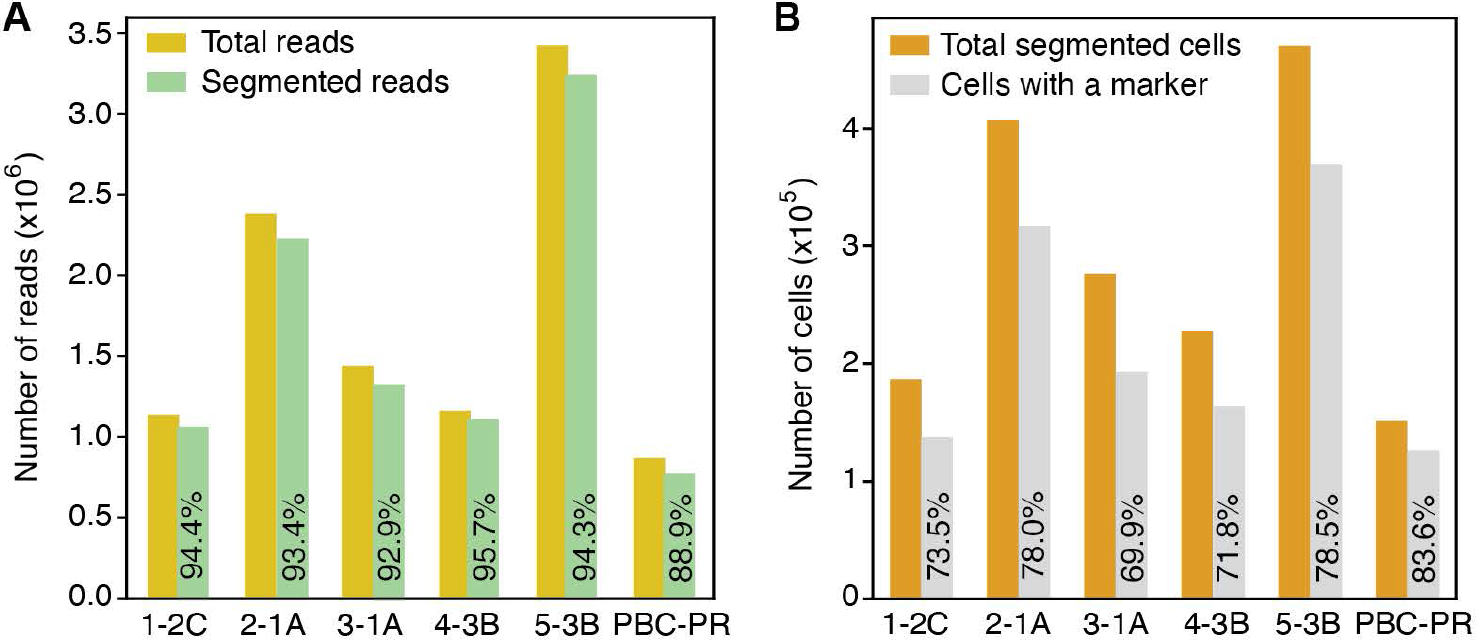
A. Bar plot showing the total number of reads and the reads that are segmented to cells using the Baysor algorithm for all tissues under study. We segmented ∼89%-95% of the reads into cells. **B.** Bar plot showing the total number of segmented cells and the identified cells with a cell-marker read. A total of 1,719,459 cells were identified across the tissues.

**Supplementary Figure 4:**
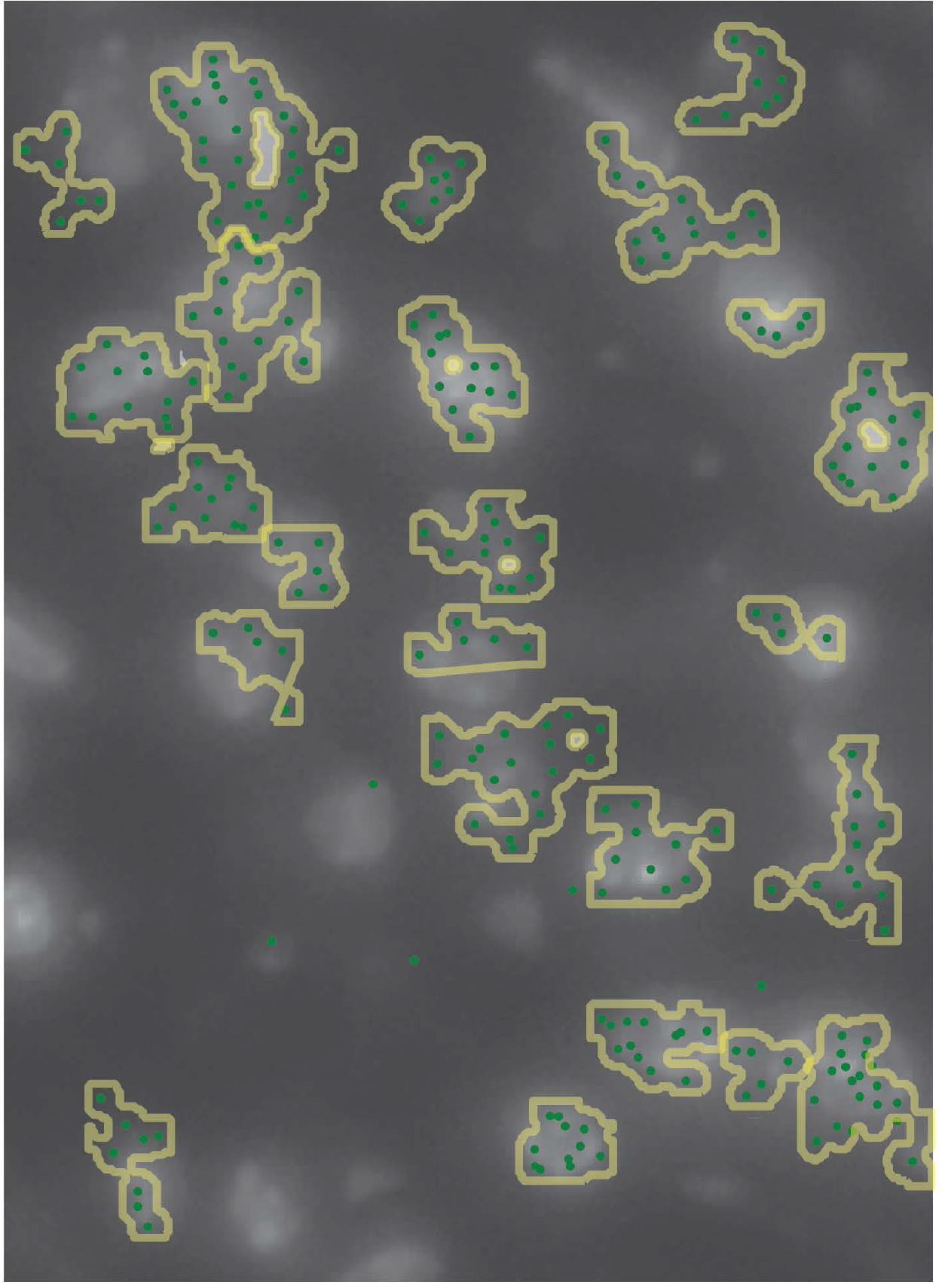
A representative image showing cell boundary polygons of the cells segmented using the Baysor algorithm. Baysor used a binary mask of the DAPI image as a prior to guide the cell segmentation.

**Supplementary Figure 5:**
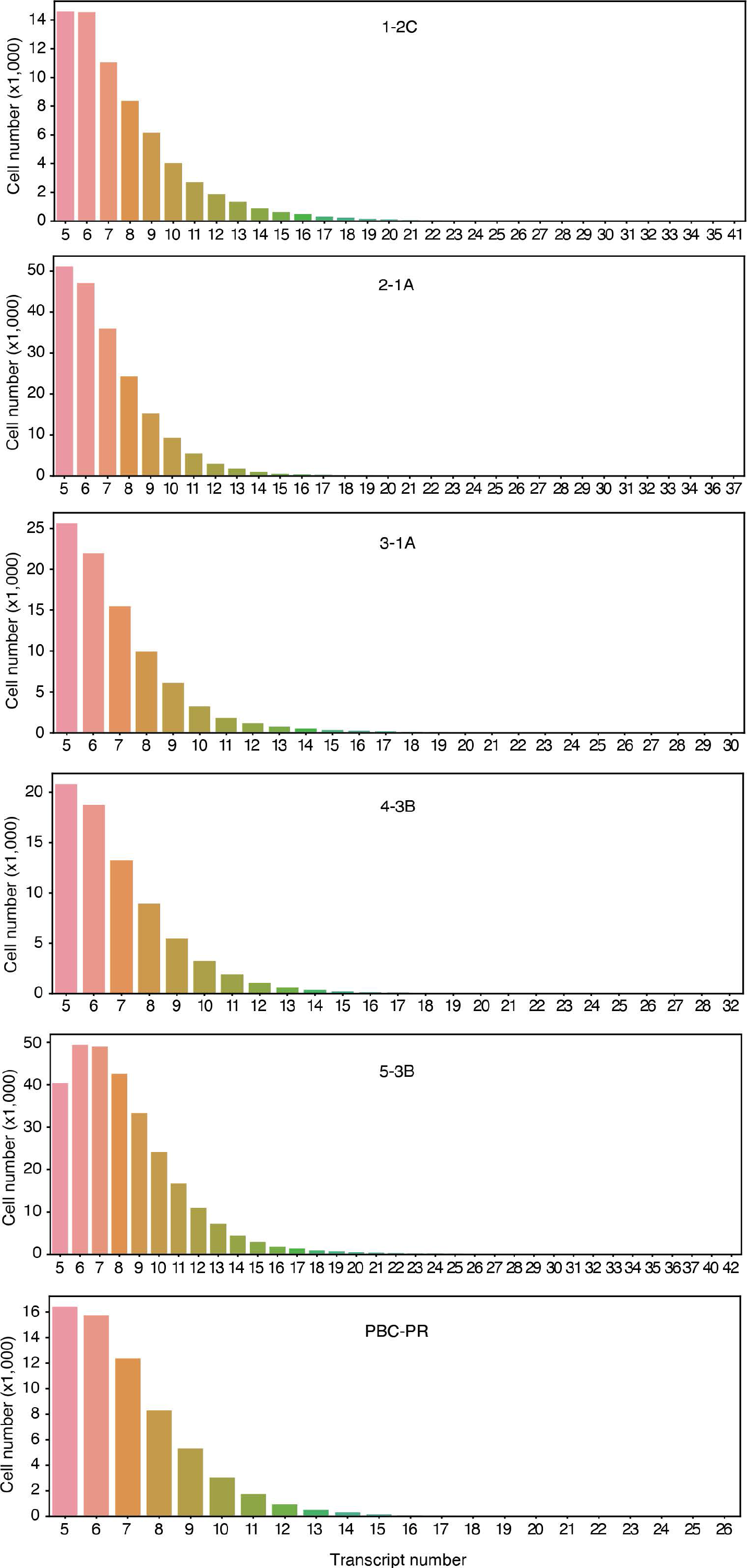
Bar plots showing the distribution of the number of transcripts (reads) that are segmented to each cell. Over 99% of the cells harbored at least 5-15 reads.

**Supplementary Figure 6:**
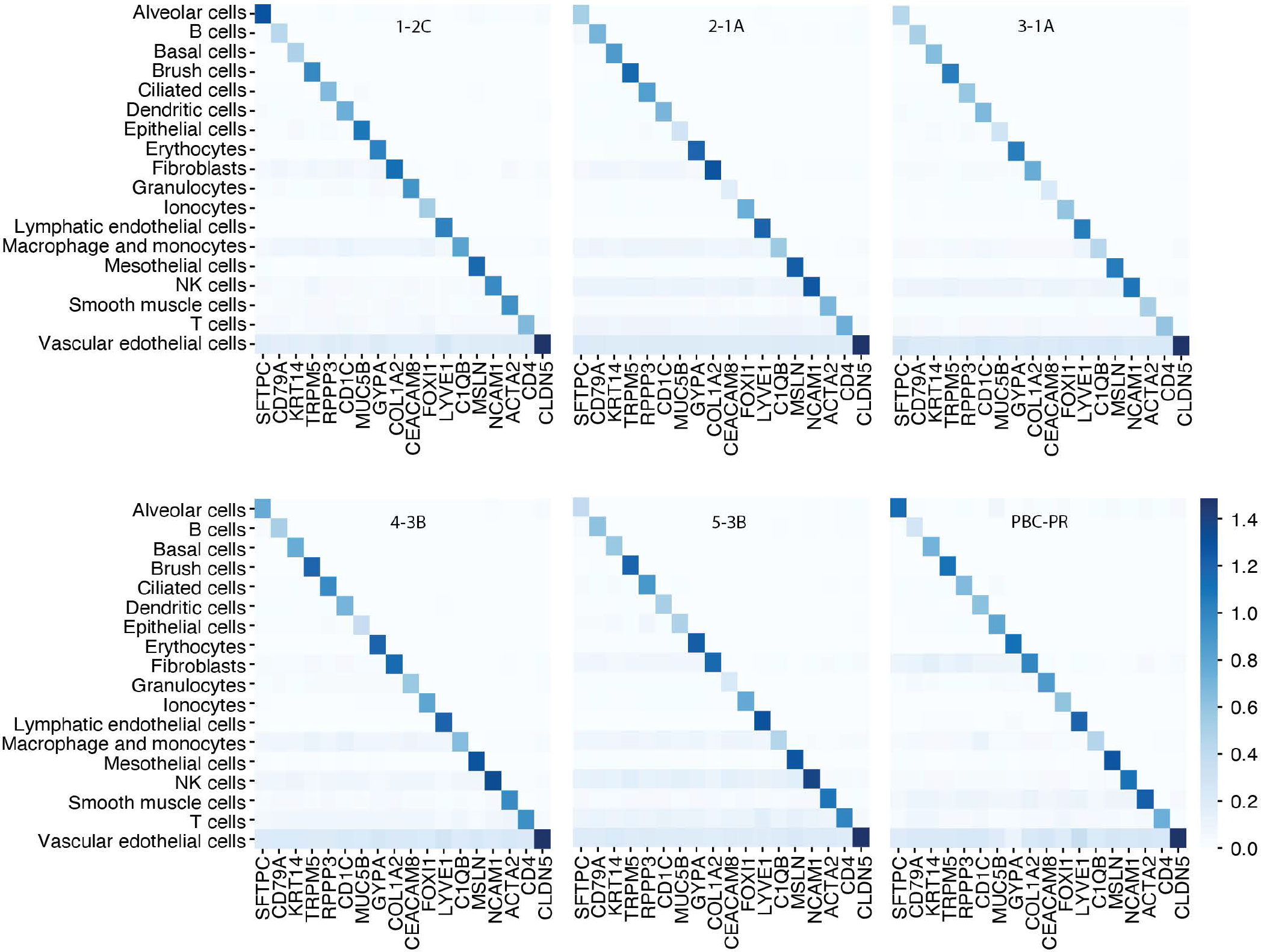
Heatmaps illustrating the average expressions of marker genes in each cell type within each tissue. In all tissues, a total of 18 cell types were identified including 11 types of parenchymal cells and 7 types of immune cells.

**Supplementary Figure 7:**
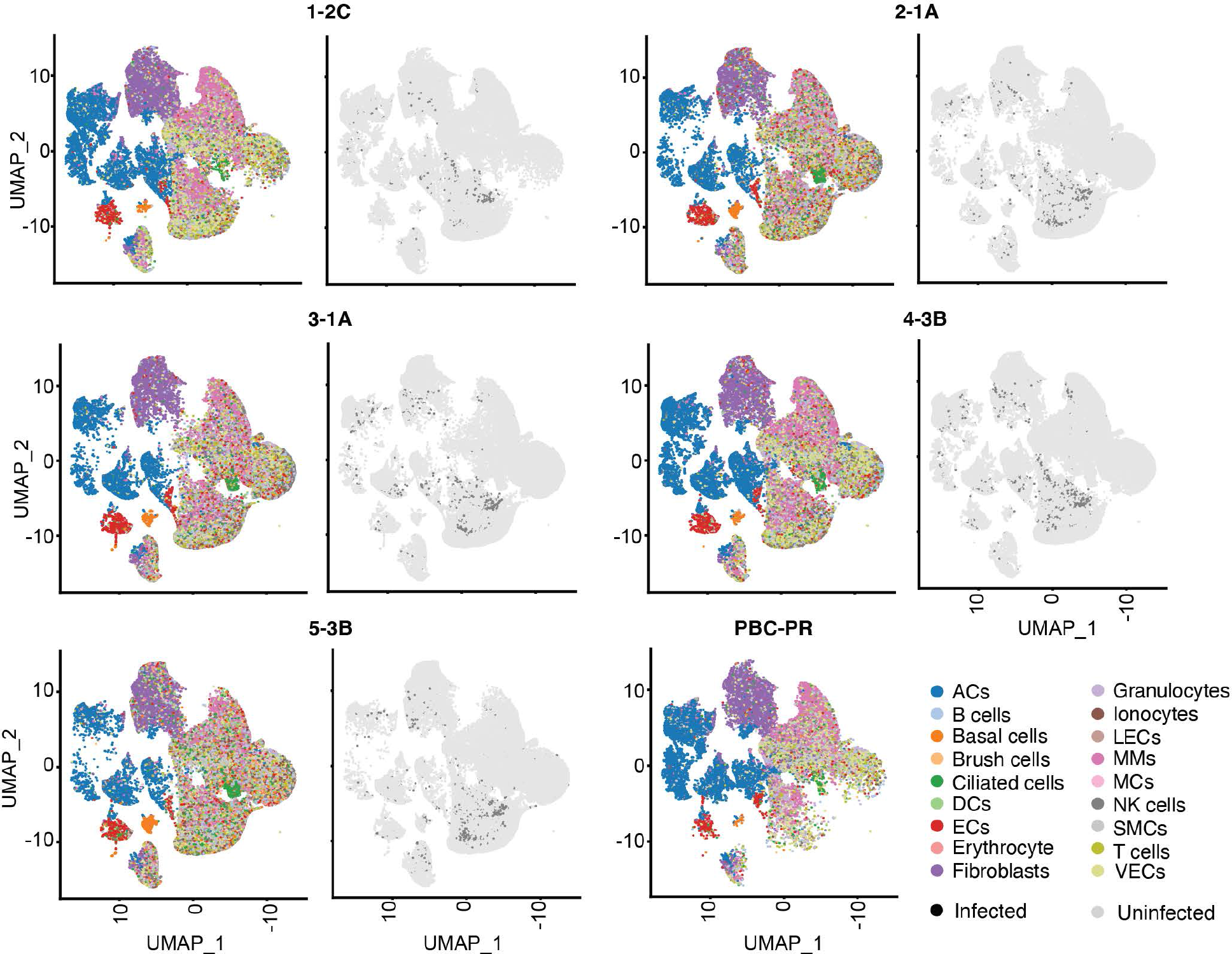
Uniform Manifold Approximation and Projection (UMAP) of the gene expressions of individual cells revealed separated clusters for major parenchymal cells including alveolar cells (ACs) and fibroblasts. By contrast, immune cells were grouped together and mixed with vascular endothelial cells (VECs) in several clusters. The poor separation of immune cell types was largely due to highly expressed Immunoglobulin kappa light chain (IGKC) and Cathepsin L (CTSL) in these cell types, which is further illustrated in Fig. S8.

**Supplementary Figure 8:**
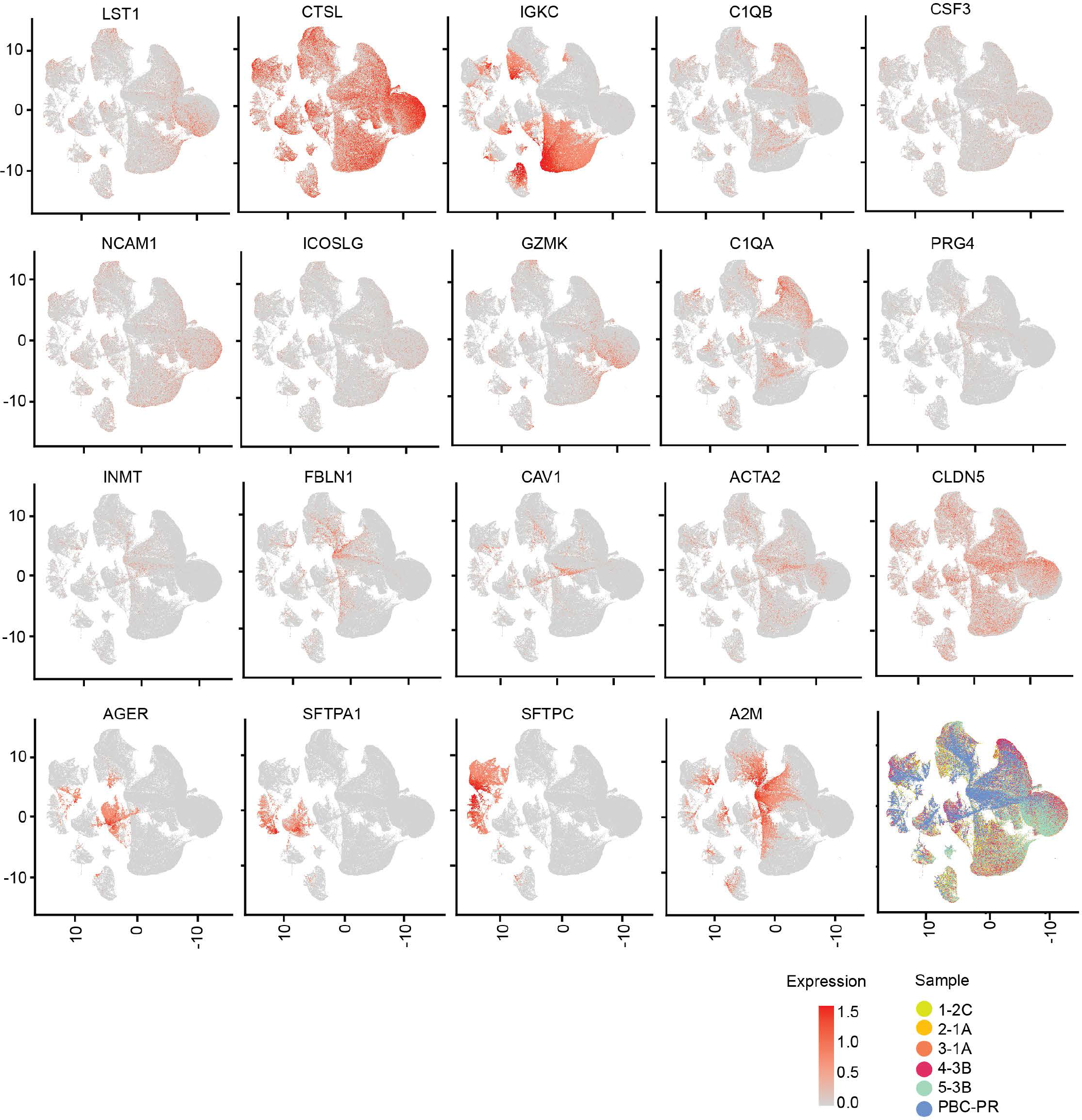
The UMAP of the gene expressions of all segmented and cell-typed cells across all tissues under study is illustrated in the bottom-right panel. Here, the color of the dots indicates the tissue samples. The rest of the UMAPs illustrate the expressions of different genes colored based on their expression in each cell. We observe that IGKC and CTSL are spread across many clusters, resulting in the poor separation of immune cell types which are mixed with VECs.

**Supplementary Figure 9:**
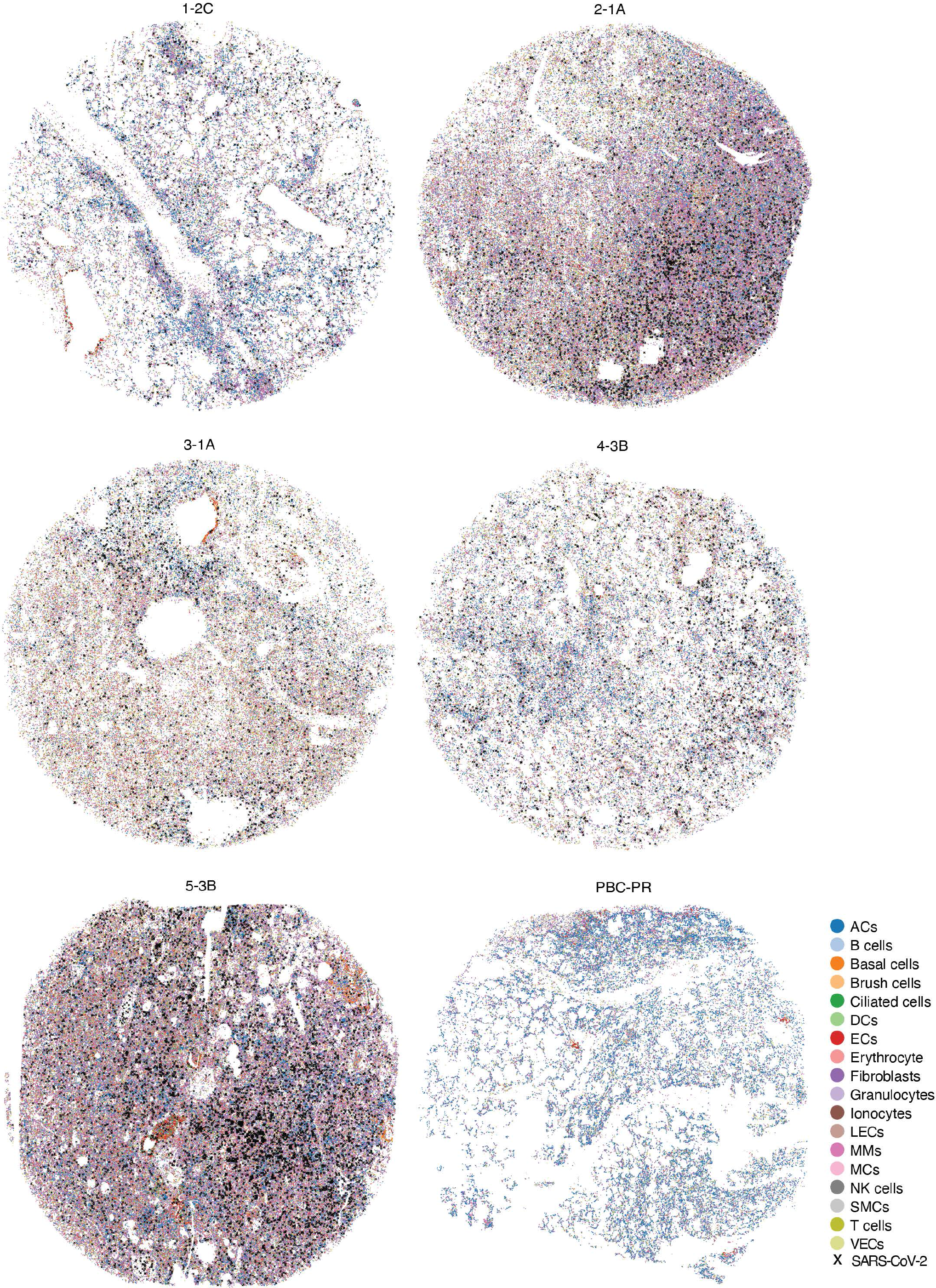
Spatial visualization of the segmented cells painted based on their cell types along with the location of the SARS-CoV-2 reads marked with an ‘X’.

**Supplementary Figure 10:**
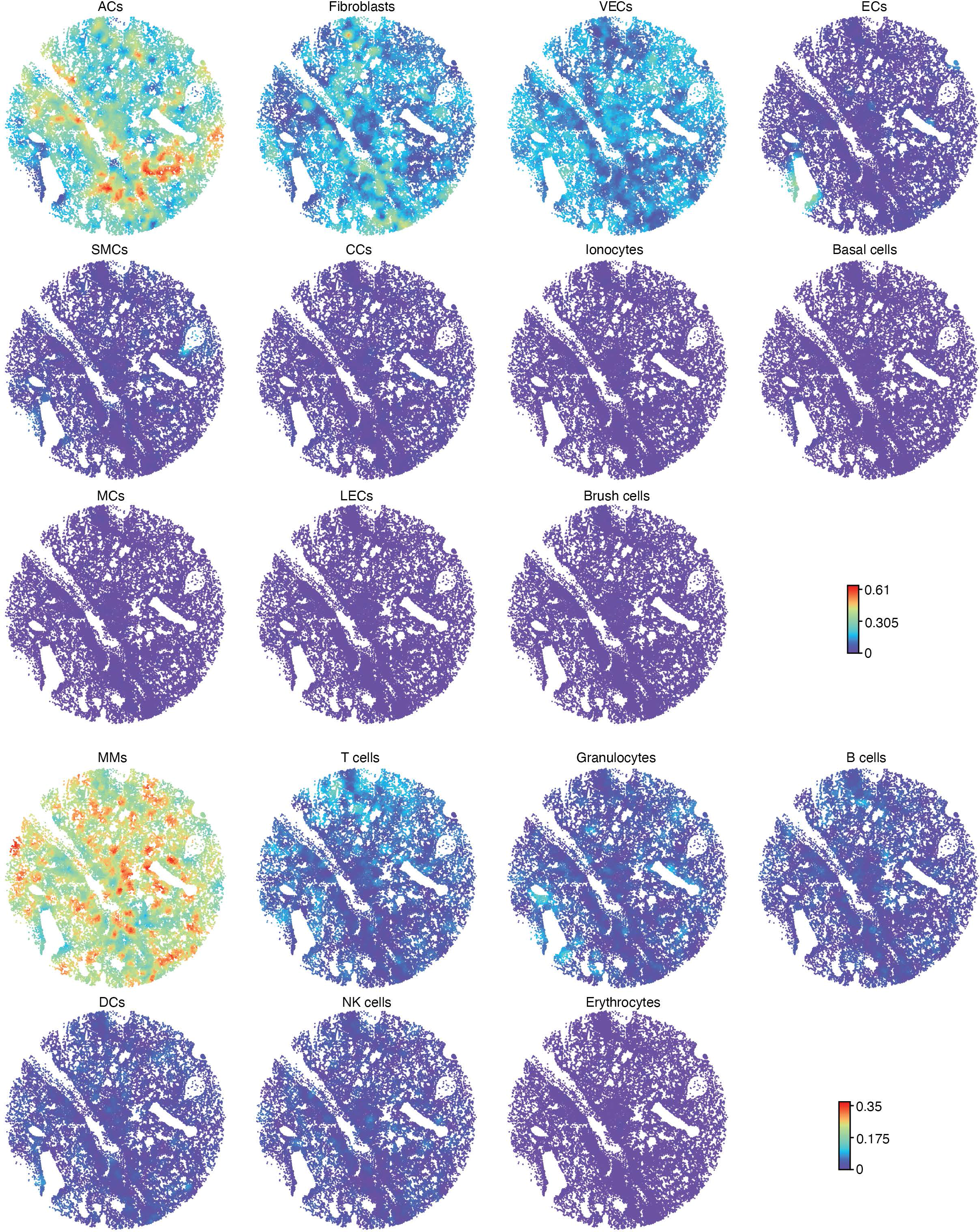
The neighborhood cell type composition (NCTC) analysis for COVID-19 tissue 1-2C is illustrated based on the cell type.

**Supplementary Figure 11:**
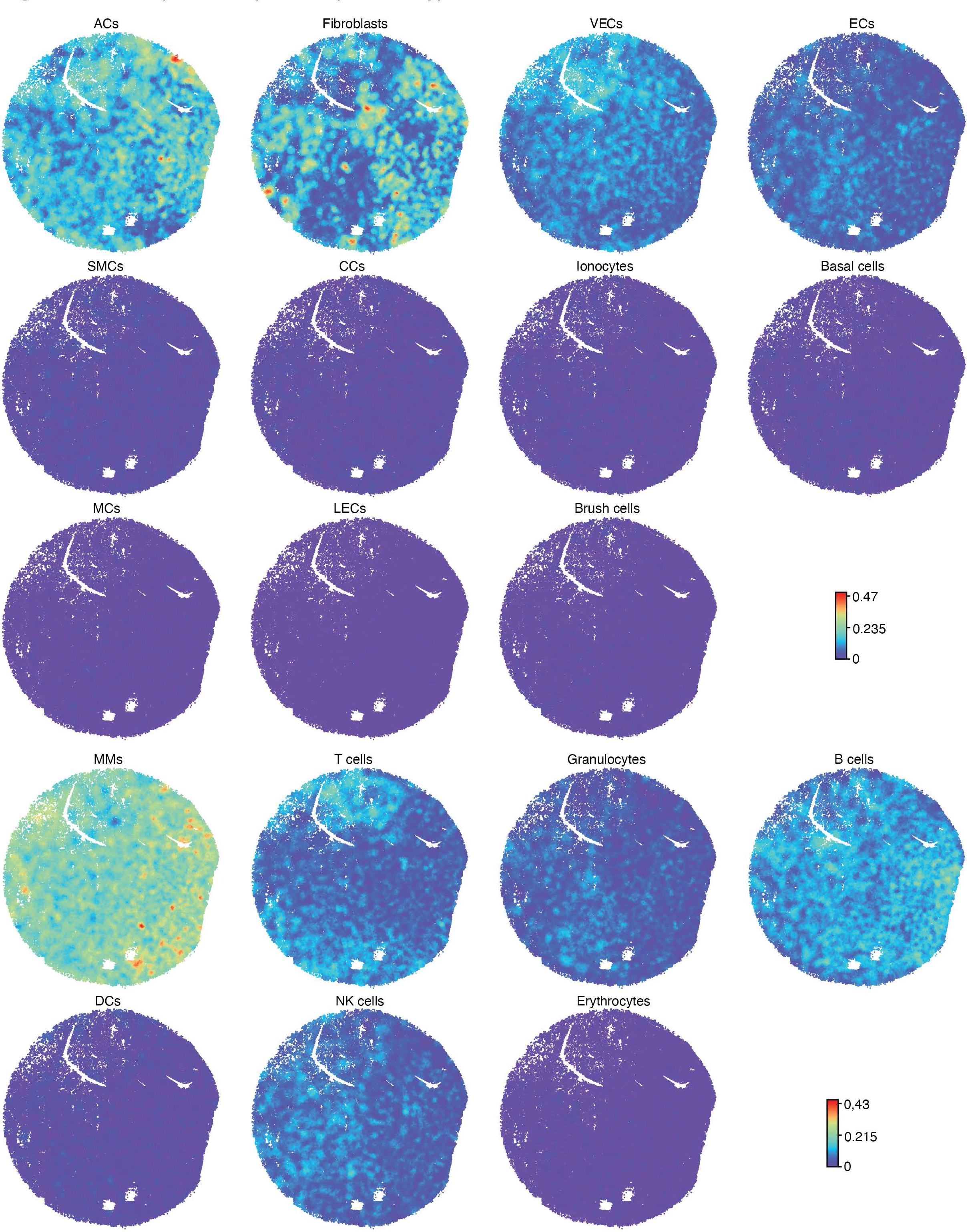
The neighborhood cell type composition (NCTC) analysis for COVID-19 tissue 2-1A is illustrated based on the cell type.

**Supplementary Figure 12:**
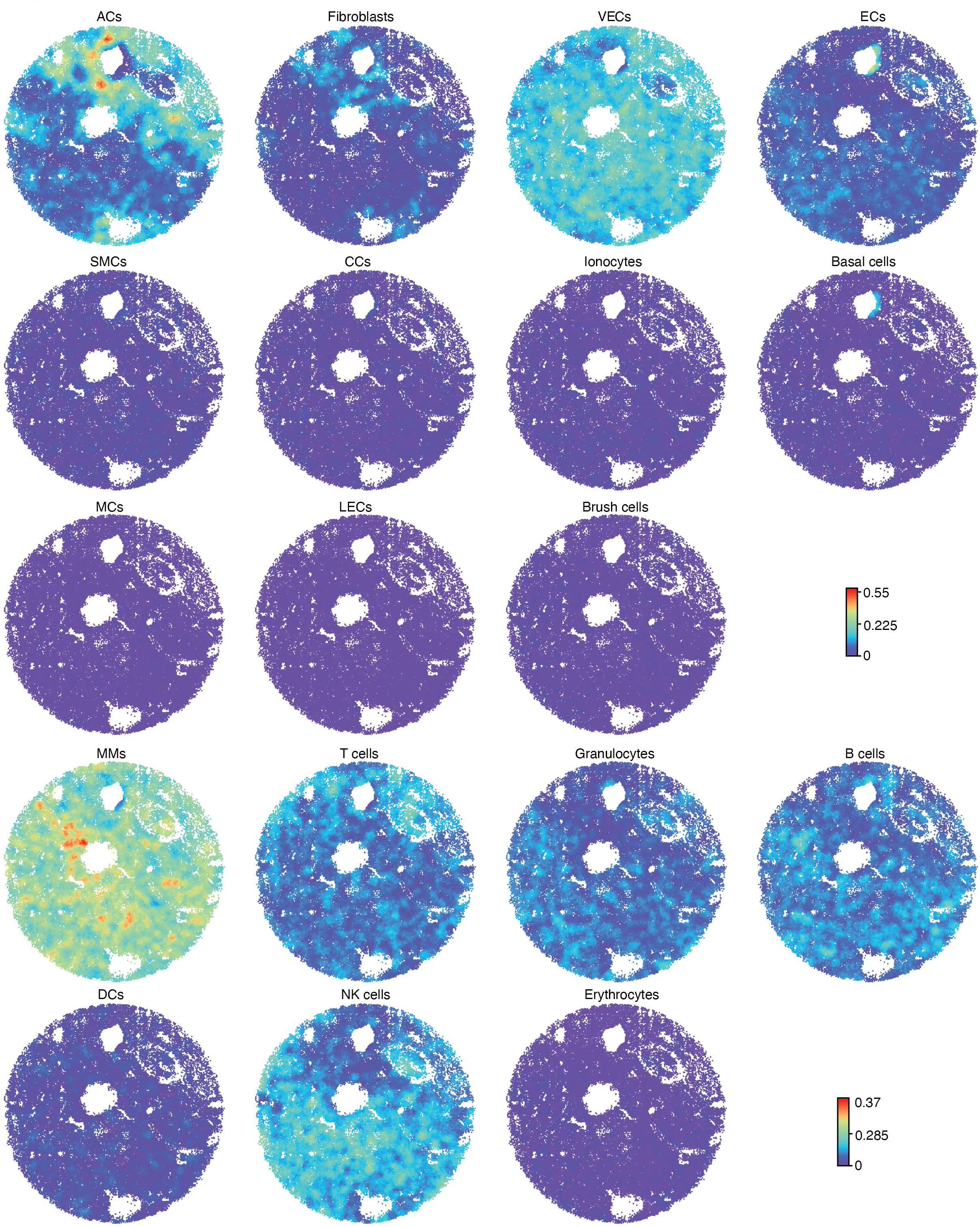
The neighborhood cell type composition (NCTC) analysis for COVID-19 tissue 3-1A is illustrated based on the cell type.

**Supplementary Figure 13:**
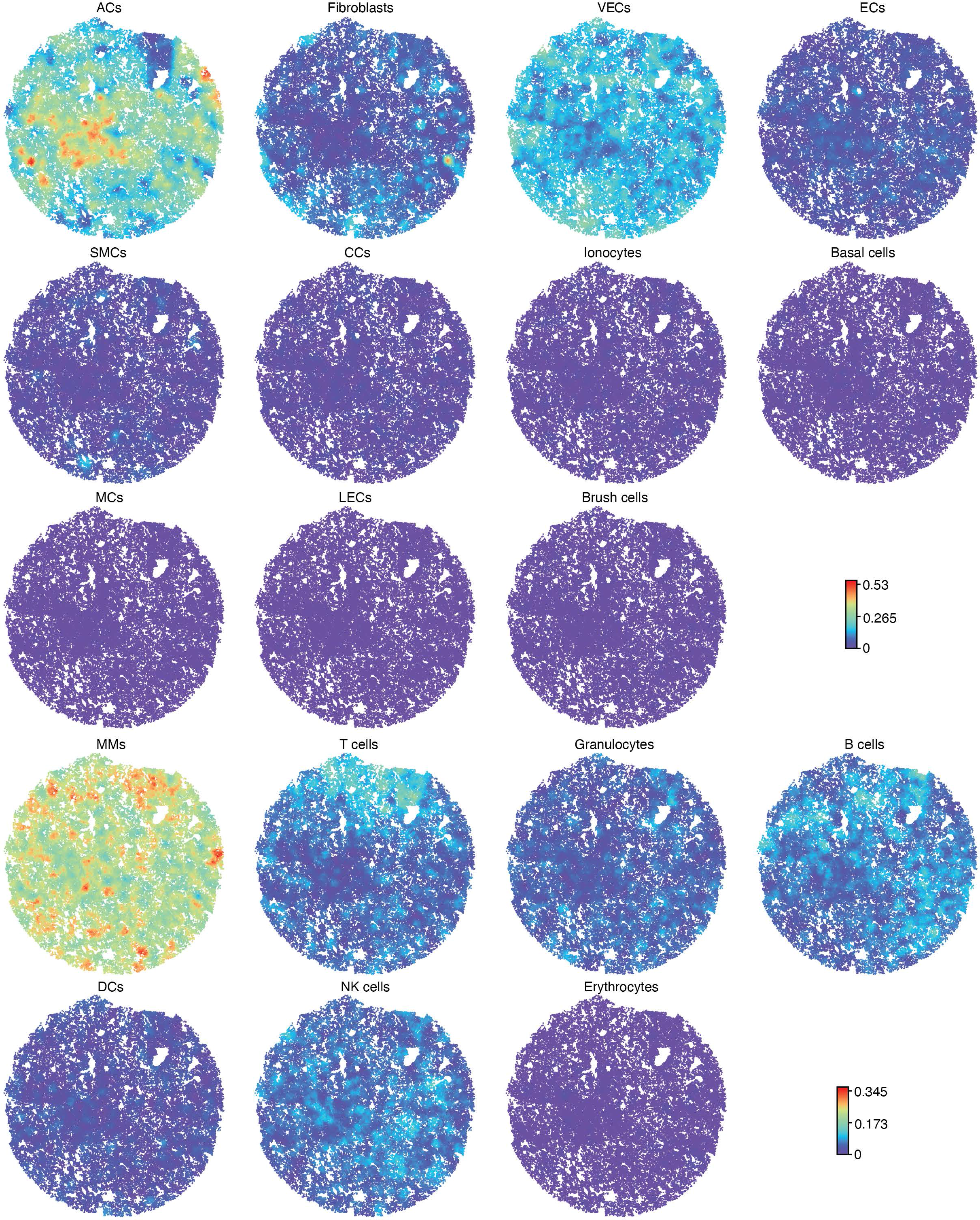
The neighborhood cell type composition (NCTC) analysis for COVID-19 tissue 4-3B is illustrated based on the cell type.

**Supplementary Figure 14:**
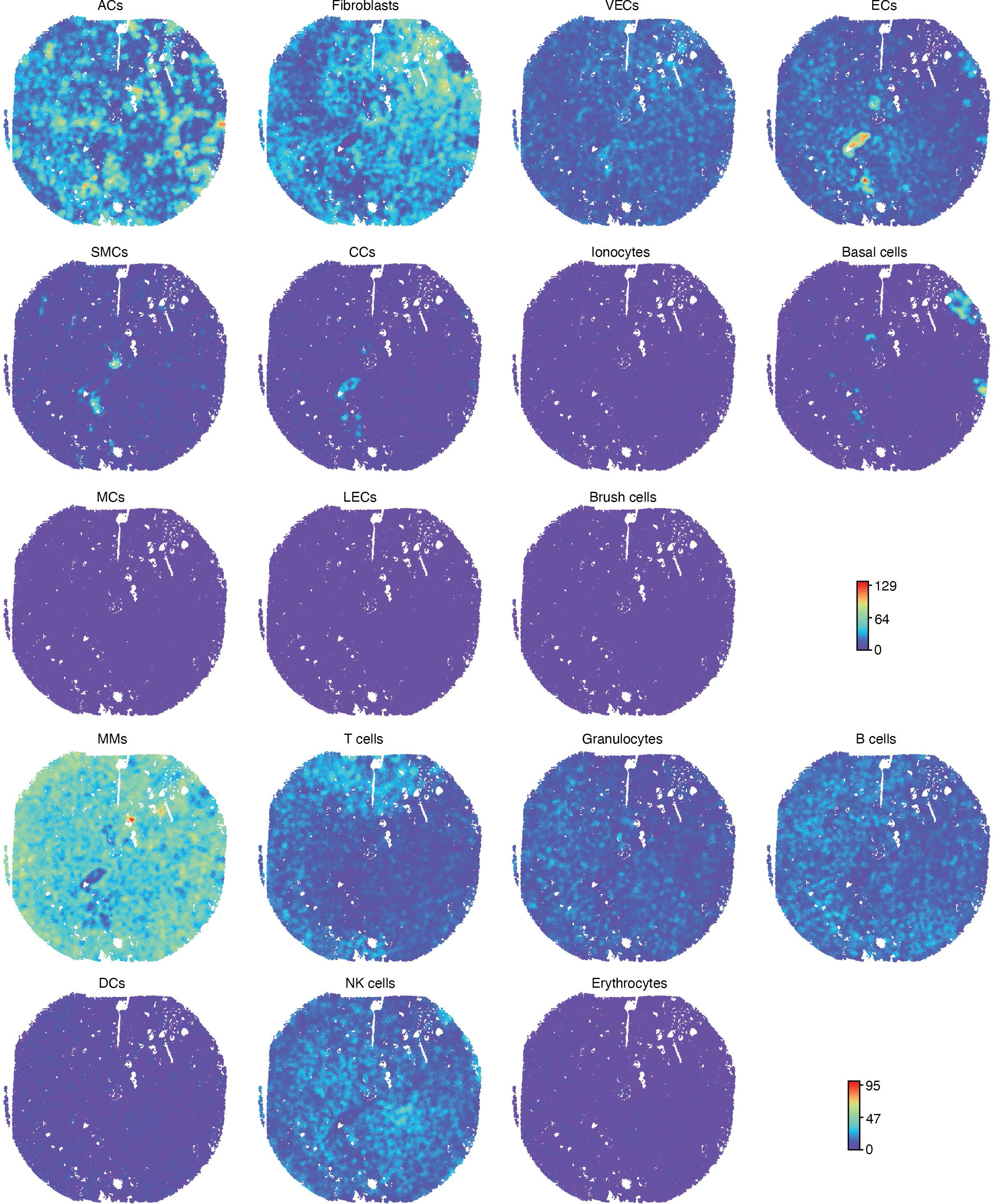
The neighborhood cell type composition (NCTC) analysis for COVID-19 tissue 5-3B is illustrated based on the cell type.

**Supplementary Figure 15:**
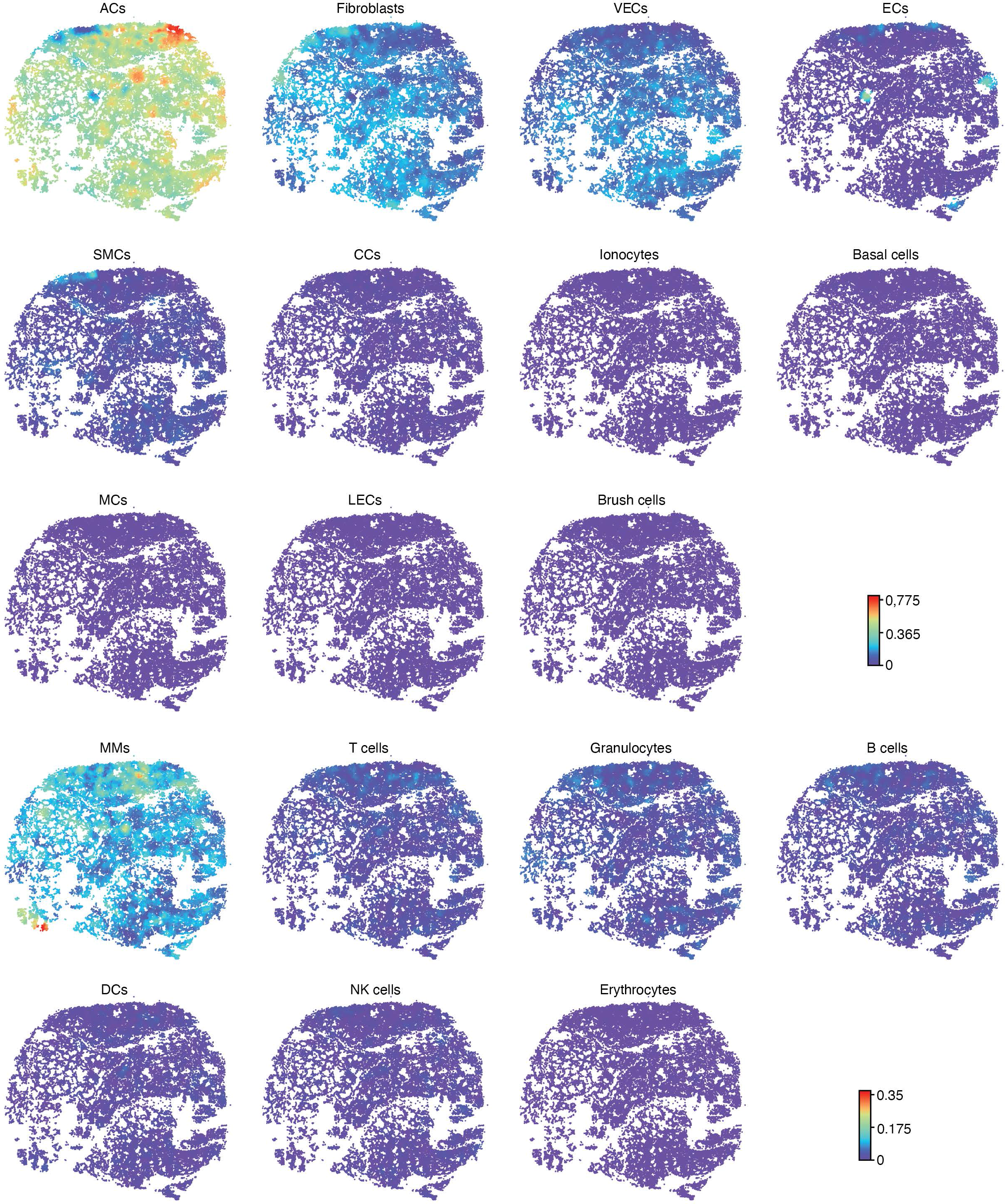
The neighborhood cell type composition (NCTC) analysis for non-COVID-19 tissue PBC-PR is illustrated based on the cell type.

**Supplementary Figure 16:**
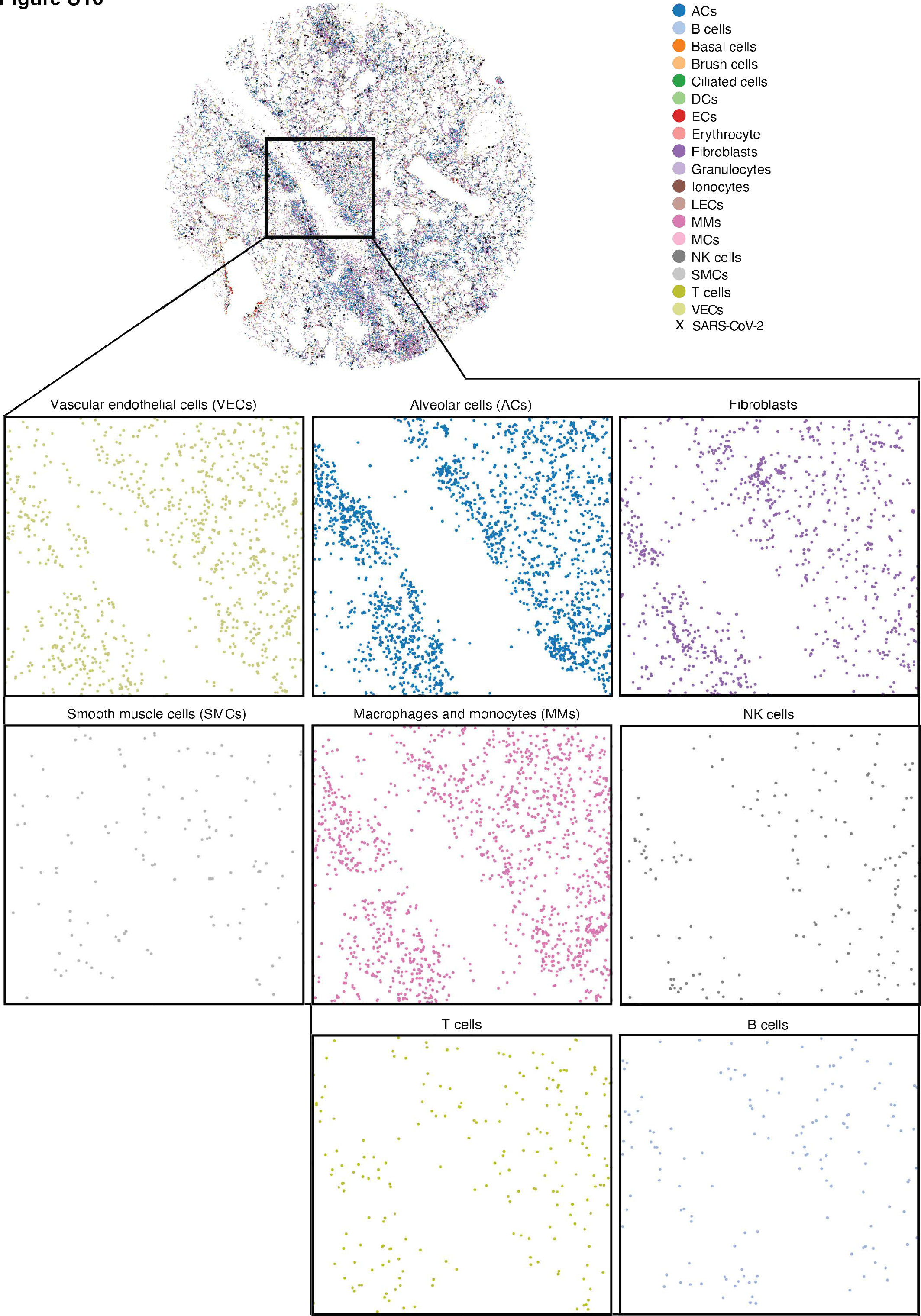
Spatial visualization of regions with blood vessels revealed the lining of VECs along the vessels together with other vessel-associated cells including ACs, fibroblasts and smooth muscle cells (SMCs). Each dot indicates a cell, and the color of each dot indicates the cell type.

**Supplementary Figure 17:**
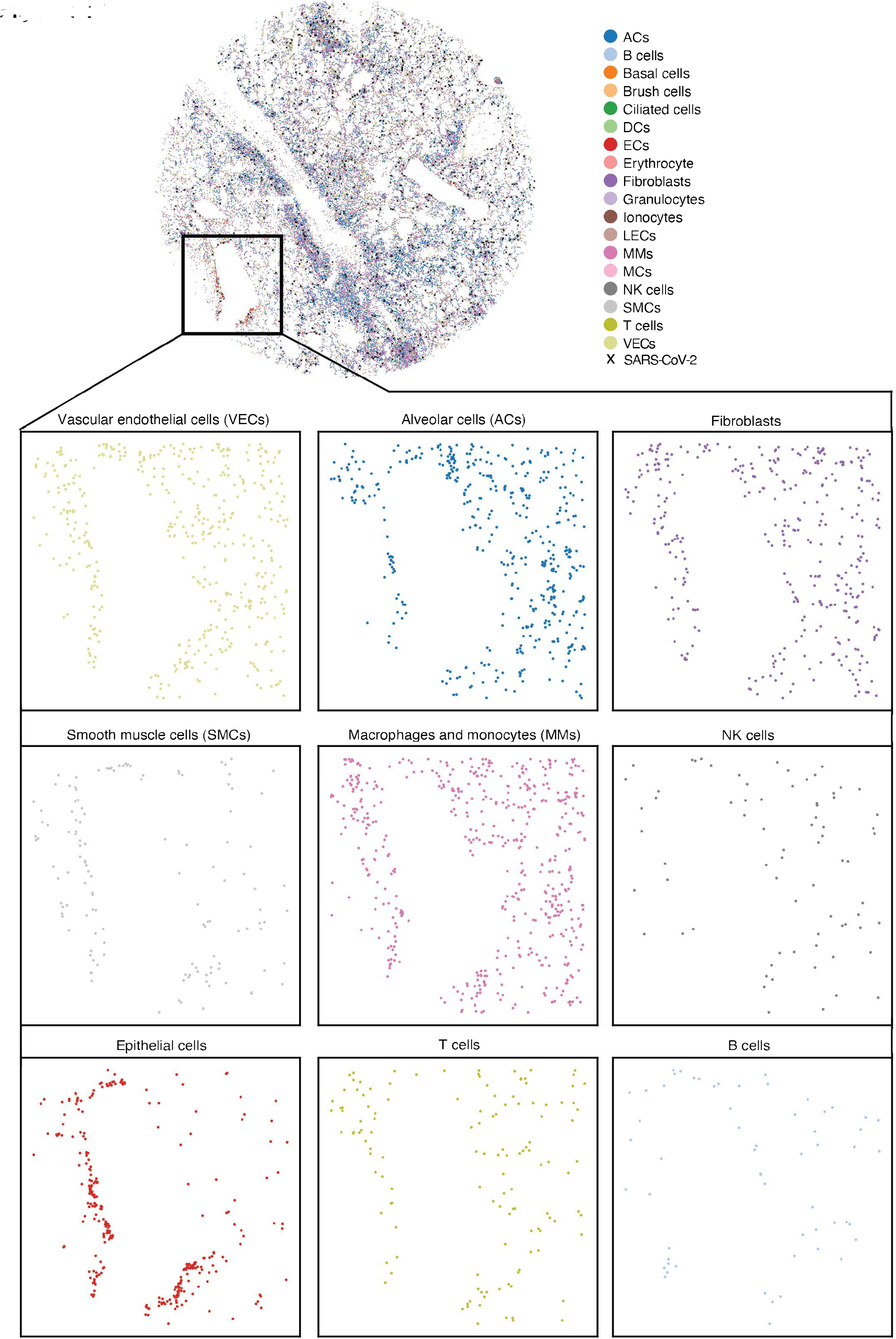
Spatial visualization of a representative region with bronchioles revealed the lining of epithelial cells (ECs), smooth muscle cells (SMCs) and VECs along the bronchiole together with other bronchiole-associated cells including ACs and fibroblasts. Each dot indicates a cell, and the color of each dot indicates the cell type.

**Supplementary Figure 18:**
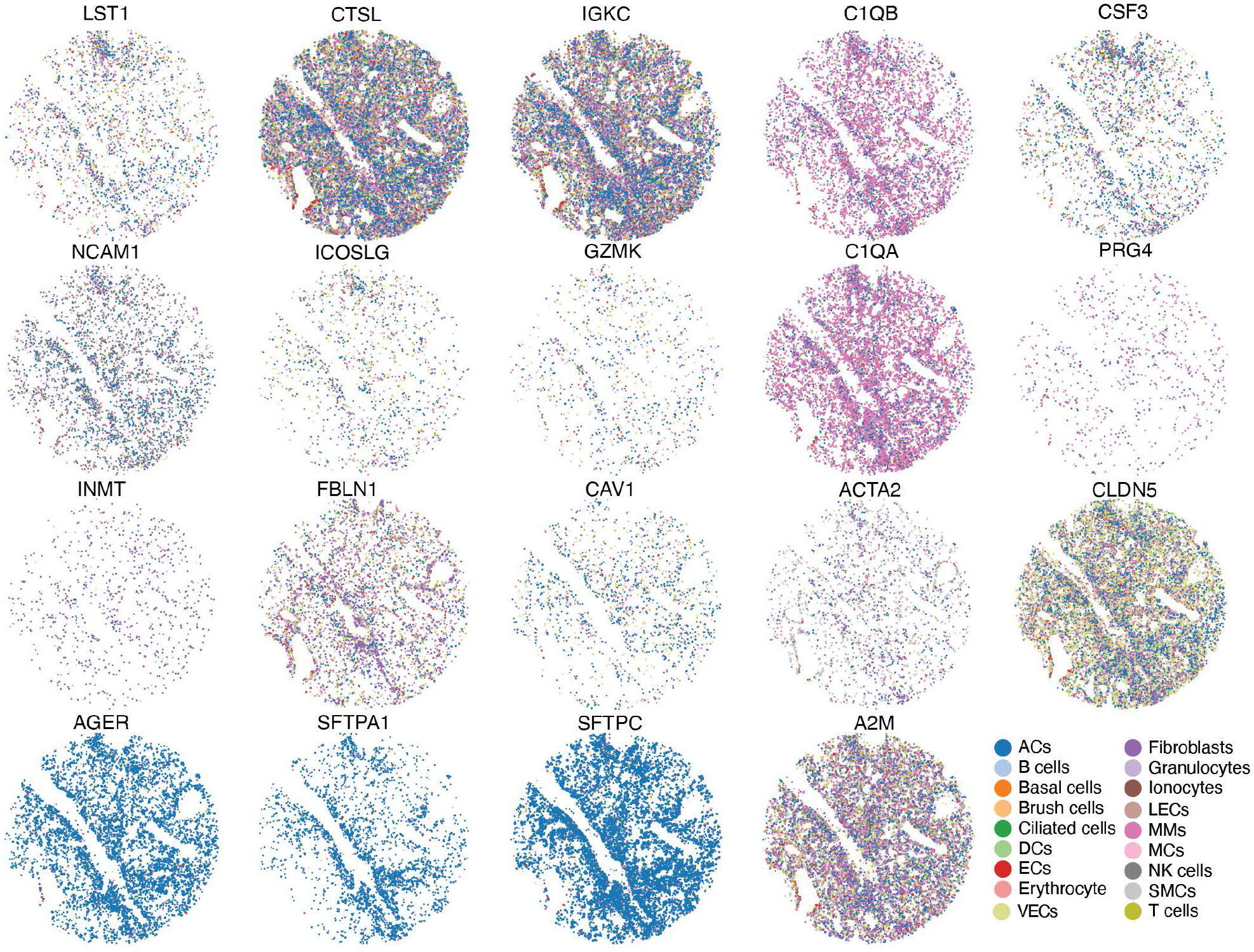
Spatial visualization of the expressions of the 19 dysregulated genes in different cell types and their tissue distributions in COVID-19 tissue 1-2C are illustrated. We observe significant variations in the expression patterns suggesting their complex involvements in different aspects of SARS-CoV-2 infection and COVID-19 lung pathology. The color of each dot indicates the cell type, and the size indicates their expression level.

**Supplementary Figure 19:**
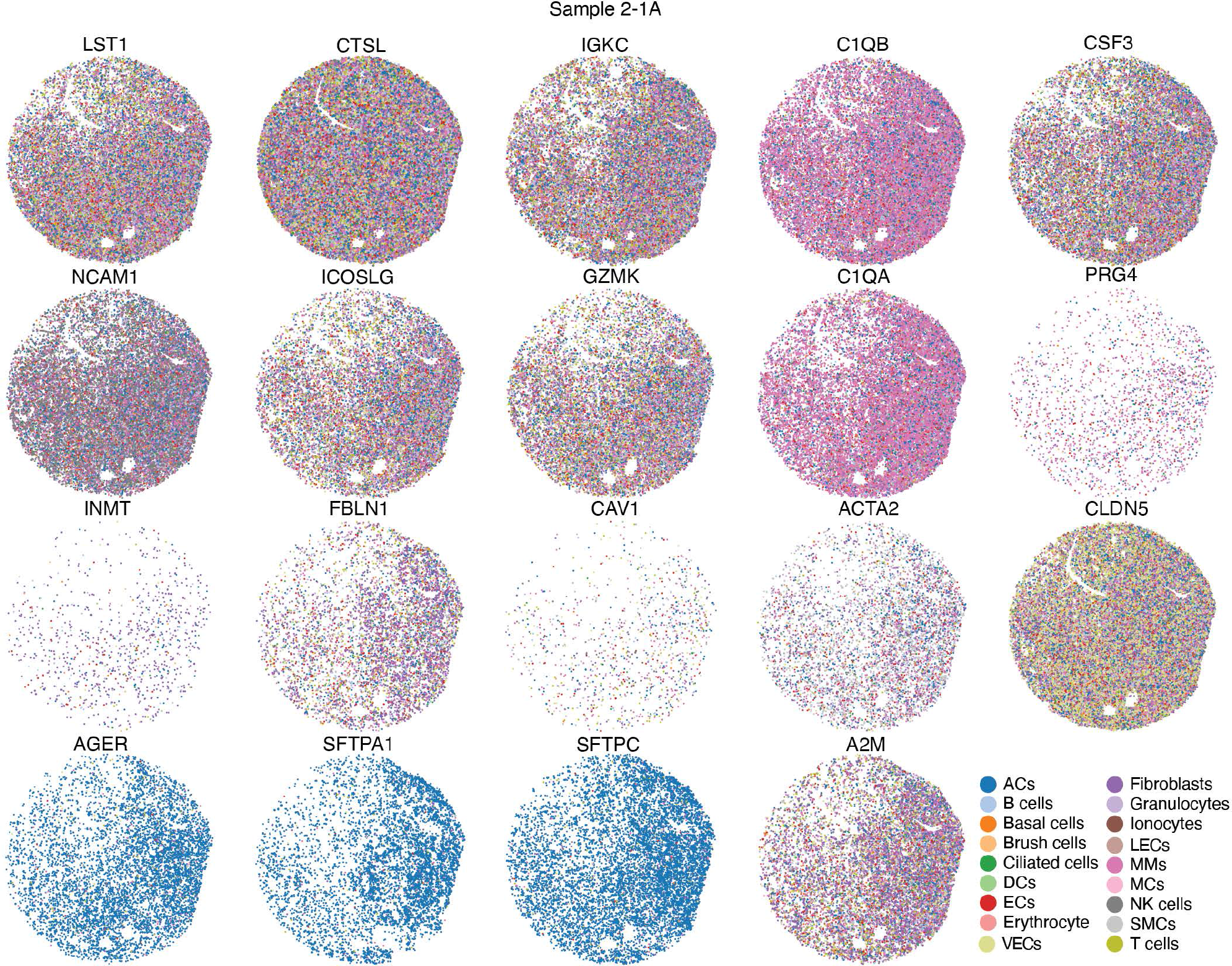
Spatial visualization of the expressions of the 19 dysregulated genes in different cell types and their tissue distributions in COVID-19 tissue 2-1A are illustrated. We observe significant variations in the expression patterns suggesting their complex involvements in different aspects of SARS-CoV-2 infection and COVID-19 lung pathology. The color of each dot indicates the cell type, and the size indicates their expression level.

**Supplementary Figure 20:**
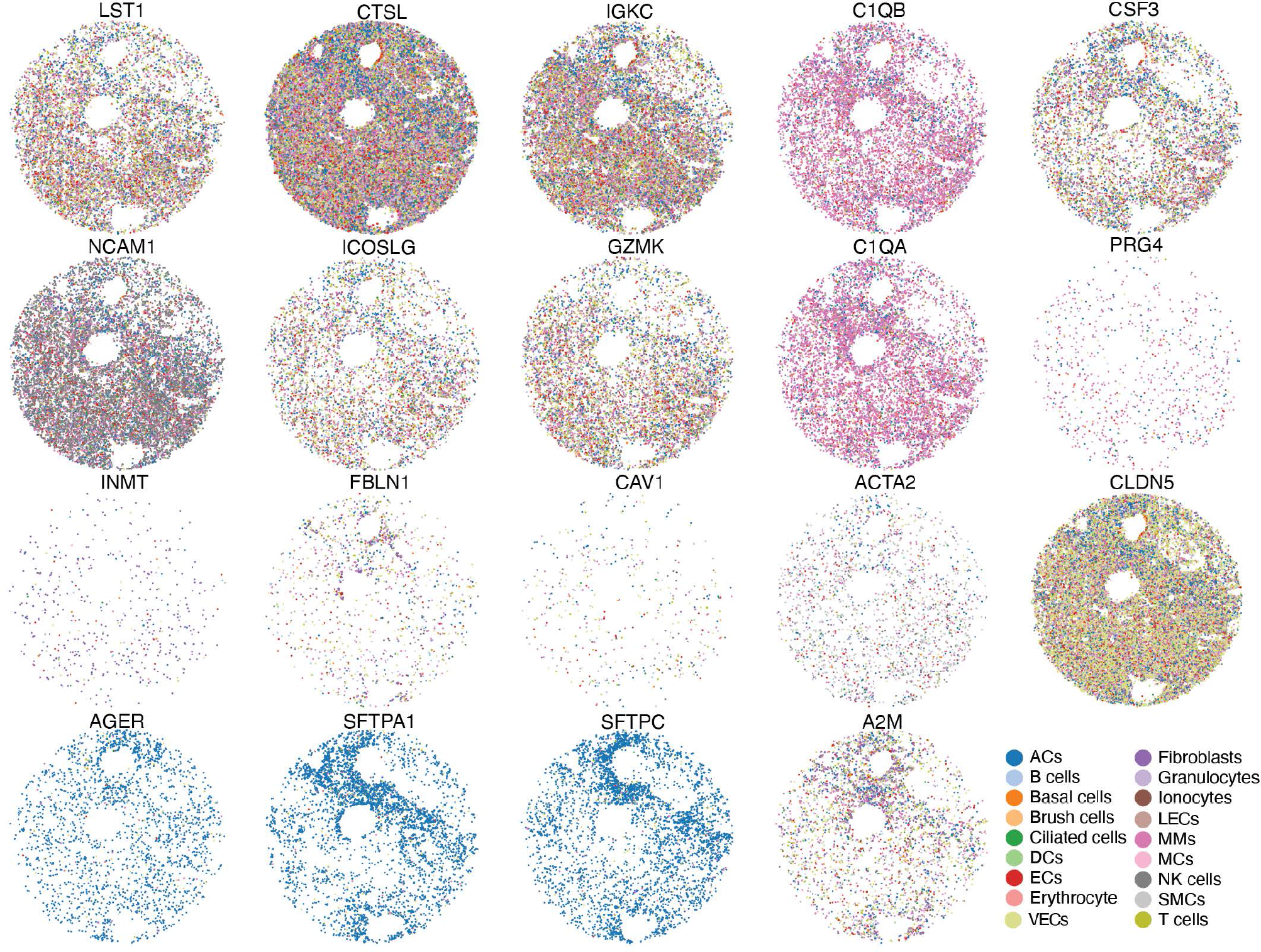
Spatial visualization of the expressions of the 19 dysregulated genes in different cell types and their tissue distributions in COVID-19 tissue 3-1A are illustrated. We observe significant variations in the expression patterns suggesting their complex involvements in different aspects of SARS-CoV-2 infection and COVID-19 lung pathology. The color of each dot indicates the cell type, and the size indicates their expression level.

**Supplementary Figure 21:**
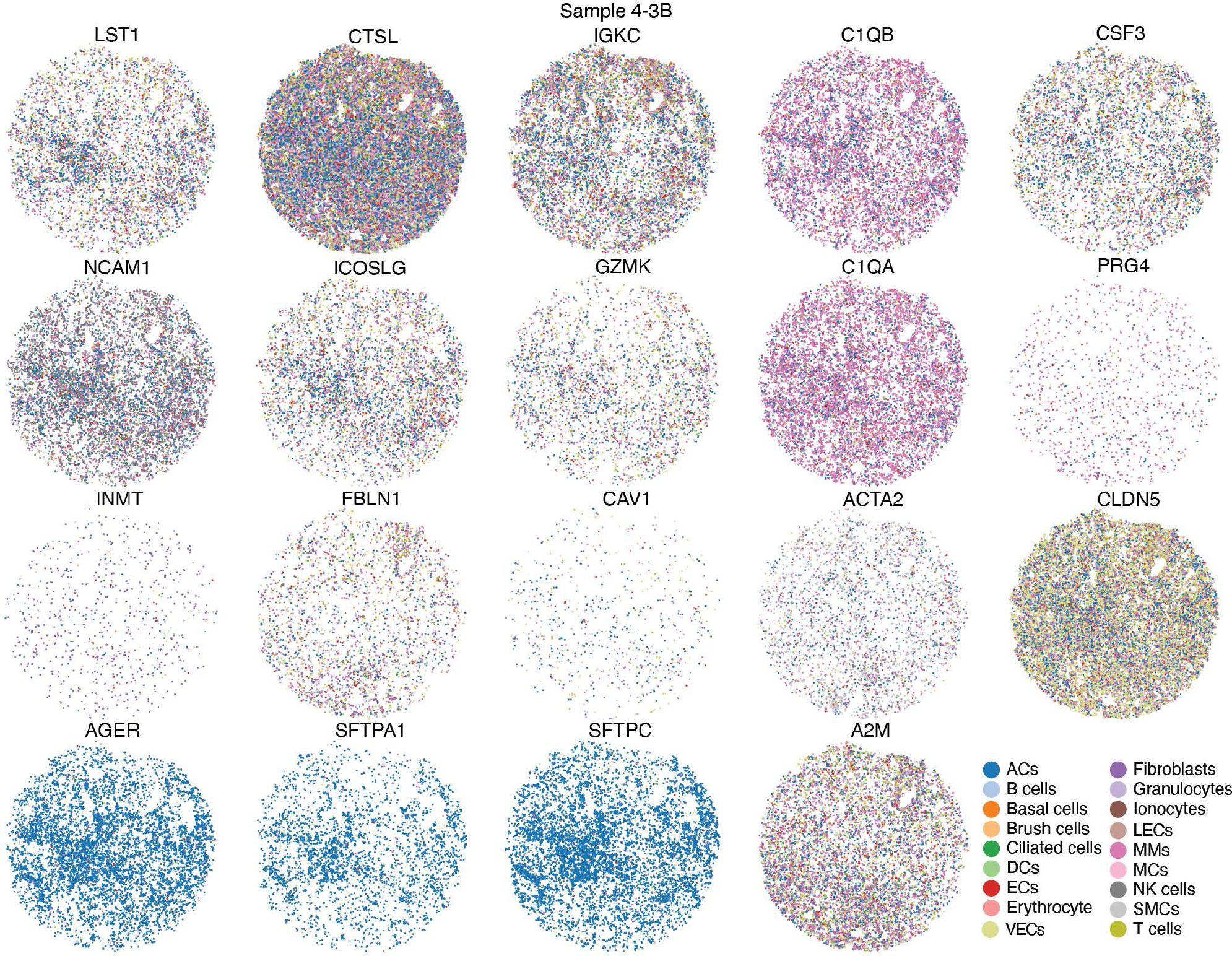
Spatial visualization of the expressions of the 19 dysregulated genes in different cell types and their tissue distributions in COVID-19 tissue 4-3B are illustrated. We observe significant variations in the expression patterns suggesting their complex involvements in different aspects of SARS-CoV-2 infection and COVID-19 lung pathology. The color of each dot indicates the cell type, and the size indicates their expression level.

**Supplementary Figure 22:**
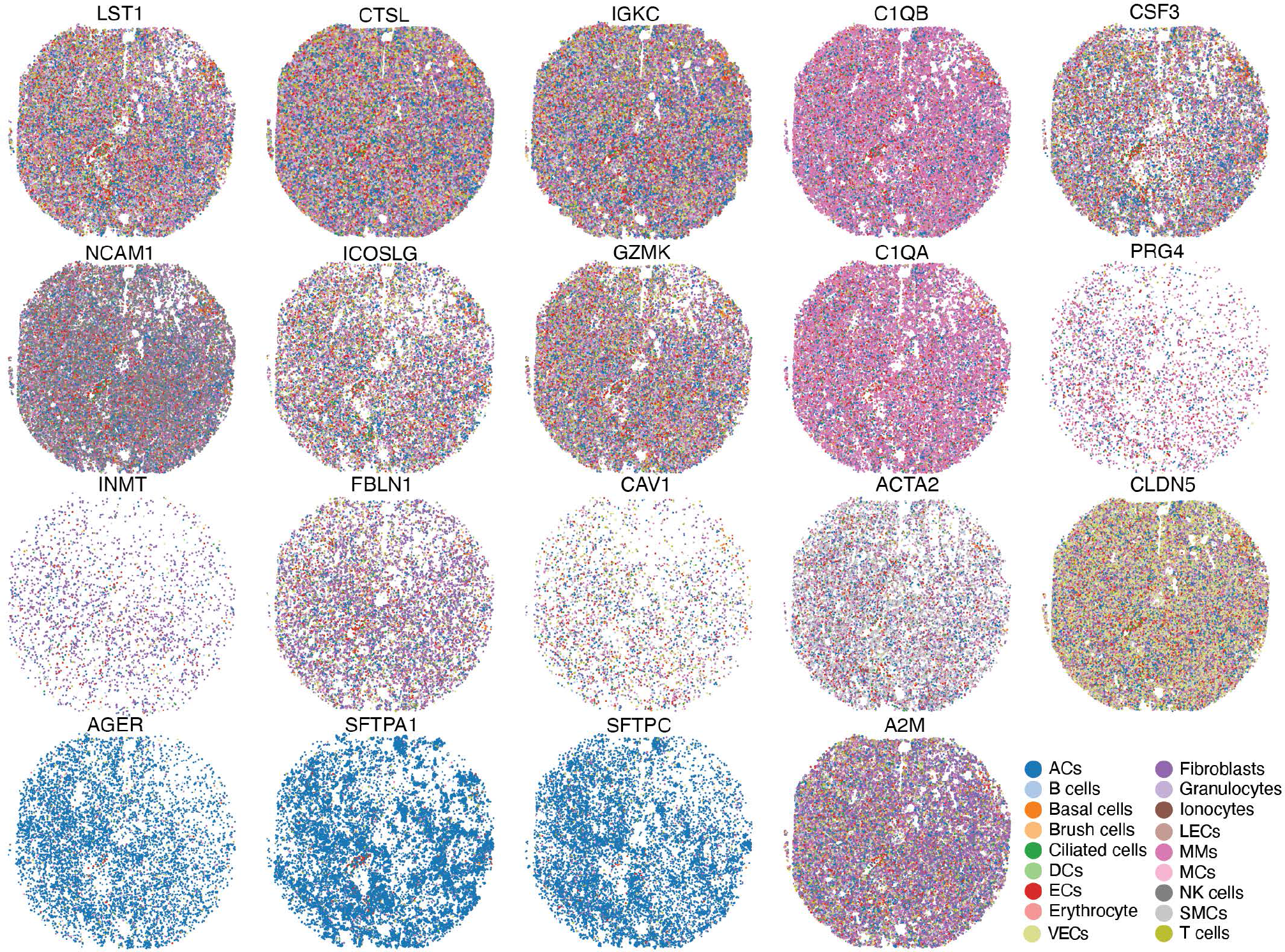
Spatial visualization of the expressions of the 19 dysregulated genes in different cell types and their tissue distributions in COVID-19 tissue 5-3B are illustrated. We observe significant variations in the expression patterns suggesting their complex involvements in different aspects of SARS-CoV-2 infection and COVID-19 lung pathology. The color of each dot indicates the cell type, and the size indicates their expression level.

**Supplementary Figure 23:**
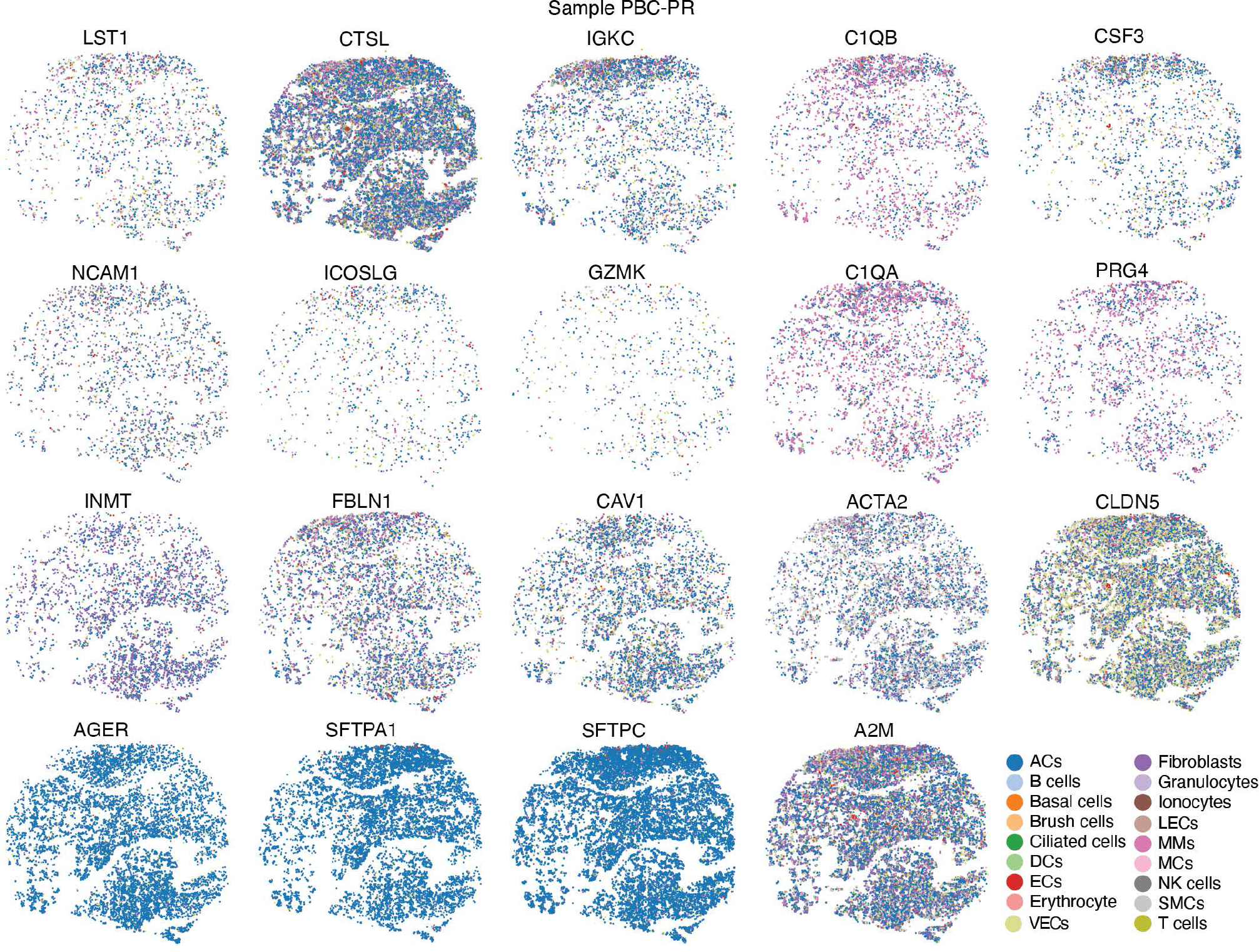
Spatial visualization of the expressions of the 19 dysregulated genes in different cell types and their tissue distributions in non-COVID-19 tissue PBC-PR are illustrated. We observe significant variations in the expression patterns suggesting their complex involvements in different aspects of SARS-CoV-2 infection and COVID-19 lung pathology. The color of each dot indicates the cell type, and the size indicates their expression level.

**Supplementary Figure 24:**
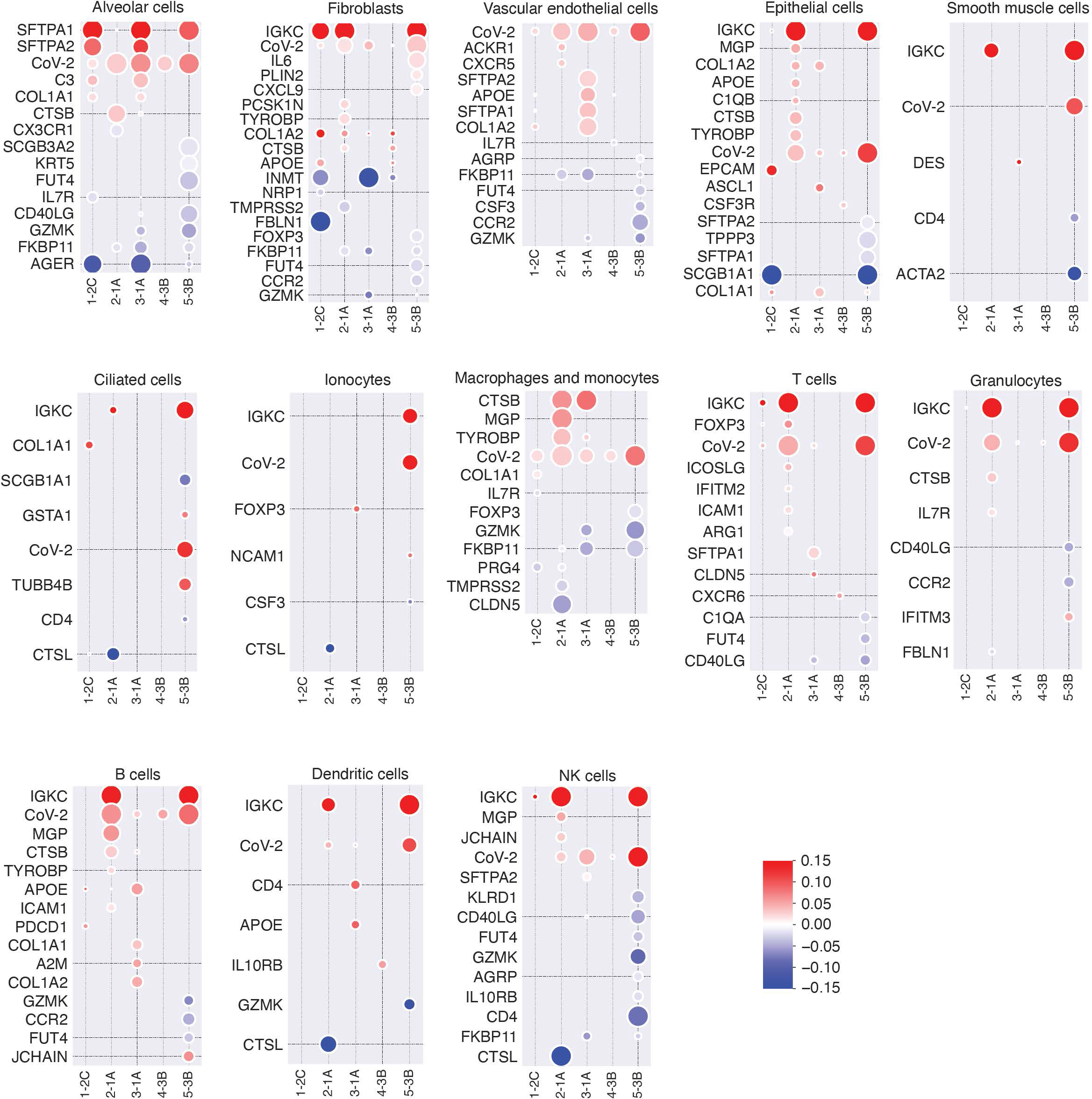
Bubble plots of cell-type specific differential expression analysis of the high infection versus the low infection regions of the same COVID-19 tissues is illustrated. The analysis further confirmed the upregulation of SFTPA1 or SFTPA2 in ACs and COL1A2 in fibroblasts in infected tissues. Within each row, the bubble size indicates the -log10 of the corrected p-value and the color indicates the log2 fold change of the corresponding gene.

**Supplementary Figure 25:**
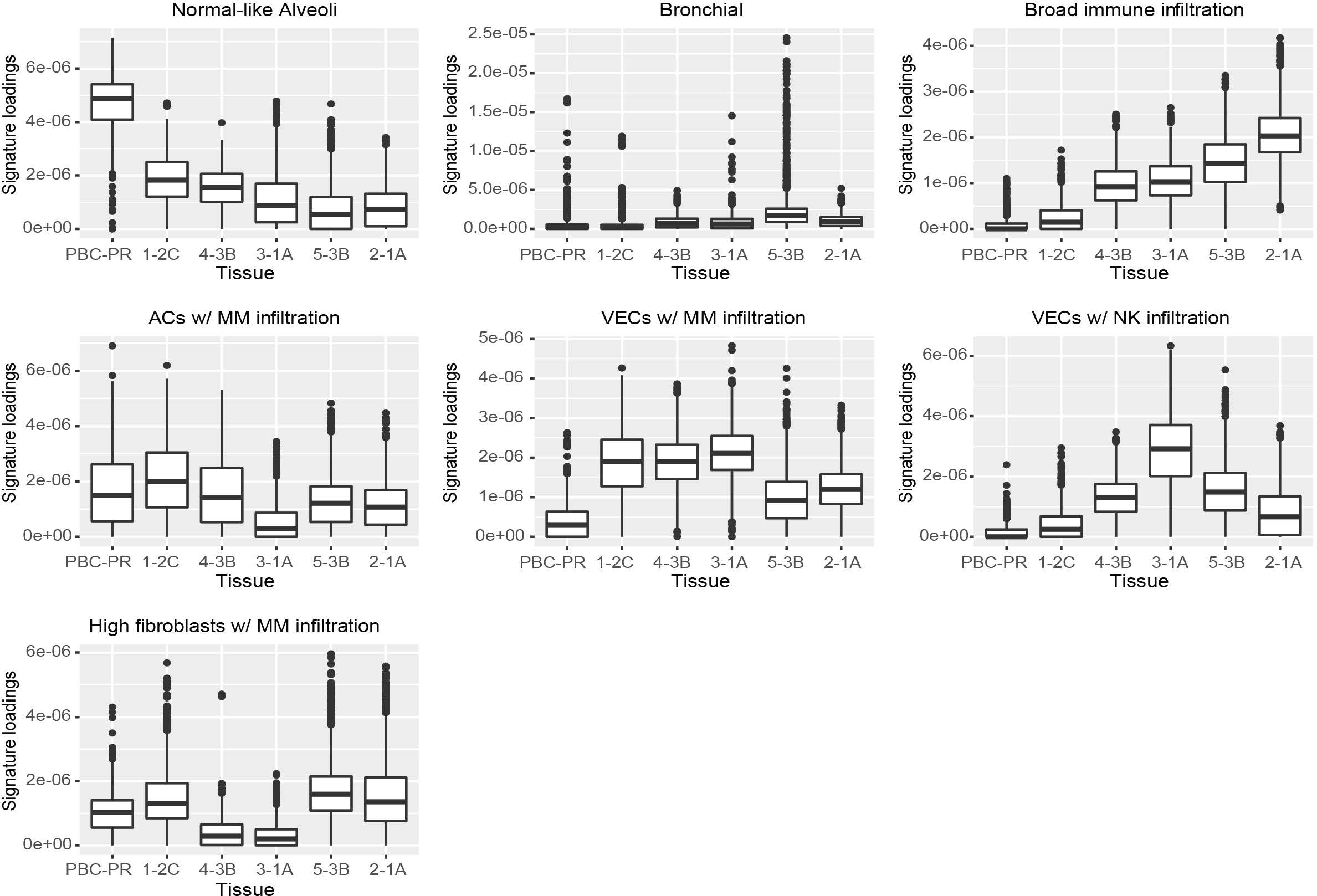
Box plots of signature loadings for each identified signature generated by the SNMF decomposition of NCTC vectors are illustrated.

**Supplementary Figure 26:**
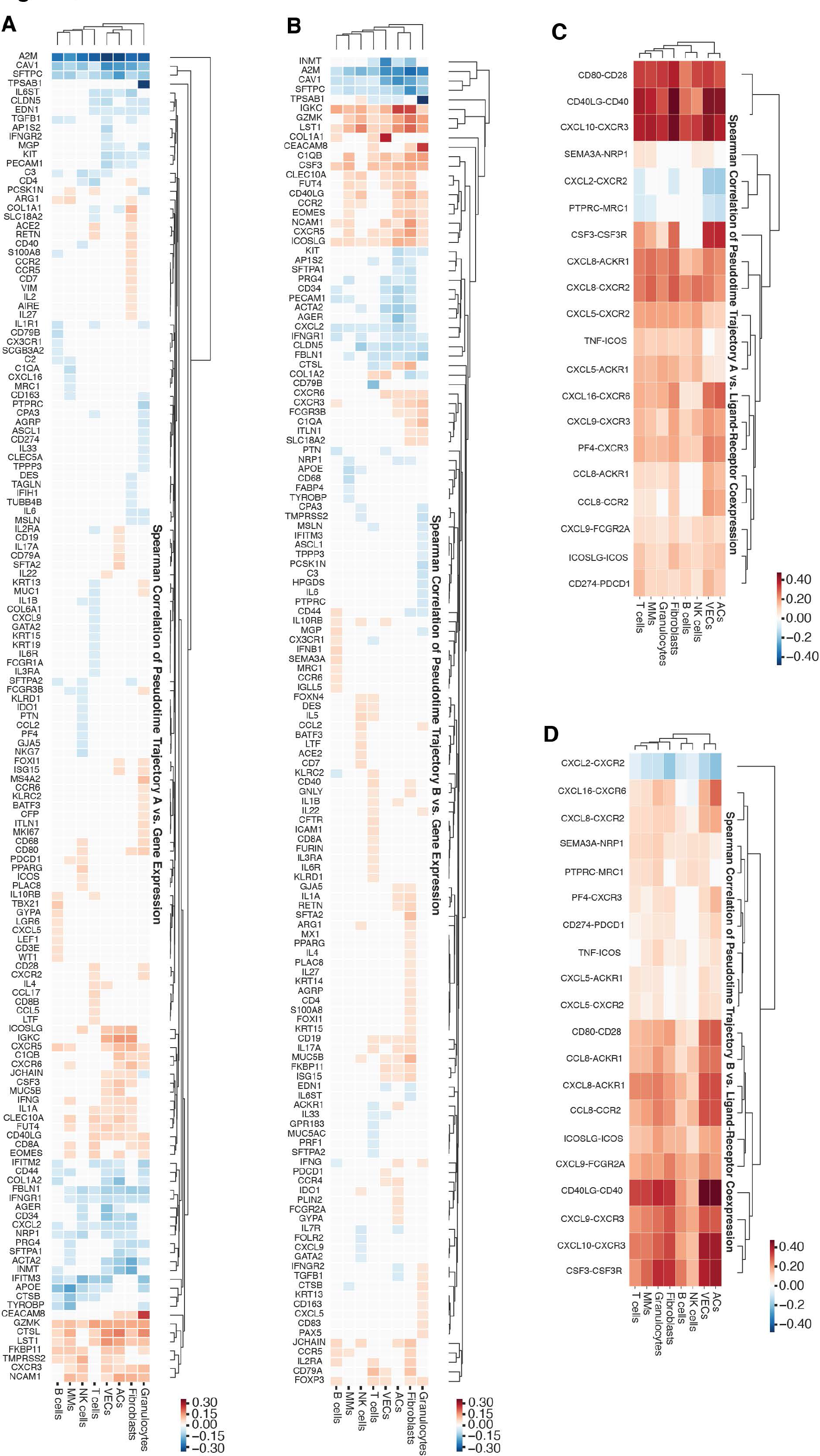
**A.** Heatmap illustrating the Spearman correlation of the pseudotime scores of Trajectory A and the gene expression. **B.** Heatmap illustrating the Spearman correlation of the pseudotime scores of Trajectory B and the gene expression. **C.** Heatmap illustrating the Spearman correlation of the pseudotime scores of Trajectory A and the ligand-receptor co-expression values of each cell. **D.** Heatmap illustrating the Spearman correlation of the pseudotime scores of Trajectory B and the ligand-receptor co-expression values of each cell. In all plots, deeper red indicates a high positive correlation and deeper blue indicates a high negative correlation. We also apply a p-value threshold (p-values<0.5) to remove the genes with lesser correlations and the removed genes are painted white to indicate no correlation.

**Supplementary Figure 27:**
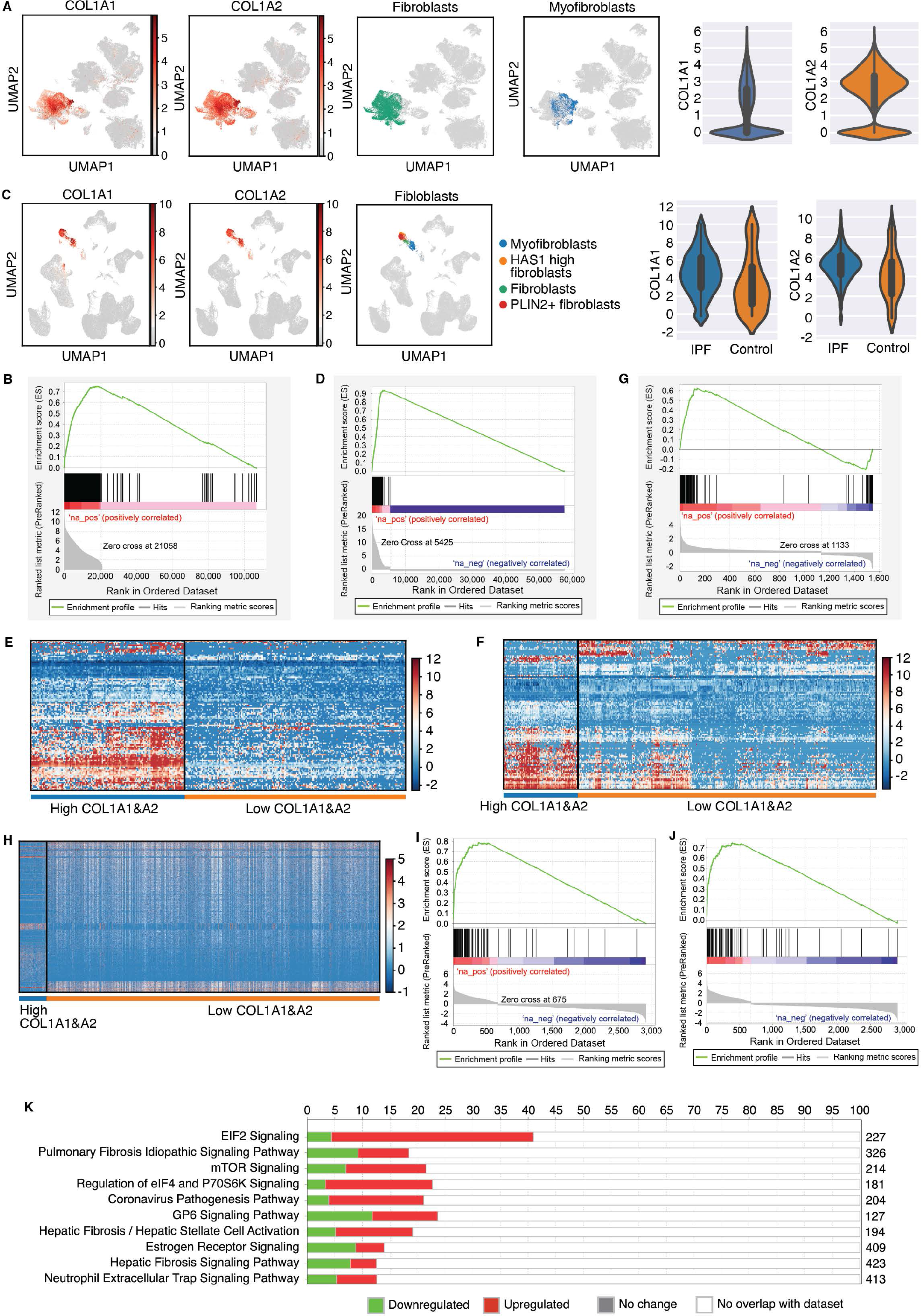
Analysis of COVID-19 and IPF single-cell RNA-seq datasets. A. UMAP projections showing expression of COL1A1 and COL1A2, paired with fibroblast and myofibroblast cell subtypes from single-cell RNA-seq COVID-19 tissue atlas. Violin plots showing expression of COL1A1 and COL1A2 in cells from COVID-19 patients. **B.** GSEA enrichment of myofibroblasts in cells ranked by expressions of COL1A1 and COL1A2 from the single-cell COVID-19 tissue atlas with a resulting p-value < 0.001. The significant enrichment and the large number of highly ranked hits indicate that most myofibroblasts from the single-cell COVID-19 tissue atlas have high expressions of COL1A1 and COL1A2. **C.** UMAP projections showing expressions of COL1A1 and COL1A2 next to UMAP of fibroblast cell subtypes from single-cell RNA-seq pulmonary fibrosis dataset. Violin plots showing expressions of COL1A1 and COL1A2 in IPF and control patients. The UMAPs and violin plots only represent cells from IPF patients or control. **D.** GSEA enrichment of myofibroblast in cells from IPF and control donors ranked by expressions of COL1A1 and COL1A2 with a resulting p-value < 0.001. The significant enrichment and high correlation between the myofibroblast cell type and high expressions of COL1A1 and COL1A2 indicate that most myofibroblasts from the single-cell pulmonary fibrosis dataset have high expressions of COL1A1 and COL1A2. **E.** A heatmap showing expressions of significantly differentially expressed genes from the ‘IPF High’ run in cells with high and low expressions of COL1A1 and COL1A2. **F.** A heatmap showing expressions of significantly differentially expressed genes from the ‘IPF Diagnosis’ cells with high and low expressions of COL1A1 and COL1A2. **G.** GSEA enrichment of significantly differentially expressed genes from the ‘IPF Diagnosis’ run against significantly differentially expressed genes ordered by fold change from the ‘IPF High’ run with a resulting p-value < 0.001. The high enrichment of upregulated genes and downregulated genes from the ‘IPF Diagnosis’ run in the ‘IPF High’ run indicates that the two differential expression analyses are capturing similar expression patterns. **H.** A heatmap showing expression of differentially expressed genes in cells with high and low COL1A1 and COL1A2 expressions from the single-cell COVID-19 dataset. **I.** GSEA enrichment of significantly differentially expressed genes from the ‘IPF High’ run in significantly differentially expressed genes from the COVID-19 dataset with a resulting p-value < 0.001. The high ranking of the ‘IPF High’ genes indicates that similar genes co-express with COL1A1 and COL1A2 in both COIVID-19 and IPF when considering IPF and control samples. **J.** GSEA enrichment of differentially expressed genes from ‘IPF Diagnosis’ against differentially expressed genes from the ’COVID High’ run ordered by fold change with a resulting p-value < 0.001. The high ranking of the ‘IPF Diagnosis’ genes indicates that the genes that co-express with COL1A1 and COL1A2 in COVID-19 have similar co-expression patterns in IPF. **K.** Top 10 enriched pathways from IPA pathway analysis for the differentially expressed genes from the COVID-19 single-cell RNA-seq dataset.

## Notes

### Competing Interest Statement

The authors have declared no competing interest.

### Author Declarations

The University of Pittsburgh IRB determined that the study is not research involving human subjects as defined by DHHS and FDA regulations and waived of ethical oversight (STUDY20050085).

## Reference

1. Ren, X., et al., COVID-19 immune features revealed by a large-scale single-cell transcriptome atlas. Cell, 2021. 184(23): p. 5838.

2. Melms, J.C., et al., A molecular single-cell lung atlas of lethal COVID-19. Nature, 2021. 595(7865): p. 114-119.

3. Wilk, A.J., et al., A single-cell atlas of the peripheral immune response in patients with severe COVID-19. Nat Med, 2020. 26(7): p. 1070–1076.

4. Delorey, T.M., et al., COVID-19 tissue atlases reveal SARS-CoV-2 pathology and cellular targets. Nature, 2021. 595(7865): p. 107-113.

5. Stephenson, E., et al., Single-cell multi-omics analysis of the immune response in COVID-19. Nat Med, 2021. 27(5): p. 904–916.

6. Erjefalt, J.S., et al., Diffuse alveolar damage patterns reflect the immunological and molecular heterogeneity in fatal COVID-19. EBioMedicine, 2022. 83: p. 104229.

7. Asp, M., J. Bergenstrahle, and J. Lundeberg, Spatially Resolved Transcriptomes- Next Generation Tools for Tissue Exploration. Bioessays, 2020. 42(10): p. e1900221.

8. Larsson, L., J. Frisen, and J. Lundeberg, Spatially resolved transcriptomics adds a new dimension to genomics. Nat Methods, 2021. 18(1): p. 15–18.

9. Ramos da Silva, S., et al., Broad Severe Acute Respiratory Syndrome Coronavirus 2 Cell Tropism and Immunopathology in Lung Tissues From Fatal Coronavirus Disease 2019. J Infect Dis, 2021. 223(11): p. 1842–1854.

10. Petukhov, V., et al., Cell segmentation in imaging-based spatial transcriptomics. Nat Biotechnol, 2022. 40(3): p. 345–354.

11. Hafemeister, C. and R. Satija, Normalization and variance stabilization of single- cell RNA-seq data using regularized negative binomial regression. Genome Biol, 2019. 20(1): p. 296.

12. Arish, M., et al., COVID-19 immunopathology: From acute diseases to chronic sequelae. J Med Virol, 2022. 95(1): p. e28122.

13. Whitsett, J.A., S.E. Wert, and T.E. Weaver, Alveolar surfactant homeostasis and the pathogenesis of pulmonary disease. Annu Rev Med, 2010. 61: p. 105–19.

14. Lin, Z., et al., Genetic Association of Pulmonary Surfactant Protein Genes, SFTPA1, SFTPA2, SFTPB, SFTPC, and SFTPD With Cystic Fibrosis. Front Immunol, 2018. 9: p. 2256.

15. Floros, J., et al., Human Surfactant Protein SP-A1 and SP-A2 Variants Differentially Affect the Alveolar Microenvironment, Surfactant Structure, Regulation and Function of the Alveolar Macrophage, and Animal and Human Survival Under Various Conditions. Front Immunol, 2021. 12: p. 681639.

16. Mikerov, A.N., et al., Inhibition of hemagglutination activity of influenza A viruses by SP-A1 and SP-A2 variants expressed in CHO cells. Med Microbiol Immunol, 2008. 197(1): p. 9–12.

17. Garcia-Laorden, M.I., et al., The role of mannose-binding lectin in pneumococcal infection. Eur Respir J, 2013. 41(1): p. 131–9.

18. Sawada, K., et al., Pulmonary collectins protect macrophages against pore- forming activity of Legionella pneumophila and suppress its intracellular growth. J Biol Chem, 2010. 285(11): p. 8434–43.

19. Fassan, M., et al., Multi-Design Differential Expression Profiling of COVID-19 Lung Autopsy Specimens Reveals Significantly Deregulated Inflammatory Pathways and SFTPC Impaired Transcription. Cells, 2022. 11(6): p. 1011.

20. Perkins, T.N., M.L. Donnell, and T.D. Oury, The axis of the receptor for advanced glycation endproducts in asthma and allergic airway disease. Allergy, 2021. 76(5): p. 1350–1366.

21. Yue, Q., et al., Receptor for Advanced Glycation End Products (RAGE): A Pivotal Hub in Immune Diseases. Molecules, 2022. 27(15): p. 4922.

22. Koerich, S., et al., Receptors for Advanced Glycation End Products (RAGE): Promising Targets Aiming at the Treatment of Neurodegenerative Conditions. Curr Neuropharmacol, 2022. Sep 22. doi: 10.2174/1570159X20666220922153903.Online ahead of print

23. Yamaguchi, K., et al., Association of the RAGE/RAGE-ligand axis with interstitial lung disease and its acute exacerbation. Respir Investig, 2022. 60(4): p. 531–542.

24. Hofmann, M.A., et al., RAGE mediates a novel proinflammatory axis: a central cell surface receptor for S100/calgranulin polypeptides. Cell, 1999. 97(7): p. 889–901.

25. Kapandji, N., et al., Importance of Lung Epithelial Injury in COVID-19-associated Acute Respiratory Distress Syndrome: Value of Plasma Soluble Receptor for Advanced Glycation End-Products. Am J Respir Crit Care Med, 2021. 204(3): p. 359–362.

26. Chiappalupi, S., et al., Hyperactivated RAGE in Comorbidities as a Risk Factor for Severe COVID-19-The Role of RAGE-RAS Crosstalk. Biomolecules, 2021. 11(6): p. 876.

27. Zhang, K. and S.H. Phan, Cytokines and pulmonary fibrosis. Biol Signals, 1996. 5(4): p. 232–9.

28. Zhu, Y., et al., Calcium in Vascular Smooth Muscle Cell Elasticity and Adhesion: Novel Insights Into the Mechanism of Action. Front Physiol, 2019. 10: p. 852.

29. Ringvold, H.C. and R.A. Khalil, Protein Kinase C as Regulator of Vascular Smooth Muscle Function and Potential Target in Vascular Disorders. Adv Pharmacol, 2017. 78: p. 203–301.

30. Morita, K., et al., Endothelial claudin: claudin-5/TMVCF constitutes tight junction strands in endothelial cells. J Cell Biol, 1999. 147(1): p. 185–94.

31. Schlingmann, B., S.A. Molina, and M. Koval, Claudins: Gatekeepers of lung epithelial function. Semin Cell Dev Biol, 2015. 42: p. 47–57.

32. Williams, M.R., et al., Gene expression of endothelial cells due to interleukin-1 beta stimulation and neutrophil transmigration. Endothelium, 2008. 15(1): p. 73–84.

33. Hashimoto, R., et al., SARS-CoV-2 disrupts respiratory vascular barriers by suppressing Claudin-5 expression. Sci Adv, 2022. 8(38): p. eabo6783.

34. Pang, L., et al., Role of caveolin-1 in human organ function and disease: friend or foe? Carcinogenesis, 2022. 43(1): p. 2–11.

35. Gokani, S. and L.K. Bhatt, Caveolin-1: A Promising Therapeutic Target for Diverse Diseases. Curr Mol Pharmacol, 2022. 15(5): p. 701–715.

36. Lagrange, J., et al., Alpha-2-macroglobulin in hemostasis and thrombosis: An underestimated old double-edged sword. J Thromb Haemost, 2022. 20(4): p. 806–815.

37. Vandooren, J. and Y. Itoh, Alpha-2-Macroglobulin in Inflammation, Immunity and Infections. Front Immunol, 2021. 12: p. 803244.

38. Mathison, R., J.S. Davison, and A.D. Befus, Neural regulation of neutrophil involvement in pulmonary inflammation. Comp Biochem Physiol C Comp Pharmacol Toxicol, 1993. 106(1): p. 39–48.

39. Sahu, S.K., et al., Emerging roles of the complement system in host-pathogen interactions. Trends Microbiol, 2022. 30(4): p. 390–402.

40. Magro, C., et al., Complement associated microvascular injury and thrombosis in the pathogenesis of severe COVID-19 infection: A report of five cases. Transl Res, 2020. 220: p. 1–13.

41. Mastaglio, S., et al., The first case of COVID-19 treated with the complement C3 inhibitor AMY-101. Clin Immunol, 2020. 215: p. 108450.

42. Afzali, B., et al., The state of complement in COVID-19. Nat Rev Immunol, 2022. 22(2): p. 77–84.

43. Freeley, S., et al., Asparaginyl Endopeptidase (Legumain) Supports Human Th1 Induction via Cathepsin L-Mediated Intracellular C3 Activation. Front Immunol, 2018. 9: p. 2449.

44. Zizzo, G., et al., Immunotherapy of COVID-19: Inside and Beyond IL-6 Signalling. Front Immunol, 2022. 13: p. 795315.

45. Zhao, M.M., et al., Cathepsin L plays a key role in SARS-CoV-2 infection in humans and humanized mice and is a promising target for new drug development. Signal Transduct Target Ther, 2021. 6(1): p. 134.

46. Trapani, J.A. and M.J. Smyth, Killing by cytotoxic T cells and natural killer cells: multiple granule serine proteases as initiators of DNA fragmentation. Immunol Cell Biol, 1993. 71 **(Pt** **3**): p. 201–8.

47. Berezin, V., et al., Targeting of ECM molecules and their metabolizing enzymes and receptors for the treatment of CNS diseases. Prog Brain Res, 2014. 214: p. 353–88.

48. Pimentel, E., Colony-stimulating factors. Ann Clin Lab Sci, 1990. 20(1): p. 36–55.

49. Venet, F., et al., Myeloid cells in sepsis-acquired immunodeficiency. Ann N Y Acad Sci, 2021. 1499(1): p. 3–17.

50. Roussel, L. and D.C. Vinh, ICOSL in host defense at epithelial barriers: lessons from ICOSLG deficiency. Curr Opin Immunol, 2021. 72: p. 21–26.

51. Panneton, V., et al., Inducible T-cell co-stimulator: Signaling mechanisms in T follicular helper cells and beyond. Immunol Rev, 2019. 291(1): p. 91–103.

52. Mulcahy, H., et al., LST1 and NCR3 expression in autoimmune inflammation and in response to IFN-gamma, LPS and microbial infection. Immunogenetics, 2006. 57(12): p. 893–903.

53. Heidemann, J., et al., Regulated expression of leukocyte-specific transcript (LST) 1 in human intestinal inflammation. Inflamm Res, 2014. 63(7): p. 513–7.

54. Chen, Z., X. Qiu, and J. Gu, Immunoglobulin expression in non-lymphoid lineage and neoplastic cells. Am J Pathol, 2009. 174(4): p. 1139–48.

55. Vettermann, C. and M.S. Schlissel, Allelic exclusion of immunoglobulin genes: models and mechanisms. Immunol Rev, 2010. 237(1): p. 22–42.

56. Napodano, C., et al., Mono/polyclonal free light chains as challenging biomarkers for immunological abnormalities. Adv Clin Chem, 2022. 108: p. 155–209.

57. Godyna, S., M. Diaz-Ricart, and W.S. Argraves, Fibulin-1 mediates platelet adhesion via a bridge of fibrinogen. Blood, 1996. 88(7): p. 2569–77.

58. Harikrishnan, K., et al., Cell Derived Matrix Fibulin-1 Associates With Epidermal Growth Factor Receptor to Inhibit Its Activation, Localization and Function in Lung Cancer Calu-1 Cells. Front Cell Dev Biol, 2020. 8: p. 522.

59. Jackson, C.B., et al., Mechanisms of SARS-CoV-2 entry into cells. Nat Rev Mol Cell Biol, 2022. 23(1): p. 3–20.

60. Felsenstein, S., et al., COVID-19: Immunology and treatment options. Clin Immunol, 2020. 215: p. 108448.

61. Guizani, I., et al., SARS-CoV-2 and pathological matrix remodeling mediators. Inflamm Res, 2021. 70(8): p. 847–858.

62. Zhang, F., et al., SARS-CoV-2 pseudovirus infectivity and expression of viral entry-related factors ACE2, TMPRSS2, Kim-1, and NRP-1 in human cells from the respiratory, urinary, digestive, reproductive, and immune systems. J Med Virol, 2021. 93(12): p. 6671–6685.

63. DeBruine, Z.J., K. Melcher, and T.J. Triche Jr, Fast and robust non-negative matrix factorization for single-cell experiments. bioRxiv, 2021: p. 2021.09. 01.458620.

64. Street, K., et al., Slingshot: cell lineage and pseudotime inference for single-cell transcriptomics. BMC Genomics, 2018. 19(1): p. 477.

65. Sweeney, R.M. and D.F. McAuley, Acute respiratory distress syndrome. Lancet, 2016. 388(10058): p. 2416–2430.

66. Wynn, T.A., Cellular and molecular mechanisms of fibrosis. J Pathol, 2008. 214(2): p. 199–210.

67. Turpin, E.A., et al., Respiratory syncytial virus infection reduces lung inflammation and fibrosis in mice exposed to vanadium pentoxide. Respir Res, 2010. 11: p. 20.

68. Elewa, Y.H.A., et al., Histopathological Correlations between Mediastinal Fat- Associated Lymphoid Clusters and the Development of Lung Inflammation and Fibrosis following Bleomycin Administration in Mice. Front Immunol, 2018. 9: p. 271.

69. Nagata, N., et al., Features of idiopathic pulmonary fibrosis with organizing pneumonia. Respiration, 1997. 64(5): p. 331–5.

70. Phan, S.H., The myofibroblast in pulmonary fibrosis. Chest, 2002. 122(6 Suppl): p. 286S–289S.

71. Ortiz-Zapater, E., et al., Lung Fibrosis and Fibrosis in the Lungs: Is It All about Myofibroblasts? Biomedicines, 2022. 10(6).

72. Habermann, A.C., et al., Single-cell RNA sequencing reveals profibrotic roles of distinct epithelial and mesenchymal lineages in pulmonary fibrosis. Sci Adv, 2020. 6(28): eaba1972.

73. Laloglu, E. and H. Alay, Role of transforming growth factor-beta 1 and connective tissue growth factor levels in coronavirus disease-2019-related lung Injury: a prospective, observational, cohort study. Rev Soc Bras Med Trop, 2022. 55: p. e06152021.

74. Vaz de Paula, C.B., et al., COVID-19: Immunohistochemical Analysis of TGF- beta Signaling Pathways in Pulmonary Fibrosis. Int J Mol Sci, 2021. 23(1): p. 168.

75. Fan, T., et al., Discovery of 9O-Substituted Palmatine Derivatives as a New Class of anti-COL1A1 Agents via Repressing TGF-beta1/Smads and JAK1/STAT3 Pathways. Molecules, 2020. 25(4): p.773.

76. Malik, M., et al., Cross-talk between Janus kinase-signal transducer and activator of transcription pathway and transforming growth factor beta pathways and increased collagen1A1 production in uterine leiomyoma cells. F S Sci, 2020. 1(2): p. 206–220.

77. Stringer, C., et al., Cellpose: a generalist algorithm for cellular segmentation. Nat Methods, 2021. 18(1): p. 100–106.

78. Qi, F., et al., Single cell RNA sequencing of 13 human tissues identify cell types and receptors of human coronaviruses. Biochem Biophys Res Commun, 2020. 526(1): p. 135–140.

79. Sungnak, W., et al., SARS-CoV-2 entry factors are highly expressed in nasal epithelial cells together with innate immune genes. Nat Med, 2020. 26(5): p. 681–687.

80. Xu, G., et al., The differential immune responses to COVID-19 in peripheral and lung revealed by single-cell RNA sequencing. Cell Discov, 2020. 6: p. 73.

81. Zou, X., et al., Single-cell RNA-seq data analysis on the receptor ACE2 expression reveals the potential risk of different human organs vulnerable to 2019-nCoV infection. Front Med, 2020. 14(2): p. 185–192.

82. Sounart, H., et al., Dual spatially resolved transcriptomics for SARS-CoV-2 host- pathogen colocalization studies in humans. bioRxiv, 2022: p. 2022.03. 14.484288.

83. Pielawski, N., et al., TissUUmaps 3: Interactive visualization and quality assessment of large-scale spatial omics data. BioRxiv, 2022. doi: https://doi.org/10.1101/2022.01.28.478131.

84. Solorzano, L., G. Partel, and C. Wahlby, TissUUmaps: interactive visualization of large-scale spatial gene expression and tissue morphology data. Bioinformatics, 2020. 36(15): p. 4363–4365.

85. Satija, R., et al., Spatial reconstruction of single-cell gene expression data. Nat Biotechnol, 2015. 33(5): p. 495–502.

86. Anselin, L., Local indicators of spatial association—LISA. Geographical analysis, 1995. 27(2): p. 93–115.

